# Development and validation of a machine learning model for prediction of type 2 diabetes in patients with mental illness

**DOI:** 10.1101/2023.09.26.23296141

**Authors:** Martin Bernstorff, Lasse Hansen, Kenneth Enevoldsen, Jakob Damgaard, Frida Hæstrup, Erik Perfalk, Andreas Aalkjær Danielsen, Søren Dinesen Østergaard

## Abstract

**Background:** Type 2 diabetes (T2D) is approximately twice as common among individuals with mental illness compared with the background population, but may be prevented by early intervention on lifestyle, diet, or pharmacologically. Such prevention relies on identification of those at elevated risk (prediction). The aim of this study was to develop and validate a machine learning model for prediction of T2D among patients with mental illness.

**Methods:** The study was based on routinely collected data from electronic health records from the psychiatric services of the Central Denmark Region. A total of 74.880 patients with 1.59 million psychiatric service contacts were included in the analyses. We included 1343 potential predictors covering patient-level information on demographics, diagnoses, pharmacological treatment, and laboratory results. T2D was operationalized as HbA1c ≥48 mmol/mol, fasting plasma glucose >7.0 mmol/mol, oral glucose tolerance test ≥11.1 mmol/mol or random plasma glucose ≥11.1 mmol/mol. Two machine learning models (XGBoost and regularized logistic regression) were trained to predict T2D based on 85% of the included contacts. The predictive performance of the best performing model was tested on the remaining 15% of the contacts.

**Findings:** The XGBoost model detected patients at high risk 2.7 years before T2D, achieving an area under the receiver operating characteristic curve of 0.84. Of the 996 patients developing T2D in the test set, the model issued at least one positive prediction for 305 (31%).

**Interpretation:** A machine learning model can accurately predict development of T2D among patients with mental illness based on routinely collected data from electronic health records. A decision support system based on such a model may inform measures to prevent development of T2D in this high-risk population.

**Funding:** The Lundbeck Foundation, the Central Denmark Region Fund for Strengthening of Health Science and the Danish Agency for Digitisation Investment Fund for New Technologies.

**Research in context:** *Evidence before this study:* We searched Pubmed for relevant studies regardless of time of publication using the search query “predict*” AND Diabetes Mellitus, Type 2 [Mesh] AND Mental Disorders [Mesh] AND Patients [Mesh]. We did not identify any studies developing T2D prediction models for patients with mental illness.

*Added value of this study:* To the best of our knowledge, this study is the first to develop and validate a machine learning model for prediction of T2D among patients with mental illness. The developed model is sensitive and specific - and detects patients at high risk 2.7 years before T2D. Notably, as only routinely collected data from electronic health records were used in the training of the model training, it can be assumed to have similar predictive performance if implemented in clinical practice. This study adds value by offering a T2D prediction model tailored specifically to patients with mental illness, which may facilitate early intervention and prevention strategies.

*Implications of all the available evidence:* The findings of this study, combined with the absence of existing T2D prediction models for patients with mental illness in the literature, offer a new possibility for identifying and potentially preventing T2D in a high-risk population. Specifically, implementing such a system in clinical practice may inform targeted interventions, such as lifestyle modifications (e.g., exercise and diet) and pharmacological treatment, to reduce the risk of T2D.

## Introduction

The prevalence of type 2 diabetes (T2D) is on the rise worldwide.^1^ As T2D is associated with both decreased quality of life and reduced life expectancy, this development is a cause of great concern.^2^ The risk of developing T2D is particularly elevated among individuals with mental illness. Indeed, among patients treated for mental illness at psychiatric hospitals, T2D prevalence estimates range from 10-20%.^3^ There are multiple causes underlying this overrepresentation. For example, individuals with mental illness tend to have an unhealthy lifestyle, e.g., poor diet, lack of physical activity and high alcohol intake.^3^ Moreover, psychopharmacological treatment also plays a causal role in the development of T2D, partly mediated via its effects on weight.^4^

Among patients with elevated risk, development of T2D can be prevented by early interventions on lifestyle and diet, or pharmacologically.^5^ The first step towards prevention of T2D is, therefore, to identify those at elevated risk of this condition. However, estimating risk of T2D is a complex task and, to our knowledge, no models are available for prediction of T2D among individuals with mental illness.

Predictors of increased risk of T2D among individuals with mental illness are likely many and probably interact, a context in which machine learning models perform particularly well since they are designed to allow for high complexity, while disregarding idiosyncrasies in the data.^6^ Indeed, prior studies have shown that machine learning models can accurately predict important clinical outcomes for patients with mental illness when trained on routinely collected data from electronic health records.^7,8^ Therefore, to aid health care workers in identifying patients suitable for intervention, we investigated whether a machine learning model trained on data from electronic health records can predict development of T2D among patients with mental illness.

## Methods

Reporting followed all proposed items which were rated “essential for inclusion” by ≥50% of the respondents in the first Delphi round for the Transparent Reporting of multivariable prediction models for Individual Prognosis Or Diagnosis with Artificial Intelligence (TRIPOD-AI).^9^

### Data source

The study is based on the PSYchiatric Clinical Outcome Prediction (PSYCOP) cohort,^10^ which contains routinely collected electronic health record data from all individuals with at least one contact to the Psychiatric Services of the Central Denmark Region in the period from January 1, 2011 to November 22, 2021. The data cover all contacts to the five public hospitals in the Central Denmark Region (both psychiatric and somatic departments). As Denmark has universal healthcare, the vast majority of hospital contacts are to public hospitals and, hence, covered by these data. Notably, blood samples from general practitioners are analysed at the public hospitals and are included in the dataset.

### Cohort definition

A flowchart illustrating the cohort definition is available in Supplementary Figure 1. Due to data instability prior to 2013 caused by the gradual implementation of a new electronic health record system in 2011,^11,12^ we restricted the cohort to contacts after January 1, 2013. Furthermore, we only included patients aged 18 years or older as registration practices in child and adolescent psychiatry differs from that of adult psychiatry in Denmark. Finally, to avoid issuing predictions when a diagnosis of T2D had already been made, we excluded all contacts for patients with known T2D – defined as meeting one of the following criteria in the period from January 1, 2011, to December 31, 2013:

1. A laboratory result indicative of diabetes. For the definition, see the list from 1 to 4 under “Definition of outcome (T2D)”, below.
2. Receiving a hospital diagnosis of either type 1 or type 2 diabetes (Supplementary Table 1)
3. Receiving treatment with antidiabetic medication (Anatomical Therapeutic Chemical (ATC) code: A10) in the hospital.

### Definition of outcome (T2D)

Guidelines from the World Health Organization (WHO) and medical organizations from the United States, United Kingdom, and Denmark define T2D as follows:^13–17^

1. Glycosylated haemoglobin (HbA1c) ≥48 mmol/mol, or
2. Fasting plasma glucose ≥7.0 mmol/l, or
3. Plasma glucose ≥11.1 mmol/l two hours after an oral glucose tolerance test, or
4. Random plasma glucose ≥11.1 mmol/l

Accompanied by diabetes symptoms or measured on two separate occasions.

We used this definition of T2D for the present study with the following modifications: As the data does not allow us to determine whether a laboratory result was accompanied by diabetes symptoms, this requirement was excluded. Furthermore, in agreement with the WHO recommendation on observational studies of T2D,^16^ we only required one laboratory result (from the list from 1 to 4) above threshold for a patient to be defined as having developed T2D.

### Data pre-processing and model training

Figure 1 illustrates the extraction of data, outcome extraction, dataset splitting, prediction-time filtering, specification of predictors and flattening, and the model training, testing and evaluation pipeline.

**Figure 1:**
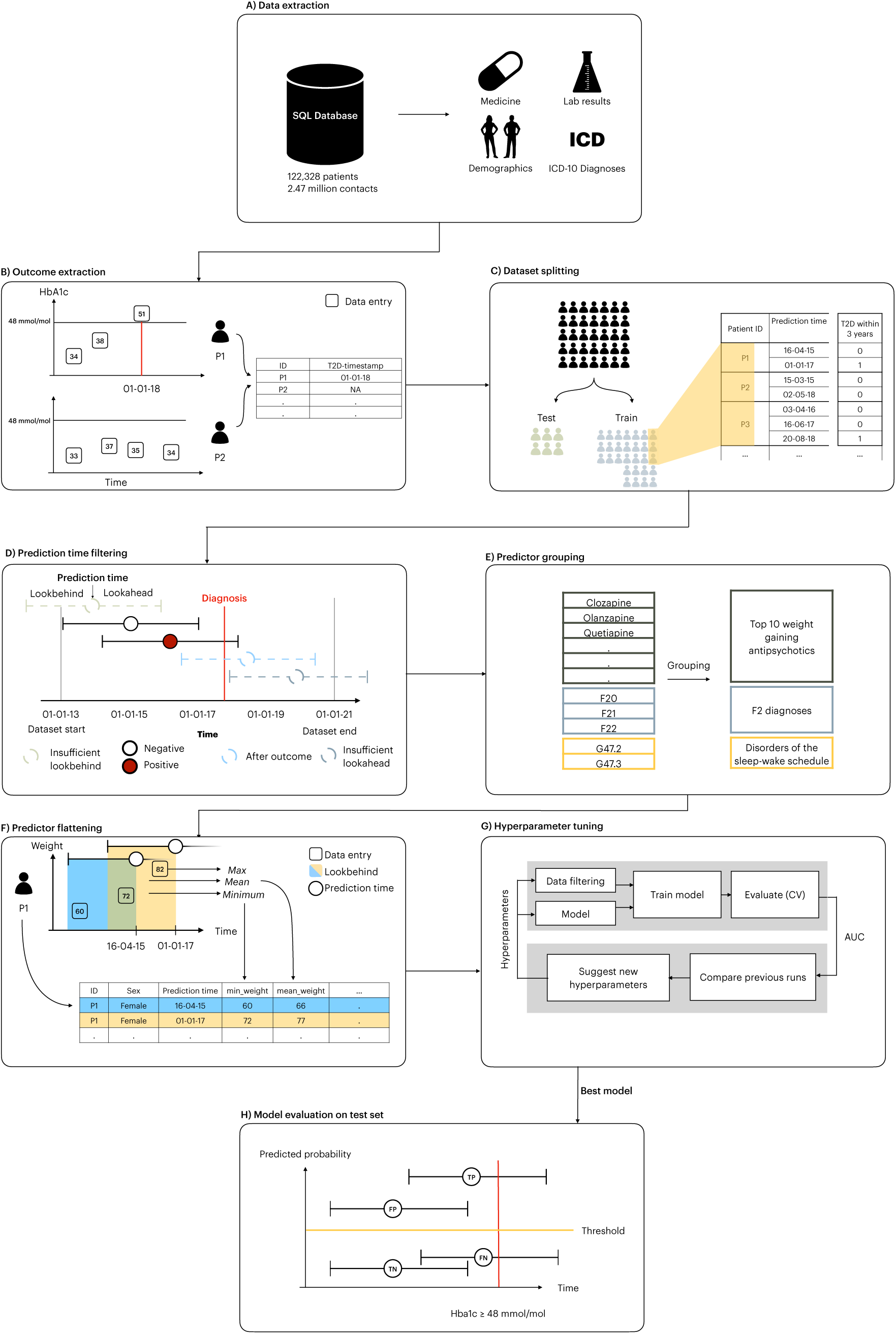
Extraction of data and outcome, dataset splitting, prediction time filtering, specification of predictors and flattening, model training, testing and evaluation A: Data was extracted from the electronic health records B: Potential T2D was identified C: The dataset obtained is randomly split into an independent training dataset (85%) and test dataset (15%) stratified on development of T2D with no patient being present in both groups. D: Prediction times were removed if their lookbehind window extended beyond the start of the dataset or their lookahead extended beyond the end of the dataset. Prediction times were also removed after a patient developed T2D. E: Predictors were grouped. The grouping only renamed the predictor name, the number of observations was held constant. F: Predictors for each prediction time were extracted by aggregating the variables within the lookbehind with multiple aggregation functions. As a result, each row in the dataset represents a specific prediction time with a column for each predictor. G: Models were trained and optimized on the training set using 5-fold cross-validation. Hyperparameters were tuned to optimize AUROC. H: The best candidate model was evaluated on the independent test set. True positive predictions were those with predicted probabilities above the decision threshold and the patient developing T2D within the lookahead window. False positive predictions were those where the model’s predicted probability was above the decision threshold, but where none of the T2D criteria were satisfied within the lookahead window. False negatives had predicted probabilities below the threshold, but the T2D criteria were met within the lookahead window. True negatives had predicted probabilities below the threshold, and the patient did not satisfy T2D criteria within the lookahead window.

### Data extraction

All electronic health record data for patients with at least one contact to the Psychiatric Services of the Central Denmark Region were extracted (Figure 1A). To facilitate potential implementation of a predictive machine learning model, we only used data that are routinely collected.

### Outcome extraction

To define outcome labels, we extracted whether patients met the study definition of T2D, and then calculated whether it occurred within the “lookahead window” of a given contact (Figure 1B). By lookahead window, we refer to the fact that we trained models predicting T2D 1, 2, 3, 4 and 5 years ahead from each prediction time (psychiatric service contacts), respectively, to find the best trade-off between early prediction (longer lookahead windows) and model accuracy (shorter lookahead windows).

### Dataset splitting

Subsequently, the data were randomly split into a training dataset (85%) and a test dataset (15%) by sampling patients, stratified by whether they met the definition of T2D during follow-up (Figure 1C). This ensured that the training and test dataset contained an even proportion of T2D patients, and that no patient occurred in both datasets. Hereafter, no information from the test dataset was examined until the final model evaluation (the test of the optimal model obtained during the training phase).

### Prediction time filtering

To ensure that predictions were issued at a time where intervention was possible, we defined prediction times as the time of any in- or outpatient contact to the Psychiatric Services (service contacts). Hence, a single patient had as many prediction times as the number of service contacts. Predictions were restricted to patients who had not already met criteria for T2D at the time of a contact (see “Definition of Outcome”, above) and to contacts for which the lookahead and lookbehind were not truncated by the dataset limits (Figure 1D). Specifically, a prediction was not issued if the lookbehind window included time before the start of follow-up (January 1, 2013). By lookbehind window, we refer to the time (past) used for predictor extraction. Similarly, a prediction was not issued if the lookahead window extended beyond the end of follow-up, the date of moving out of the Central Denmark Region, or death. These ’truncations’ are artifacts caused by data collection and could lead the model to learn patterns that do not exist during implementation, resulting in discrepancies between the model’s performance during testing and its actual performance when implemented. Finally, to remove contacts where the patient had been diagnosed with T2D outside of the dataset (prevalent cases), only contacts where the patient had lived in the Central Denmark Region for 2 years (wash-in) were included.

### Predictor grouping and flattening

A full list of all 1,343 predictors and their definition/grouping is available in Supplementary Table 2 and illustrated in Figure 1E. The predictors were chosen as follows: First, we replicated available predictors from a recent meta-analysis of prediction models for T2D.^20^ Lab results included mSupplementary Tabolic markers such as hemoglobin A1c (HbA1c), high-density lipoprotein (HDL), low-density lipoprotein (LDL), triglycerides, and general markers found to be predictive in the meta-analysis, such as alanine aminotransferase (ALAT), C-reactive protein (CRP), and estimated glomerular filtration rate (eGFR). The following demographics were included: age, sex, weight, height, and Body Mass Index (BMI). Of note, the dataset does not include structured registration of physical activity or dietary habits, so these could not be considered as predictors. With regard to diagnoses, we included all psychiatric subchapters from the ICD-10 (F0-F9), as well as a selected set of diagnoses representing conditions known to be strongly associated with T2D, namely obstructive sleep apnea (ICD-10: G47.3), sleep disorders (ICD-10: G47.9), polycystic ovarian syndrome (ICD-10: E28.2), hyperlipidaemia (ICD-10: E78.0 and E78.5) and essential hypertension (ICD-10: I109).^20^ Antipsychotics were covered by three categories: I) The 10 antipsychotics that produce the largest weight gain (clozapine: N05AH02, zotepine: N05AX11, olanzapine: N05AH03, sertindole: N05AE03, chlorpromazine: N05AA01, iloperidone: N05AX14, quetiapine (N05AH04), paliperidone (N05AX13), trifluoperazine (N05AB06), and risperidone (N05AX08)), II) Clozapine (ATC-code: N05AH02), which was considered on its own due to its unique role in treatment-resistant schizophrenia, and III) all antipsychotics (ATC codes: N05A*, except N05AN01 (lithium)). This approach allows the model to pick up potential differential effects of these three categories. As mood stabilisers appear to have differential associations with type 2 diabetes,^4^ we included lithium (N05AN01), valproate (N03AG01), and lamotrigine (N03AX09) individually. Predictors were aggregated over the lookbehind windows using the “timeseriesflattener” Python package (Figure 1F).^21^ For further details on predictor specification and flattening, see the Supplementary Methods.

### Hyperparameter tuning

Due to the large number of possible model configurations, we chose to focus on two models, namely XGBoost and elastic net regularized logistic regression (Figure 1G).^22^ XGBoost was chosen as gradient boosting methods generally show superior predictive performance on tabular data, are fast to train, and they model missing values internally. Logistic regression using elastic net regularization was used as a strong baseline. Models were trained using 5-fold cross-validation. For each of the five lookahead windows (i.e., 1., 2, 3, 4 and 5 years), we conducted hyperparameter optimization to maximise the area under the receiver operating characteristic curve (AUROC) using the tree-structured parzen estimator algorithm implemented in Optuna v2.10.1. For further details, see Supplementary Table 3 and the Supplementary Methods.

### Model evaluation on test data

The model that achieved the best trade-off between AUROC and early detection of potential T2D in the training phase was evaluated on the test data (Figure 1H). Specifically, we calculated the AUROC for global performance, as well as sensitivity, specificity, positive predictive value, and negative predictive value. Regarding early detection, we calculated the mean time from the first positive prediction until a patient met the definition of T2D. All performance estimates were calculated for different “predicted positive rates” of 1%, 2%, 3%, 4% and 5%. The predicted positive rate is the proportion of all prediction times that are marked as positive. Predictor importance was estimated via information gain.^23^ In the case of XGBoost, the information gain of a predictor is the change in predicted probability at a given node split, averaged across all trees in the model.

### Sensitivity analyses

For a description of analyses of the stability of model predictions over I) time, II) patient characteristics, III) feature availability, and IV) whether the model is overly reliant on “clinical suspicion”, see the Supplementary Methods. To determine whether true negatives or false positives were driven by the T2D-defining laboratory tests not being carried out during follow-up, we plotted the cumulative proportion of the last test (and the last HbA1c test specifically) over time.

### Post hoc analysis

Based on the observation of approximately equal performance of the XGBoost models irrespective of lookahead, we also tested the performance of the other four models (1, 2, 3 and 4-year lookahead) on the test set. Furthermore, based on the results on information gain, where HbA1c features dominated, we tested a parsimonious model with only sex, age and mean HbA1c within the last 2 years as features.

### Ethics

The use of electronic health records from the Central Denmark Region for this study was approved by the Legal Office of the Central Denmark Region in accordance with the Danish Health Care Act §46, Section 2. According to the Danish Committee Act, ethical review board approval is not required for studies based solely on data from electronic health records (waiver for this project: 1-10-72-1-22). Data were processed and stored in accordance with the European Union General Data Protection Regulation and the project is registered on the internal list of research projects having the Central Denmark Region as data steward.

## Results

The eligible cohort consisted of 74,880 patients with a total of 1.6 million contacts. Demographic and clinical information of the cohort is shown in Table 1. The full dataset contained 1343 predictors covering routinely collected laboratory tests, diagnoses, and medications (Supplementary Table 2). The incidence of T2D was constant after exclusion, indicating successful filtering of prevalent cases (Supplementary Figure 2).

**Table 1.**
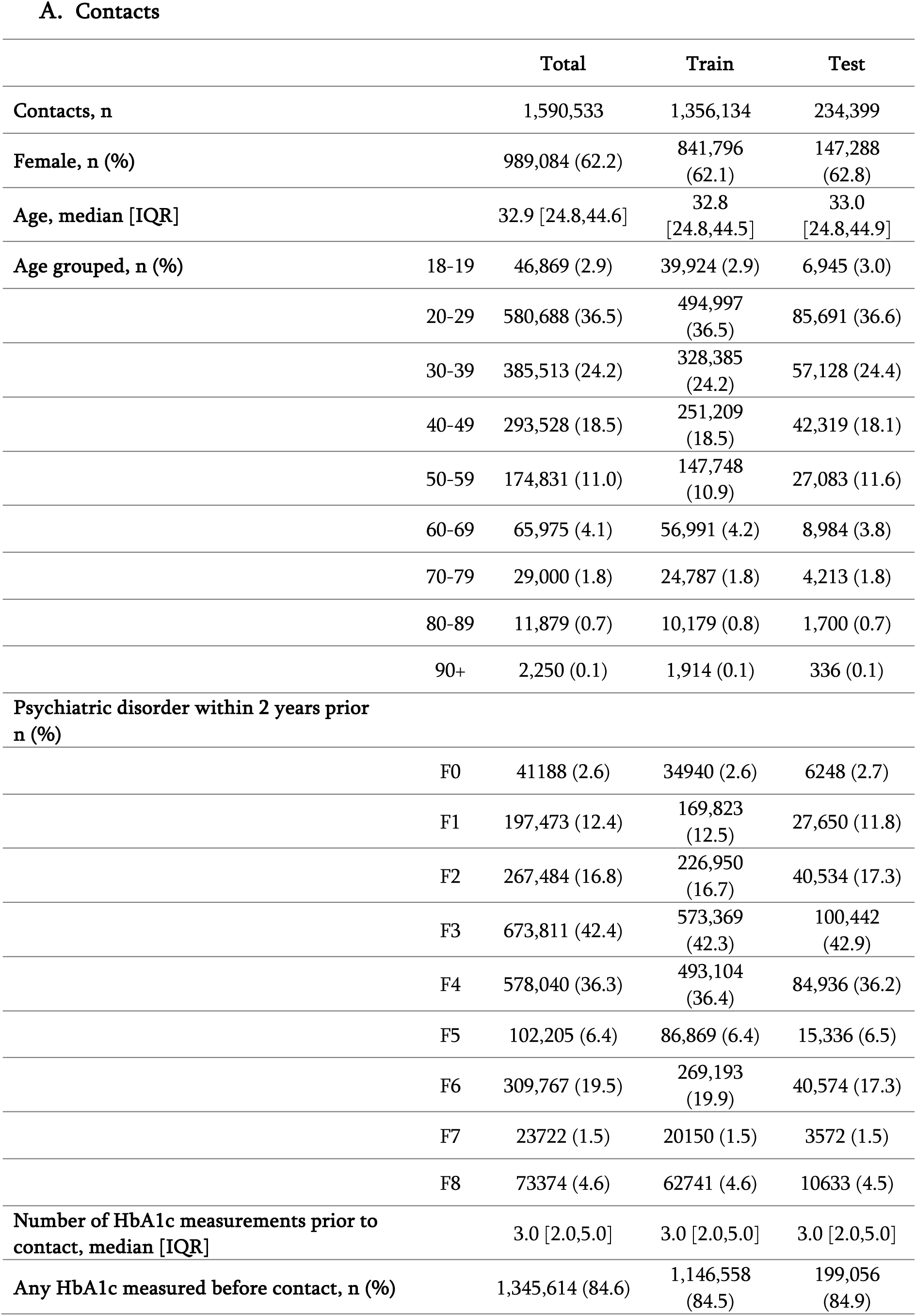
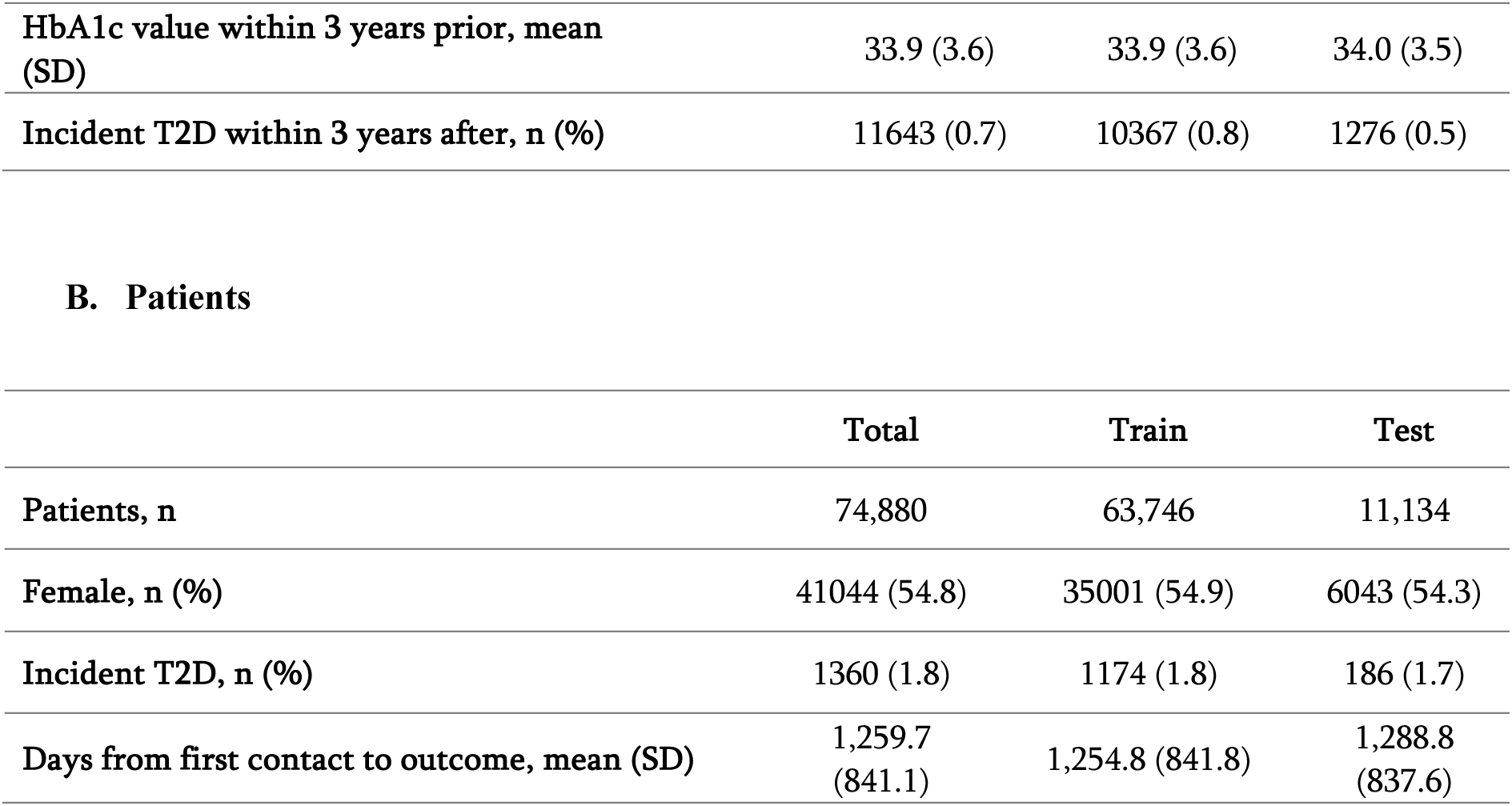
Descriptive statistics for contacts (A) and patients (B) that were eligible for prediction.

|  |  | **Total** | **Train** | **Test** |
| --- | --- | --- | --- | --- |
| **Contacts, n** |  | 1,590,533 | 1,356,134 | 234,399 |
| **Female, n (%)** |  | 989,084 (62.2) | 841,796 (62.1) | 147,288 (62.8) |
| **Age, median [IQR]** |  | 32.9 [24.8,44.6] | 32.8 [24.8,44.5] | 33.0 [24.8,44.9] |
| **Age grouped, n (%)** | 18-19 | 46,869 (2.9) | 39,924 (2.9) | 6,945 (3.0) |
|  | 20-29 | 580,688 (36.5) | 494,997 (36.5) | 85,691 (36.6) |
|  | 30-39 | 385,513 (24.2) | 328,385 (24.2) | 57,128 (24.4) |
|  | 40-49 | 293,528 (18.5) | 251,209 (18.5) | 42,319 (18.1) |
|  | 50-59 | 174,831 (11.0) | 147,748 (10.9) | 27,083 (11.6) |
|  | 60-69 | 65,975 (4.1) | 56,991 (4.2) | 8,984 (3.8) |
|  | 70-79 | 29,000 (1.8) | 24,787 (1.8) | 4,213 (1.8) |
|  | 80-89 | 11,879 (0.7) | 10,179 (0.8) | 1,700 (0.7) |
|  | 90+ | 2,250 (0.1) | 1,914 (0.1) | 336 (0.1) |
| **Psychiatric disorder within 2 years prior n (%)** |  |  |  |  |
|  | F0 | 41188 (2.6) | 34940 (2.6) | 6248 (2.7) |
|  | F1 | 197,473 (12.4) | 169,823 (12.5) | 27,650 (11.8) |
|  | F2 | 267,484 (16.8) | 226,950 (16.7) | 40,534 (17.3) |
|  | F3 | 673,811 (42.4) | 573,369 (42.3) | 100,442 (42.9) |
|  | F4 | 578,040 (36.3) | 493,104 (36.4) | 84,936 (36.2) |
|  | F5 | 102,205 (6.4) | 86,869 (6.4) | 15,336 (6.5) |
|  | F6 | 309,767 (19.5) | 269,193 (19.9) | 40,574 (17.3) |
|  | F7 | 23722 (1.5) | 20150 (1.5) | 3572 (1.5) |
|  | F8 | 73374 (4.6) | 62741 (4.6) | 10633 (4.5) |
| **Number of HbA1c measurements prior to contact, median [IQR]** |  | 3.0 [2.0,5.0] | 3.0 [2.0,5.0] | 3.0 [2.0,5.0] |
| **Any HbA1c measured before contact, n (%)** |  | 1,345,614 (84.6) | 1,146,558 (84.5) | 199,056 (84.9) |

|  |  |  |  |
| --- | --- | --- | --- |
| HbA1c value within 3 years prior, mean (SD) | 33.9 (3.6) | 33.9 (3.6) | 34.0 (3.5) |
| Incident T2D within 3 years after, n (%) | 11643 (0.7) | 10367 (0.8) | 1276 (0.5) |

B. Patients
|  | Total | Train | Test |
| --- | --- | --- | --- |
| Patients, n | 74,880 | 63,746 | 11,134 |
| Female, n (%) | 41044 (54.8) | 35001 (54.9) | 6043 (54.3) |
| Incident T2D, n (%) | 1360 (1.8) | 1174 (1.8) | 186 (1.7) |
| Days from first contact to outcome, mean (SD) | 1,259.7 (841.1) | 1,254.8 (841.8) | 1,288.8 (837.6) |

### Model training

As highlighted in Figure 2A, we estimated that the best trade-off between AUROC and early detection was for XGBoost with a 5-year lookahead. XGBoost outperformed logistic regression across all lookahead windows.

**Figure 2.**
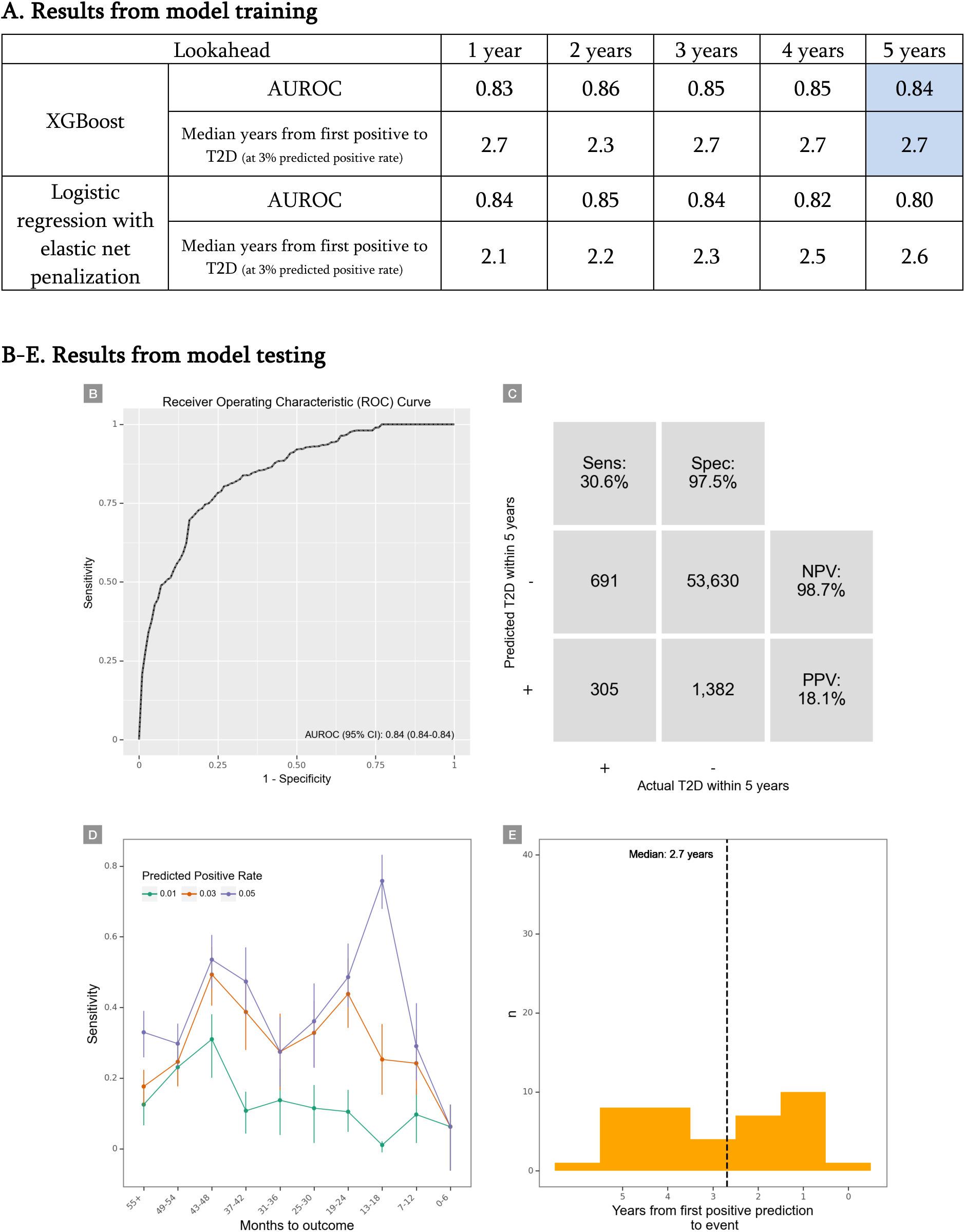
Results from model training (A) and test (B-E) A) Performance of the best performing models for each model type at every lookahead during the training phase. The cells marked in blue highlight the model that was run on the test-set, which was used in panels B-E. **B)** Receiver operating characteristics (ROC) curve. **C)** Confusion matrix. PPV: Positive predictive value. NPV: Negative predictive value. **D)** Sensitivity by months from prediction time to event, stratified by desired predicted positive rate (PPR). Note that the numbers do not match those in Table 1, since all prediction times with insufficient lookahead distance have been dropped. **E)** Time (months) from the first positive prediction to the patient developing T2D at a 3% predicted positive rate (PPR).

### Model testing

Figure 2B shows the results for the XGBoost model with a 5-year lookahead window applied to the test data. It achieved an AUROC of 0.84 (95% CI: 0.84; 0.84). Figure 2C shows the resulting confusion matrix at a predicted positive rate of 3% with a positive predictive value of 18% and a negative predictive value of 99%, reflecting that for every five positive predictions, one prediction time was followed by T2D within 5 years. At this predicted positive rate, the sensitivity at the level of prediction times was 31%, and 31% of all patients who developed T2D were predicted positive at least once (Table 2). Figure 2D shows the sensitivity of the model by time until T2D and by predicted positive rate, with no clear temporal trends. Figure 2E shows that the model marked patients as being at high risk an average of 2.7 years before they developed T2D.

**Table 2.** Performance by predicted positive rate for XGBoost with 5 years of lookahead on the test set.

| Predicted positive rate | True prevalence | PPV | NPV | Sensitivity | Specificity | FPR | FNR | Accuracy | TP | TN | FP | FN | % of all patients with T2D captured | Median years from first positive to T2D |
| --- | --- | --- | --- | --- | --- | --- | --- | --- | --- | --- | --- | --- | --- | --- |
| 5.0% | 1.8% | 14.8% | 98.9% | 41.6% | 95.7% | 4.3% | 58.4% | 94.7% | 414 | 52,625 | 2,387 | 582 | 40.1% | 3.2 |
| 4.0% |  | 15.4% | 98.8% | 34.7% | 96.5% | 3.5% | 65.3% | 95.4% | 346 | 53,113 | 1,899 | 650 | 34.0% | 2.8 |
| 3.0% |  | 18.1% | 98.7% | 30.6% | 97.5% | 2.5% | 69.4% | 96.3% | 305 | 53,630 | 1,382 | 691 | 31.0% | 2.7 |
| 2.0% |  | 20.6% | 98.6% | 23.3% | 98.4% | 1.6% | 76.7% | 97.0% | 232 | 54,117 | 895 | 764 | 19.2% | 2.7 |
| 1.0% |  | 27.5% | 98.5% | 15.5% | 99.3% | 0.7% | 84.5% | 97.8% | 154 | 54,605 | 407 | 842 | 14.0% | 2.5 |

**Predicted positive rate:** The proportion of contacts predicted positive by the model. Since the model outputs a predicted probability, this is a threshold set during evaluation.
**True prevalence:** The proportion of contacts that qualified for type 2 diabetes within the lookahead window.
**PPV:** Positive predictive value.
**NPV:** Negative predictive value.
**FPR:** False positive rate.
**FNR:** False negative rate.
**TP:** True positives. Numbers are service contacts.
**TN:** True negatives. Numbers are service contacts.
**FP:** False positives. Numbers are service contacts.

Supplementary Table 4 lists the 100 most important predictors by information gain. The 5 most important predictors based on information gain were the mean HbA1c within the last 2 years, the most recent weight within the last year, the maximal HbA1c within the last 2 years, the most recent triglyceride level within the last year and patient age at the time of prediction.

### Sensitivity analyses

Figure 3 highlights that the model performed acceptably irrespective of the sex and age of the patients. Moreover, it was stable across the number of HbA1c measurements prior to the contact and months since first contact, both proxies of how much information was available, as well as across months of the year and day of the week. When the model was only informed whether a measurement had been made (a proxy for clinical suspicion), not the value of the measurement, it performed relatively poorly with an AUROC of 0.60. Supplementary Figure 3 shows that true negatives and false positives were not driven by the T2D-defining laboratory tests not being carried out during follow-up.

**Figure 3.**
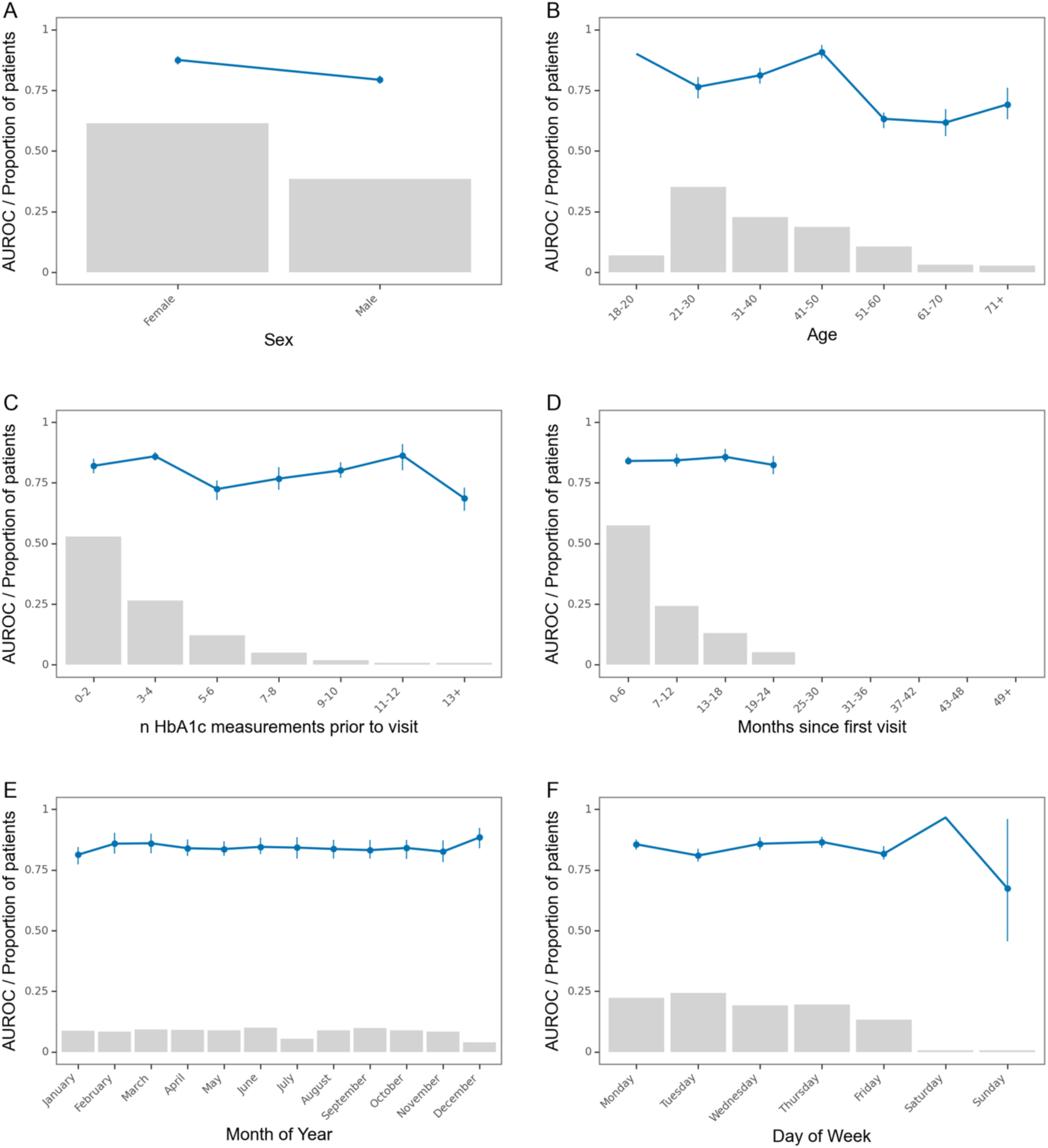
Robustness of the best performing model Robustness of the model across stratifications. Blue line is the area under the receiver operating characteristics curve. Grey bars represent the proportion of prediction times in each bin. Error bars are 95%-confidence intervals from 100-fold bootstrap. Due to the low n in some of the bins, some bootstrap folds contained only one class. This resulted in missing error bars for those bins.

### Post hoc analysis

The performance of XGBoost models using 1, 2, 3 and 4-year lookaheads is available in Supplementary Figure 4-11 and Supplementary Table 5-8. The parsimonious model that was only provided with age, sex and the mean HbA1c within the past 2 years achieved an AUROC of 0.82.

## Discussion

We investigated whether a machine learning model trained on routinely collected electronic health record data can predict development of T2D among patients with mental illness. At the level of service contacts, the best performing model predicted T2D with an AUROC of 0.84, a positive predictive value of 18% and a negative predictive value of 99%. For the patients that developed T2D and which were detected by the model, the median time from first positive prediction to T2D was 2.7 years. A parsimonious model with only sex, age and the mean HbA1c within 2 years achieved an AUROC of 0.82. This highlights the importance of these three features, and the possibility of generalising to healthcare systems with less comprehensive datasets. Both for the main- and the parsimonious model, the obtained level of prediction indicates the potential for implementation.

To the best of our knowledge, this is the first study using machine learning to predict the onset of T2D in patients with mental illness. As such, all comparisons must be made to studies of non-psychiatric populations, which limits comparability. Notable studies include those by Abassi et al.,24 which performed a comprehensive external validation of existing models on a new research dataset; Alghamdi et al.25, which predicted incident diabetes within 5 years based on medical records of cardiovascular fitness; and Cahn et al.8, which trained a prediction model on The Health Improvement Network database from the United Kingdom and tested it on the Israeli Maccabi Health Services dataset. While these studies obtained impressive model performance, they are all either based on research datasets with data that are more comprehensive than what is typically collected as part of routine clinical practice,^25^ or have restricted the study populations to individuals without missing values.^8,24,25^ This limits the generalizability of the findings and substantially complicates clinical implementation. For this reason, in the present study, we deliberately used only routinely collected data on an unrestricted population, ensuring that data can be fed directly from the electronic health record into the model during implementation and that predictions can be generated for all relevant service contacts.

If implemented, the model predictions should be presented to healthcare staff allowing them to initiate intervention at the level of the individual patient. Which interventions to initiate will depend on the situation. Specifically, those identified as being at high risk of T2D are a combination of those who I) have already developed T2D, but are undiagnosed, and II) will develop T2D within the next five years. In either case, the first step will be to test for manifest T2D, for example by an HbA1c measurement. If T2D is indeed present, treatment should follow the available guidelines.^5^ If T2D is not present, the patient remains at high risk, and should be treated as such. In these cases, lifestyle interventions are uncontroversial and cost-effective in both low- and high-income countries across a wide range of cumulative incidence rates^26^, and appear to also be effective in patients with mental illness, e.g., schizophrenia.^27^ In some cases/settings, preventive treatment with metformin may also be an option, although its evidence base is still under debate.^28,29^ In any case, preventive interventions should be tailored to the specific healthcare system/patient population in question, taking the cost-benefit ratio into account. If applied wisely, the preventive potential for such interventions is substantial.^5^

### Limitations

There are limitations to the study, which should be considered by the reader. First, in cohort studies, prevalent cases can be misclassified as being incident, causing a false spike in incidence in the start of the follow-up period. We mitigated this by employing a 2-year wash-in period, which effectively eliminated the false incidence spike (Supplementary Figure 2). Second, prediction models may issue predictions based on clinical suspicion, sometimes referred to as “bias by intensity of monitoring”.^30^ This occurs if healthcare staff changes the monitoring pattern on suspicion of T2D, and this signal may be picked up as a result of the model training. If this is the case, the model will perform poorly when doctors do not suspect T2D, which is when the model is needed the most. We imitated clinical suspicion by only feeding the model information on whether a predictor was measured or not. This model performed poorly (AUROC: 0.60), providing assurance that bias by intensity of monitoring is not severe. Third, when selecting how far a model should look into the future, we are confronted with a trade-off: For prevention, earlier detection is generally more beneficial. However, for the model, the longer into the future we look, the more contacts we must ignore because they do not have sufficient follow-up, thereby decreasing sample size, leading to poorer model performance. To select a reasonable point on this trade-off, we trained models across multiple lookahead windows. For further limitations pertaining to selection of wash-in periods, the definition of T2D, and effects of potential model implementation, see the Supplementary Discussion. Finally, machine learning models vary markedly in their generalisability. We use routinely collected data from a system with universal healthcare. Reusing the unmodified model in different settings would likely yield less optimal prediction. However, the overall approach is likely generalisable and, therefore, retraining the model on data from other settings with the same architecture may allow for transferability.

### Conclusions

The results of this study show that a machine learning model trained on routinely collected data from electronic health records can accurately predict T2D in patients with mental illness. We will, therefore, work towards implementing the model as a clinical decision support tool to facilitate prevention of T2D in this high-risk group.

## Role of the funding source

The study is supported by grants from the Lundbeck Foundation (grant number: R344-2020-1073), the Danish Cancer Society (grant number: R283-A16461), the Central Denmark Region Fund for Strengthening of Health Science (grant number: 1-36-72-4-20) and the Danish Agency for Digitisation Investment Fund for New Technologies (grant number 2020-6720) to Østergaard, who reports further funding from the Lundbeck Foundation (grant number: R358-2020-2341), the Novo Nordisk Foundation (grant number: NNF20SA0062874) and Independent Research Fund Denmark (grant numbers: 7016-00048B and 2096-00055A). The funders played no role in study design, collection, analysis or interpretation of data, the writing of the report or the decision to submit the paper for publication.

## Conflicts of interest

Danielsen has received a speaker honorarium from Otsuka Pharmaceuticals. Østergaard received the 2020 Lundbeck Foundation Young Investigator Prize. Furthermore, Østergaard owns/has owned units of mutual funds with stock tickers DKIGI, IAIMWC and WEKAFKI, and has owned units of exchange traded funds with stock tickers BATE, TRET, QDV5, QDVH, QDVE, SADM, IQQH, USPY, EXH2, 2B76 and EUNL. The remaining authors declare no conflicts of interest.

## Acknowledgements

The authors thank Bettina Nørremark from Aarhus University Hospital – Psychiatry for assistance with extraction of data and Mathias Brønd Sørensen from the Business Intelligence Office, Central Denmark Region, for assistance in building the infrastructure for model training.

## Data availability

According to Danish law, the personally sensitive data used in this study is only available for research projects conducted by employees in the Central Denmark Region following approval from the Legal Office under the Central Denmark Region (in accordance with the Danish Health Care Act §46, Section 2).

## Notes

### Competing Interest Statement

Danielsen has received a speaker honorarium from Otsuka Pharmaceuticals. Oestergaard received the 2020 Lundbeck Foundation Young Investigator Prize. Furthermore, Oestergaard owns/has owned units of mutual funds with stock tickers DKIGI, IAIMWC and WEKAFKI, and has owned units of exchange traded funds with stock tickers BATE, TRET, QDV5, QDVH, QDVE, SADM, IQQH, USPY, EXH2, 2B76 and EUNL. The remaining authors declare no conflicts of interest.

### Author Declarations

Waiver by the The Central Denmark Region Committees on Health Research Ethics, Request 59/2022: According to the Consolidation Act on Research Ethics Review of Health Research Projects, Consolidation Act number 1338 of 1 September 2020, section 14(2) notification of medical database research projects to the research ethics committee system is only required if the project involves human biological material. Therefore your study may be conducted without an approval from the Committees.

