## Supplementary materials for "Development and validation of a machine learning model for prediction of type 2 diabetes in patients with mental illness"

***Supplementary Table 5:*** *Performance by predicted positive rate of XGBoost with 1 years of lookahead.70*

***Supplementary Table 6:*** *Performance by predicted positive rate of XGBoost with 2 years of lookahead.71*

***Supplementary Table 7:*** *Performance by predicted positive rate of XGBoost with 3 years of lookahead.72*

***Supplementary Table 8:*** *Performance by predicted positive rate of XGBoost with 4 years of lookahead.73*

### Supplementary Methods

#### Predictor specification

Supplementary Table 2 has many predictors which did not have values within the lookbehind window. These do not, however, represent missing data in the traditional sense as they do not result from data that were not entered, but from an actual absence of data. This absence reflects clinical practice and should, therefore, be included, as it matches implementation of a potentially predictive model into clinical practice.

#### Predictor flattening

All predictors are time-series. For example, each HbA1c measurement includes both the time it was measured as well as its value. Traditional machine learning models require that each prediction time, e.g., each psychiatric service contact, be represented as one row. However, each HbA1c measurement might be relevant to more than one prediction time. Furthermore, more than one measurement might be relevant to the same prediction time. This means we need a many-to-many join, filter by time, and aggregate within this time filter. Specifically, we looked back a given distance from the prediction time (lookbehind window), and aggregated the measurements within this window (Figure 1F) using the "timeseriesflattener" Python package.[32] The final dataset was quality checked manually by inspecting features, as well as with the Deepchecks suite v0.13.1. It passed all relevant checks, e.g. lack of patient overlap between splits and no predictor being too strongly correlated with the outcome as a sign of leakage.[33]

Hyperparameter tuning

For each lookahead window, the algorithm optimised settings for lookbehind combinations, predictor selection methods, imputation methods, and model hyperparameters (see Supplementary Table 2 for a full description of all settings used for experiments). The search was conducted for 12+ hours on a server with a 30-core Intel Xeon CPU, 512GB of RAM and an NVIDIA A100 GPU, resulting in approximately 250 trained models per lookahead window. Training was terminated when the increase in AUROC over the last 2 hours of training was <0.001. Models were trained using 5-fold cross-validation. The code for the full project is available on Github: https://github.com/Aarhus-Psychiatry-Research/psycop-common/tree/main/src/psycop/projects/t2d

Sensitivity analyses

The following sensitivity analyses were carried out: First, as it could be expected that the T2D pattern would vary across age, sex, and number of prior HbA1c-measurements, we examined predictive performance across these characteristics. Second, we examined the temporal stability of predictions by plotting AUROC across time since first contact to the Psychiatric Services of the Central Denmark Region, as well as examined periodicity across months of the year and days of the week. Third, model performance with only sex, age, and HbA1c as predictors was assessed to estimate the additional value of adding a rich feature set. Fourth, since prediction models may issue predictions based on clinical suspicion, we reran the analysis only providing the model with information on whether a measurement was carried out or not – without providing the value/result of the measurement. This imitates the information about the clinical suspicion that the model is provided with. For instance, if the healthcare staff frequently measures the patient's LDL-level, it likely reflects suspicion of unhealthy lifestyle or predisposition, irrespective of whether LDL is elevated.

### Supplementary Figures and Supplementary Tables

**Supplementary Figure 1.** Selection of eligible prediction times before model selection

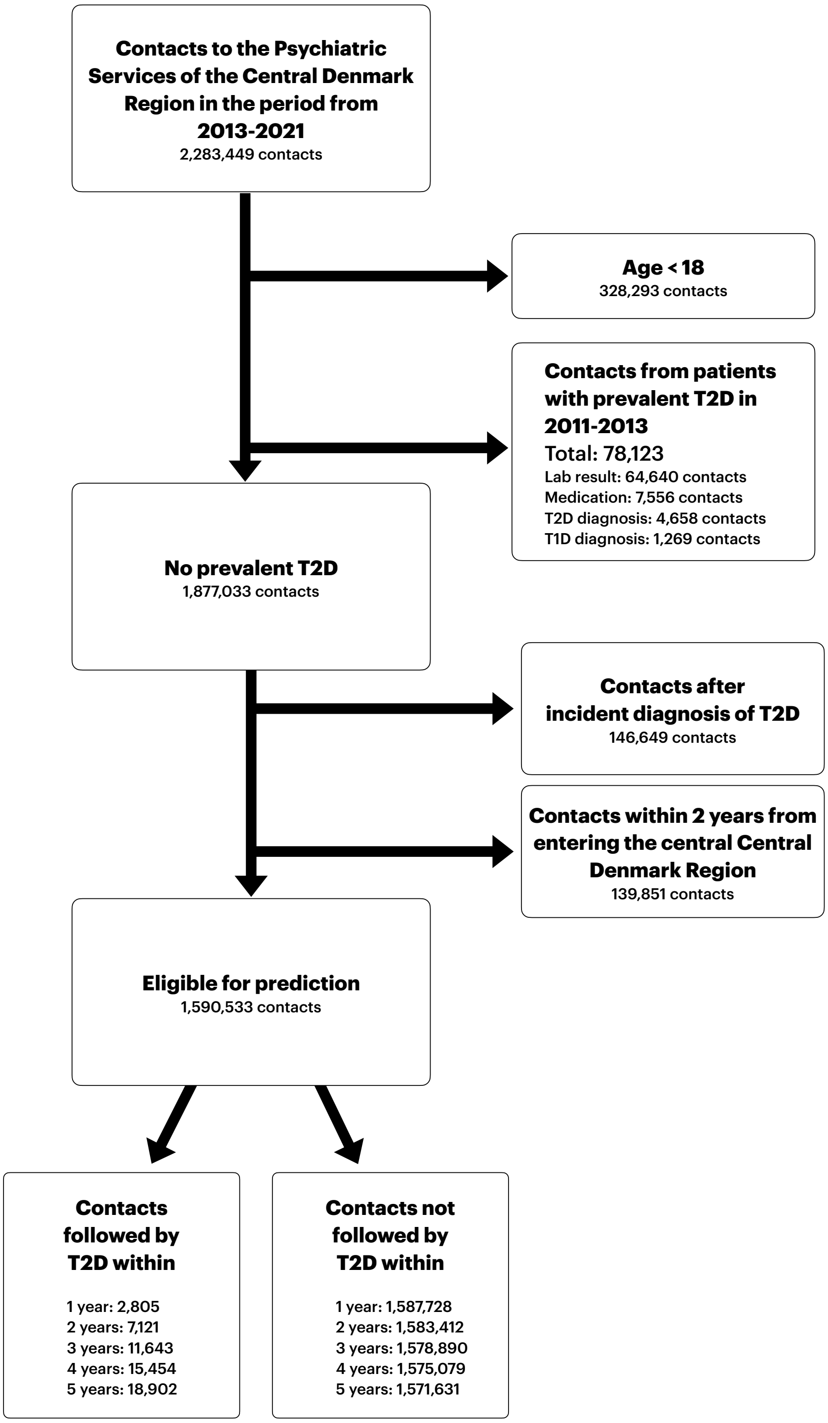

Note that prediction time filtering based on sufficient lookbehind and lookahead (see Figure 1B) happens after the selection illustrated by this figure. As such, counts will differ based on pipeline lookahead and lookbehind. For example, since a total of 8 years of data is available, a model with 1 year of lookbehind and 5 years of lookahead will filter out (5+1)/8 = 75% of the eligible prediction times due to insufficient data.

**Supplementary Figure 2.** First HbA1c ≥ 48 by month for the patients in the dataset

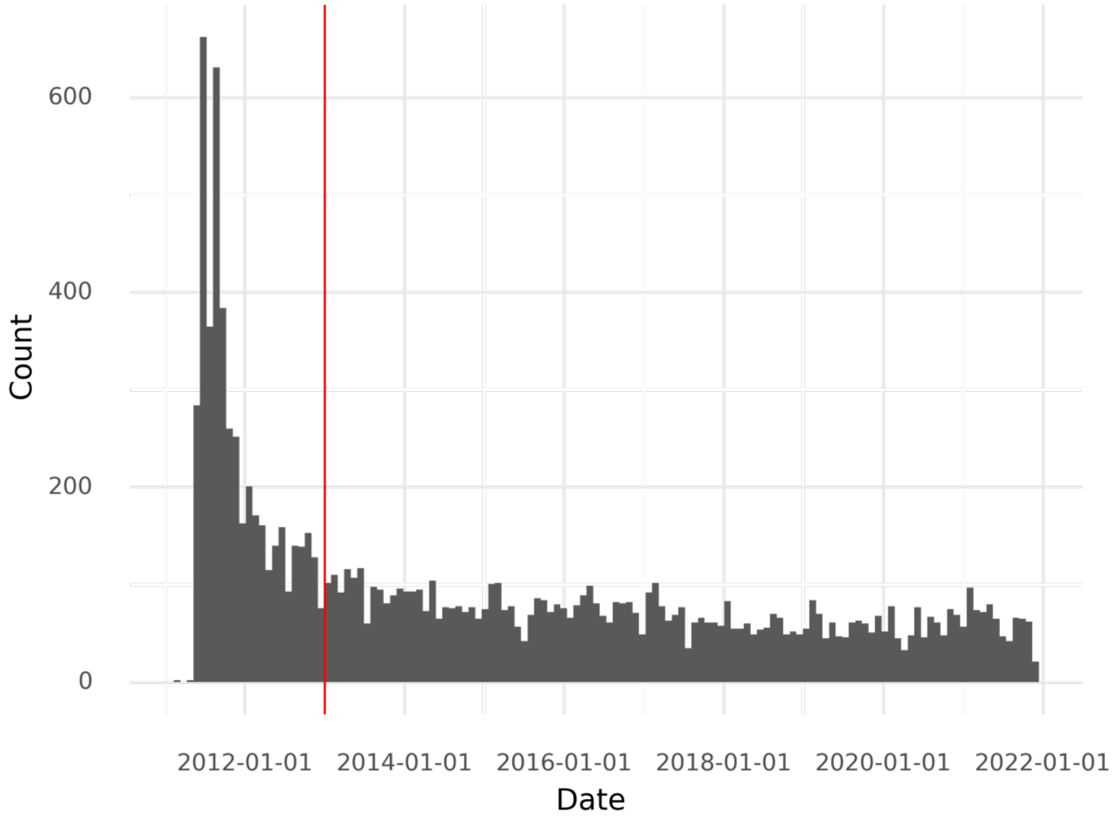

The red line represents the end of the wash-in period (January 1st 2013).

**Supplementary Figure 3:** Cumulative proportion of the last diabetes-measurement within the 5-year lookahead window by classification

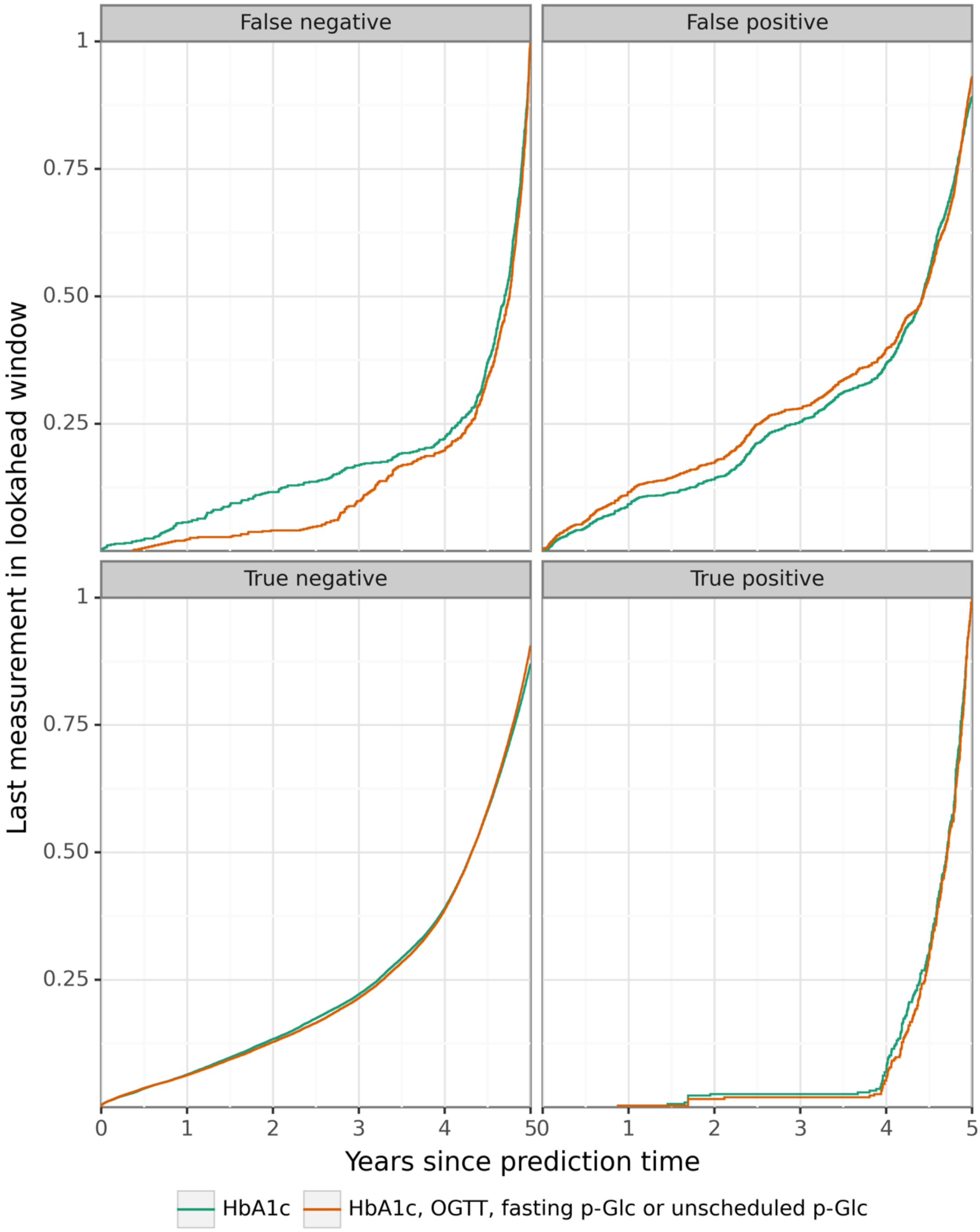

**HbA1c:** Haemoglobin A1c, **OGTT:** Oral glucose tolerance test, **p-Glc:** Plasma glucose.

**Supplementary Figure 4:** Performance of XGBoost at a 3% predicted positive rate with 1 year of lookahead

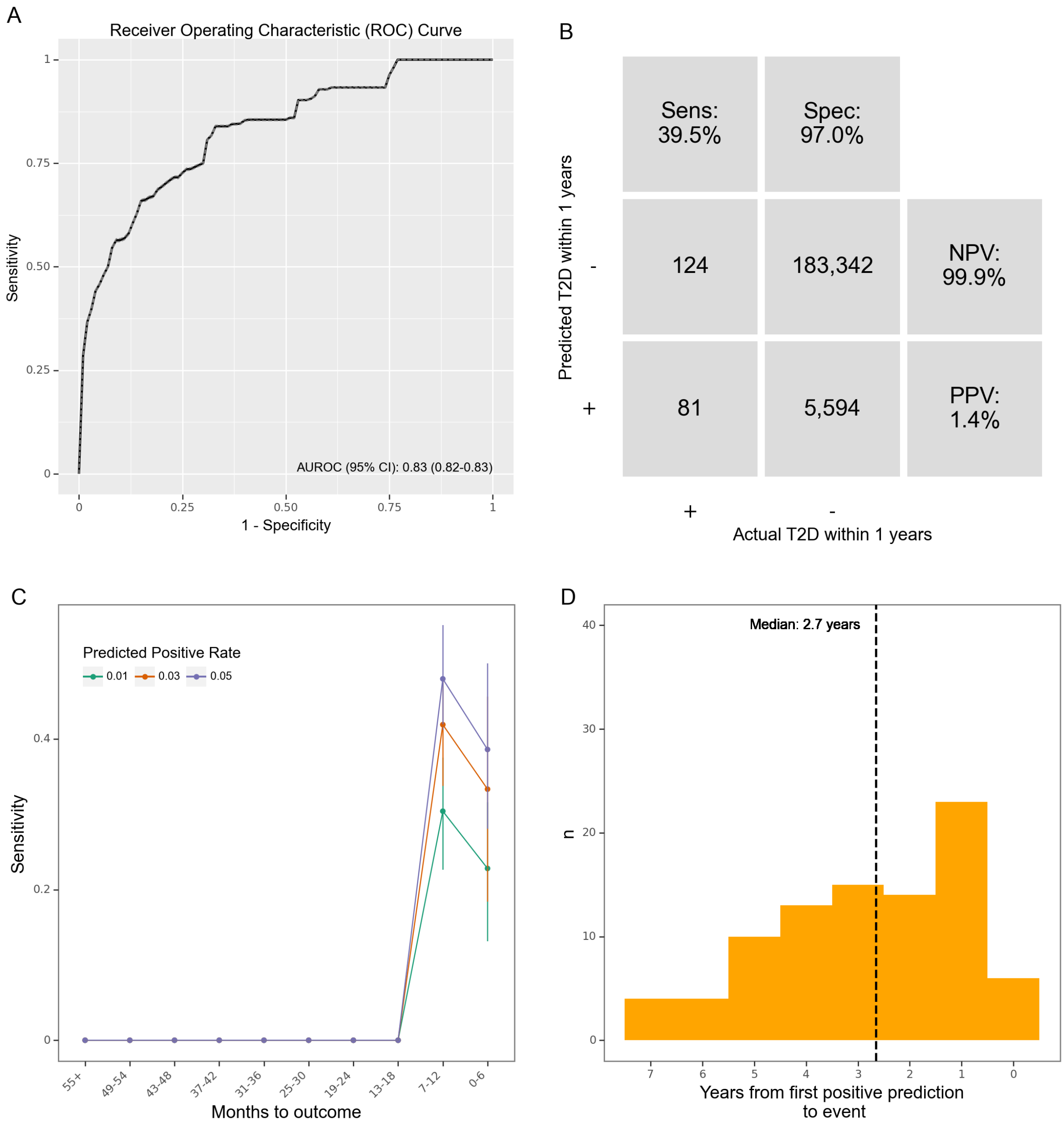

**A**: Receiver operating characteristics (ROC) curve. **B**: Confusion matrix. PPV: Positive predictive value. NPV: Negative predictive value. **C**: Sensitivity by months from prediction time to event, stratified by desired predicted positive rate (PPR). Note that the numbers do not match those in Table 1, since all prediction times with insufficient lookahead distance have been dropped. **D**: Time (years) from the first positive prediction to the patient having developed T2D at a 3% predicted positive rate (PPR).

**Supplementary Figure 5:** Robustness of XGBoost at a 3% predicted positive rate with 1 years of lookahead

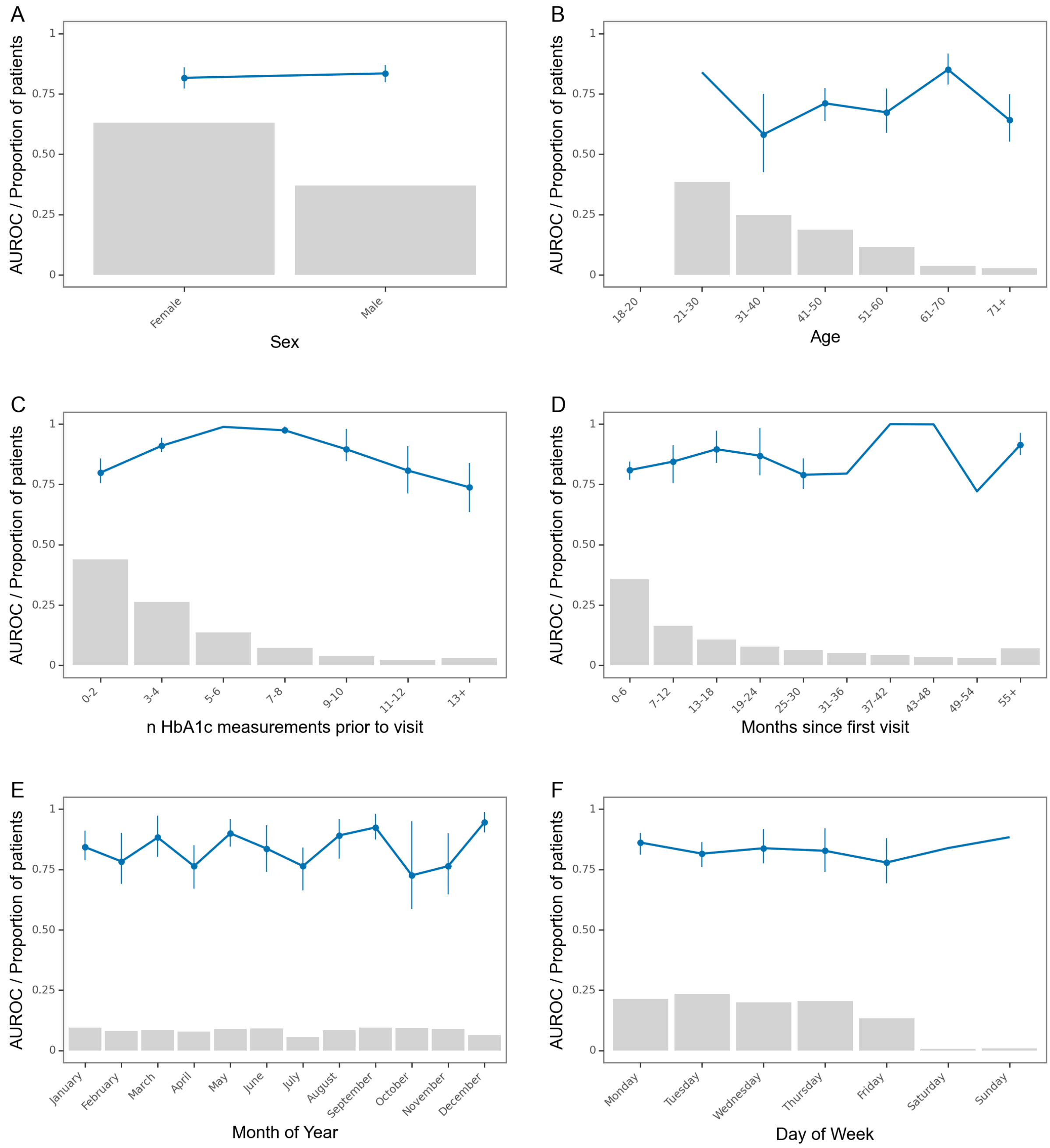

Robustness of the model stratifications. Blue line is the area under the receiver operating characteristics curve. Grey bars represent the proportion of visits that are present in each group. Error bars are 95%-confidence intervals from 100-fold bootstrap.

**Supplementary Figure 6:** Performance of XGBoost at a 3% predicted positive rate with 2 years of lookahead

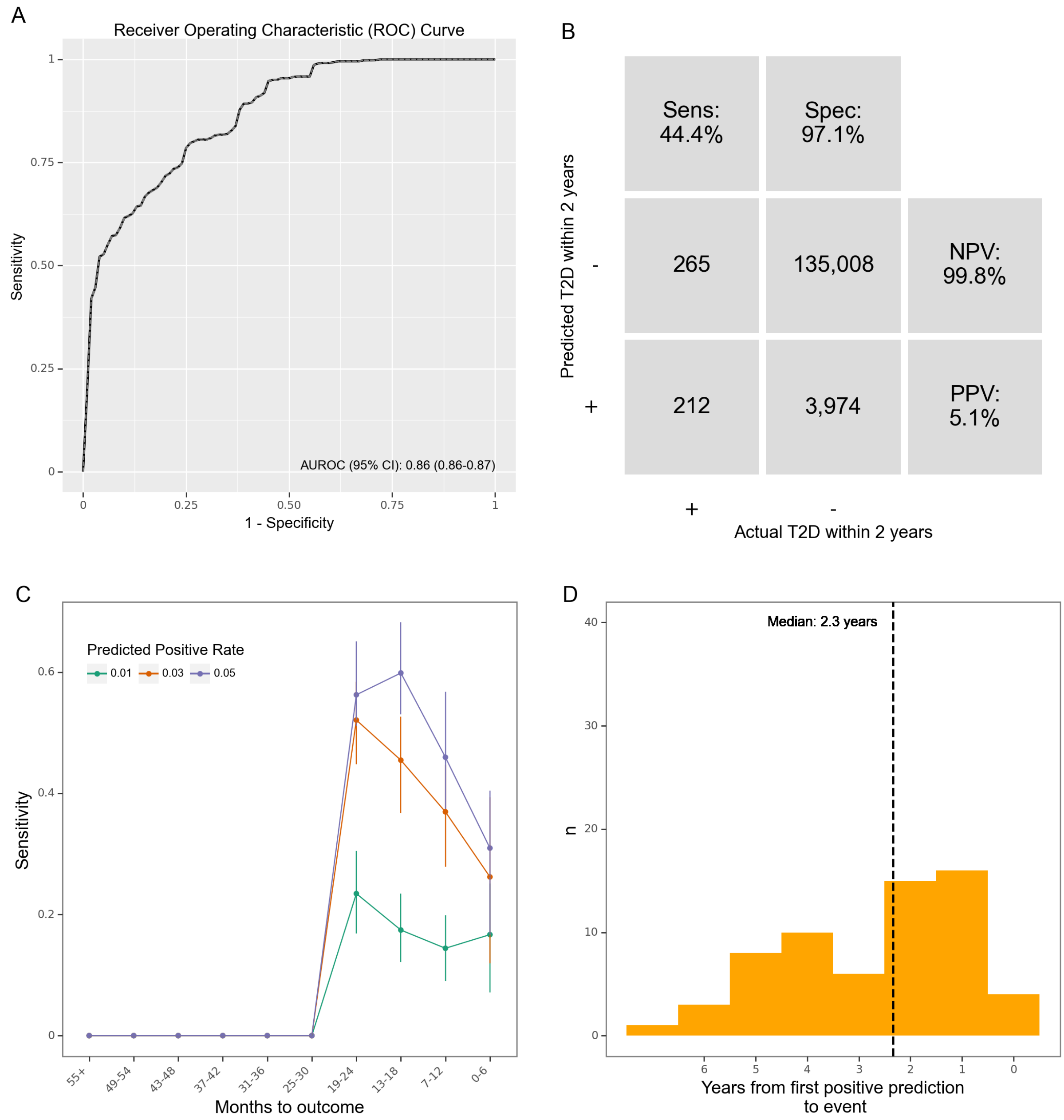

**A**: Receiver operating characteristics (ROC) curve. **B**: Confusion matrix. PPV: Positive predictive value. NPV: Negative predictive value. **C**: Sensitivity by months from prediction time to event, stratified by desired predicted positive rate (PPR). Note that the numbers do not match those in Table 1, since all prediction times with insufficient lookahead distance have been dropped. **D**: Time (years) from the first positive prediction to the patient having developed T2D at a 3% predicted positive rate (PPR).

**Supplementary Figure 7:** Robustness of XGBoost at a 3% predicted positive rate with 2 years of lookahead

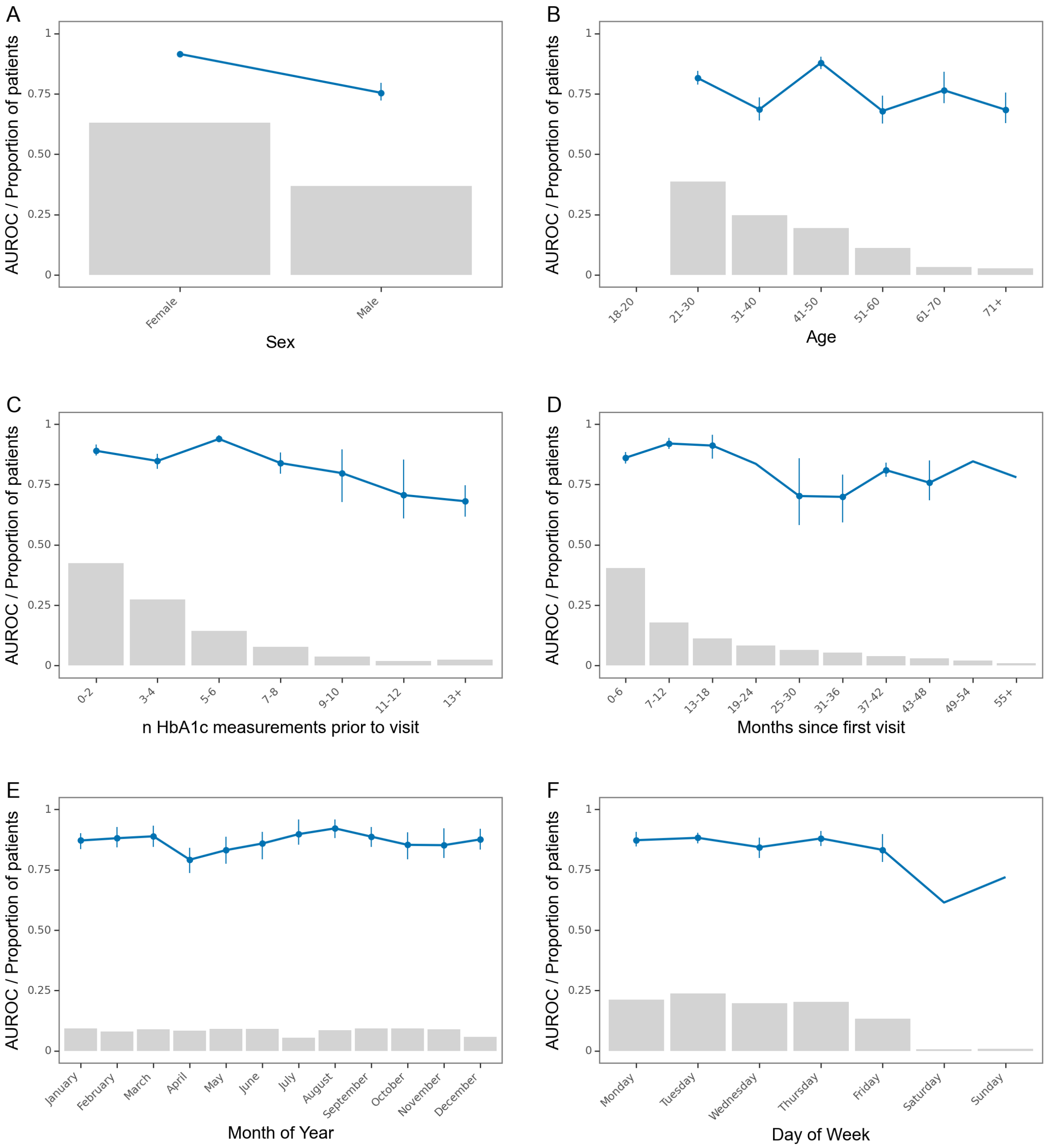

Robustness of the model stratifications. Blue line is the area under the receiver operating characteristics curve. Grey bars represent the proportion of visits that are present in each group. Error bars are 95%-confidence intervals from 100-fold bootstrap.

**Supplementary Figure 8:** Performance of XGBoost at a 3% predicted positive rate with 3 years of lookahead

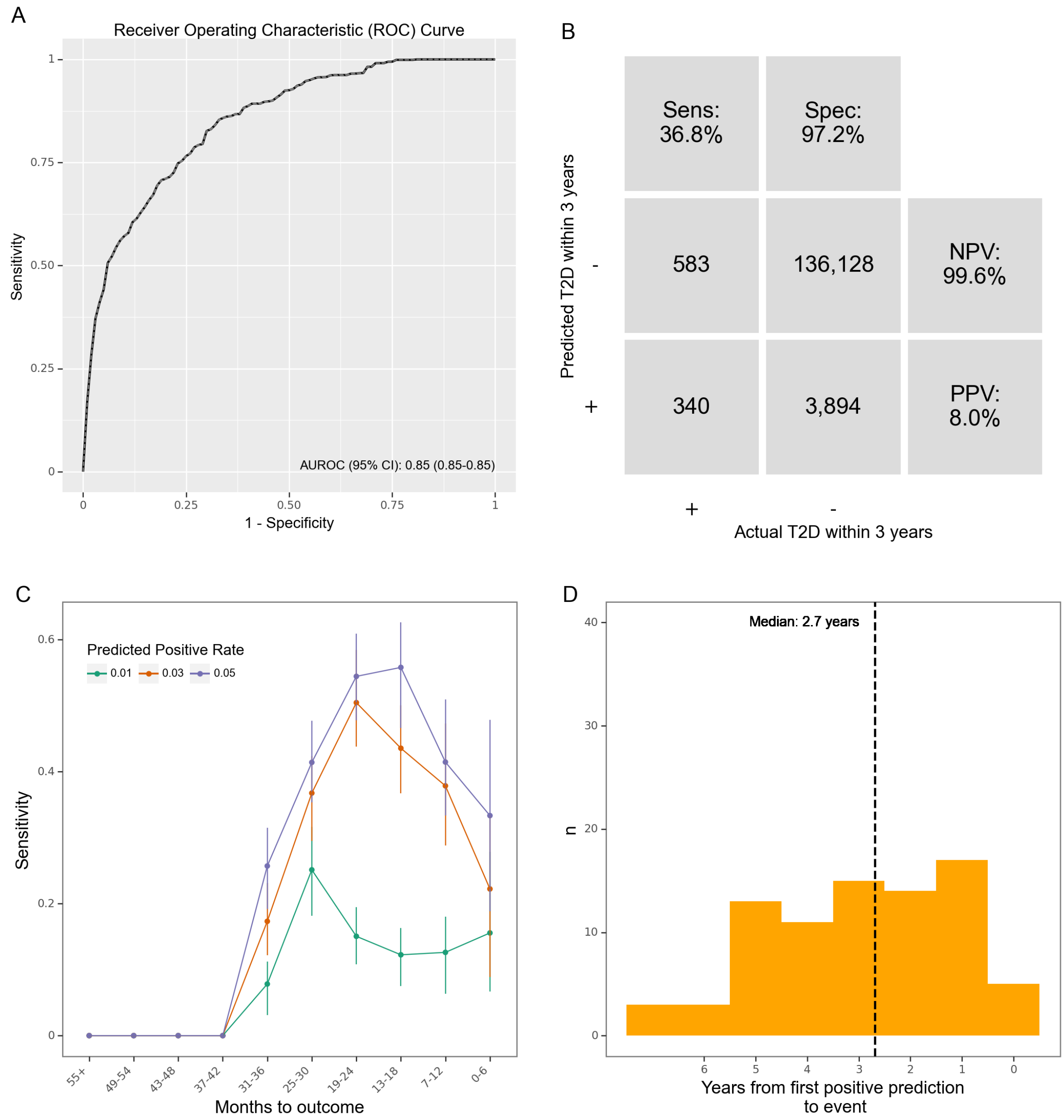

**A**: Receiver operating characteristics (ROC) curve. **B**: Confusion matrix. PPV: Positive predictive value. NPV: Negative predictive value. **C**: Sensitivity by months from prediction time to event, stratified by desired predicted positive rate (PPR). Note that the numbers do not match those in Table 1, since all prediction times with insufficient lookahead distance have been dropped. **D**: Time (years) from the first positive prediction to the patient having developed T2D at a 3% predicted positive rate (PPR).

**Supplementary Figure 9:** Robustness of XGBoost at a 3% predicted positive rate with 3 years of lookahead

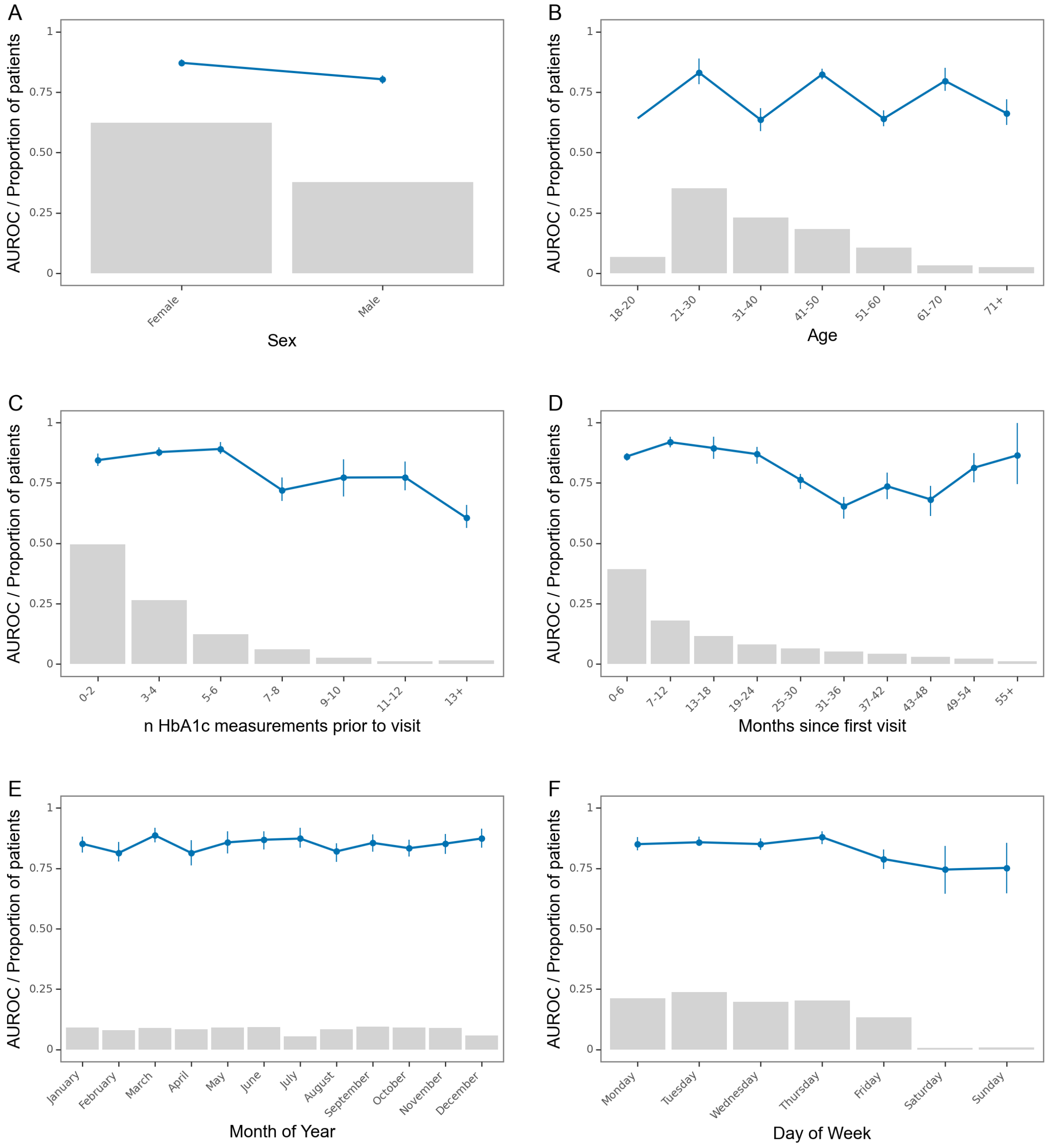

Robustness of the model stratifications. Blue line is the area under the receiver operating characteristics curve. Grey bars represent the proportion of visits that are present in each group. Error bars are 95%-confidence intervals from 100-fold bootstrap.

**Supplementary Figure 10:** Performance of XGBoost at a 3% predicted positive rate with 4 years of lookahead

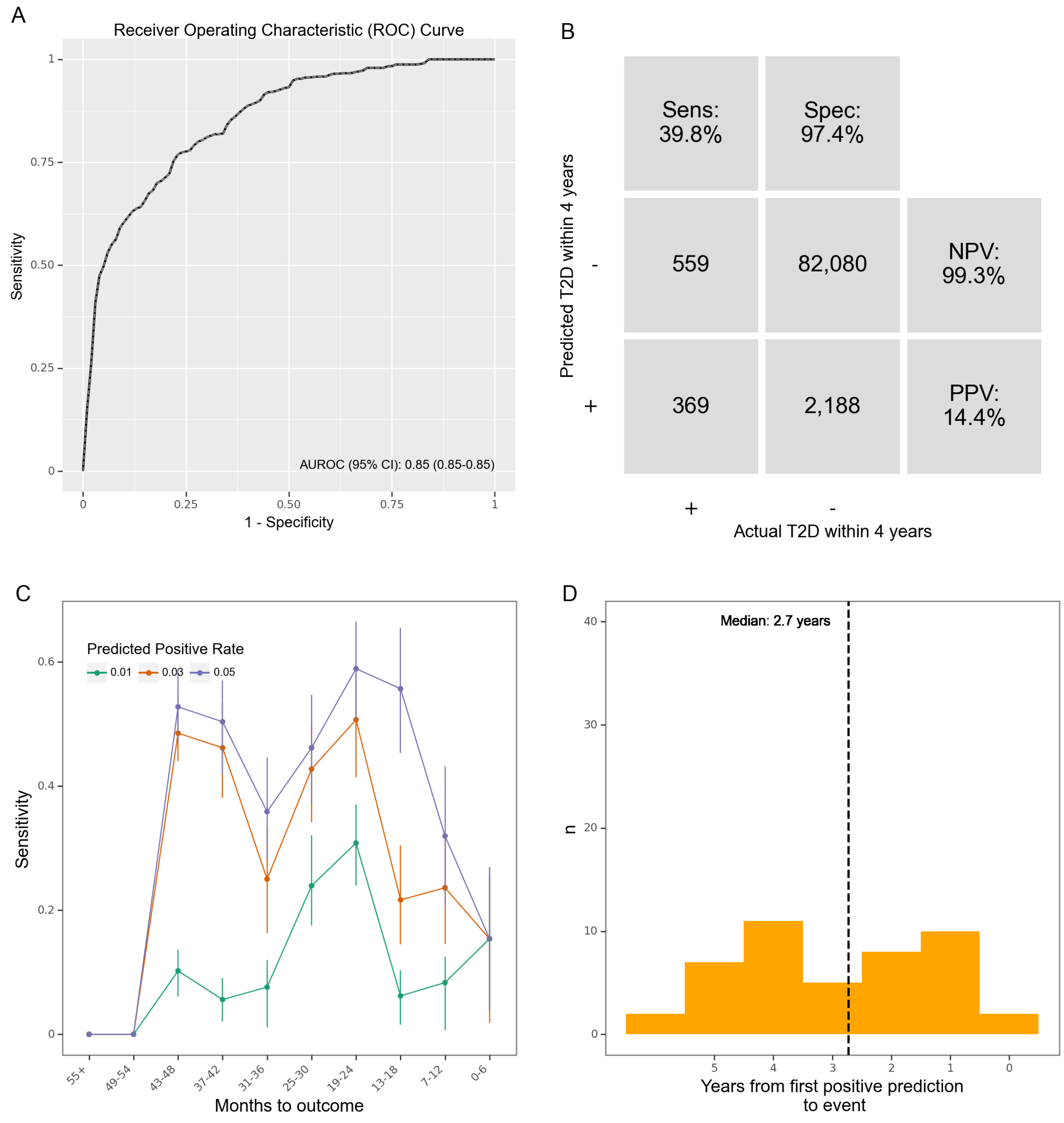

**A**: Receiver operating characteristics (ROC) curve. **B**: Confusion matrix. PPV: Positive predictive value. NPV: Negative predictive value. **C**: Sensitivity by months from prediction time to event, stratified by desired predicted positive rate (PPR). Note that the numbers do not match those in Table 1, since all prediction times with insufficient lookahead distance have been dropped. **D**: Time (years) from the first positive prediction to the patient having developed T2D at a 3% predicted positive rate (PPR).

**Supplementary Figure 11:** Robustness of XGBoost at a 3% predicted positive rate with 4 years of lookahead.

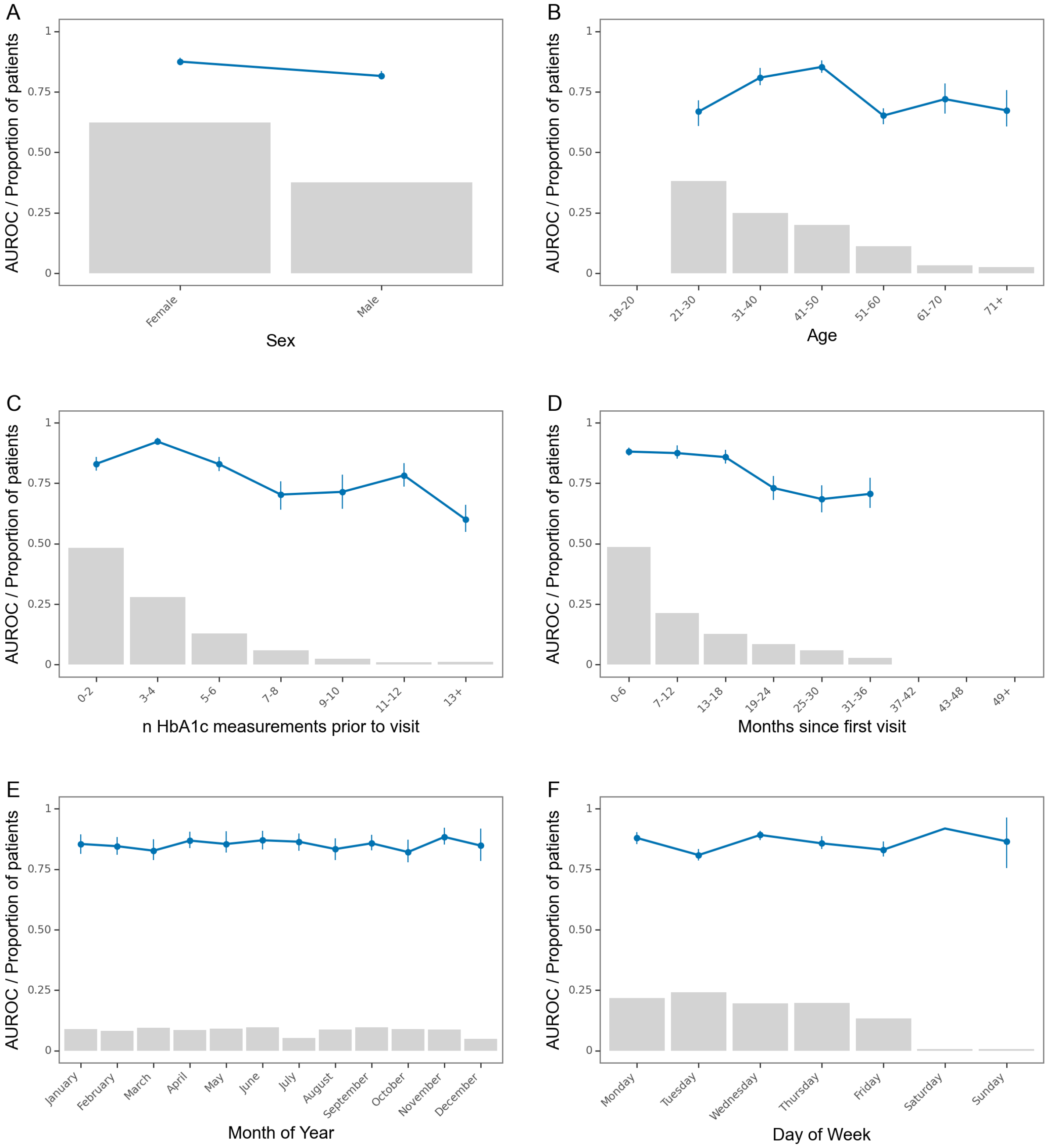

Robustness of the model stratifications. Blue line is the area under the receiver operating characteristics curve. Grey bars represent the proportion of visits that are present in each group. Error bars are 95%-confidence intervals from 100-fold bootstrap

**Supplementary Table 2:** Descriptive statistics for predictors (a list of abbreviations is inserted below the table)

| Predictor | Lookbehind days | Aggregation function | Unique values in predictor | Fallback strategy | Mean | Proportion using fallback | 25-percentile | 50-percentile | 75-percentile |
|---|---|---|---|---|---|---|---|---|---|
| alat | 30 | maximum | 510 | nan | 32.29 | 0.82 | 15 | 21 | 32 |
| alat | 30 | minimum | 379 | nan | 27.17 | 0.82 | 14 | 20 | 31 |
| alat | 1095 | minimum | 231 | nan | 18.73 | 0.07 | 11 | 15 | 22 |
| alat | 730 | minimum | 253 | nan | 19.93 | 0.1 | 12 | 16 | 23 |
| alat | 365 | minimum | 290 | nan | 22.06 | 0.21 | 13 | 18 | 26 |
| alat | 180 | minimum | 327 | nan | 23.84 | 0.39 | 14 | 19 | 28 |
| alat | 1095 | maximum | 765 | nan | 48.7 | 0.07 | 19 | 27 | 44 |
| alat | 1825 | maximum | 792 | nan | 54.01 | 0.06 | 20 | 29 | 48 |
| alat | 730 | maximum | 735 | nan | 44.42 | 0.1 | 18 | 26 | 41 |
| alat | 365 | maximum | 681 | nan | 38.55 | 0.21 | 16 | 23 | 37 |
| alat | 180 | maximum | 614 | nan | 35.18 | 0.39 | 16 | 22 | 34 |
| alat | 1825 | minimum | 218 | nan | 17.6 | 0.06 | 11 | 15 | 21 |
| alat | 1460 | minimum | 223 | nan | 18.04 | 0.06 | 11 | 15 | 21 |
| alat | 1825 | mean | 24744 | nan | 28.96 | 0.06 | 15.5 | 21.2 | 31 |
| alat | 1460 | maximum | 780 | nan | 51.66 | 0.06 | 19 | 28 | 47 |
| alat | 180 | mean | 6445 | nan | 28.2 | 0.39 | 15 | 21 | 31 |
| alat | 30 | mean | 1940 | nan | 29.42 | 0.82 | 15 | 21 | 31.5 |
| alat | 1825 | latest | 467 | nan | 25.96 | 0.06 | 14 | 20 | 30 |
| alat | 1460 | latest | 467 | nan | 25.97 | 0.06 | 14 | 20 | 30 |
| alat | 1095 | latest | 467 | nan | 26 | 0.07 | 14 | 20 | 30 |
| alat | 365 | latest | 464 | nan | 26.26 | 0.21 | 14 | 20 | 30 |
| alat | 180 | latest | 459 | nan | 26.6 | 0.39 | 15 | 20 | 31 |
| alat | 730 | latest | 465 | nan | 26.07 | 0.1 | 14 | 20 | 30 |

| | | | | | | | | | |
|---|---|---|---|---|---|---|---|---|---|
| alat | 1460 | mean | 23198 | nan | 28.85 | 0.06 | 15.5 | 21.1 | 31 |
| alat | 1095 | mean | 20409 | nan | 28.69 | 0.07 | 15.3 | 21 | 31 |
| alat | 730 | mean | 16304 | nan | 28.51 | 0.1 | 15 | 21 | 31 |
| alat | 365 | mean | 10417 | nan | 28.19 | 0.21 | 15 | 21 | 31 |
| alat | 30 | latest | 439 | nan | 28.26 | 0.82 | 15 | 21 | 31 |
| albumine_creatinine_ratio | 180 | maximum | 410 | nan | 77.18 | 0.98 | 3.4 | 7 | 14 |
| albumine_creatinine_ratio | 1460 | maximum | 493 | nan | 58.96 | 0.93 | 4 | 7 | 15 |
| albumine_creatinine_ratio | 365 | maximum | 439 | nan | 64.57 | 0.97 | 3.4 | 6.7 | 14 |
| albumine_creatinine_ratio | 730 | maximum | 464 | nan | 60.29 | 0.95 | 4 | 7 | 14 |
| albumine_creatinine_ratio | 1825 | minimum | 331 | nan | 18.67 | 0.93 | 3 | 5 | 9.4 |
| albumine_creatinine_ratio | 1095 | maximum | 481 | nan | 57.99 | 0.94 | 4 | 7 | 14.1 |
| albumine_creatinine_ratio | 1825 | maximum | 497 | nan | 58.02 | 0.93 | 4 | 7 | 15.4 |
| albumine_creatinine_ratio | 180 | minimum | 375 | nan | 46.1 | 0.98 | 3.4 | 6.7 | 13 |
| albumine_creatinine_ratio | 365 | minimum | 382 | nan | 30.51 | 0.97 | 3 | 6 | 12 |
| albumine_creatinine_ratio | 730 | minimum | 366 | nan | 24.25 | 0.95 | 3 | 5 | 11 |
| albumine_creatinine_ratio | 1095 | minimum | 343 | nan | 20.5 | 0.94 | 3 | 5 | 10 |
| albumine_creatinine_ratio | 30 | maximum | 296 | nan | 93.35 | 1 | 4 | 7 | 16 |
| albumine_creatinine_ratio | 30 | minimum | 291 | nan | 87.91 | 1 | 4 | 7 | 16 |
| albumine_creatinine_ratio | 1460 | minimum | 339 | nan | 19.56 | 0.93 | 3 | 5 | 10 |
| albumine_creatinine_ratio | 1825 | latest | 498 | nan | 31.45 | 0.93 | 3 | 6 | 12 |
| albumine_creatinine_ratio | 180 | latest | 414 | nan | 56.63 | 0.98 | 3.4 | 6.7 | 14 |
| albumine_creatinine_ratio | 30 | mean | 315 | nan | 90.72 | 1 | 4 | 7 | 16 |
| albumine_creatinine_ratio | 1825 | mean | 2100 | nan | 33.32 | 0.93 | 3.5 | 6 | 13 |
| albumine_creatinine_ratio | 730 | mean | 1404 | nan | 38.51 | 0.95 | 3.4 | 6 | 12.7 |
| albumine_creatinine_ratio | 1095 | mean | 1752 | nan | 35.2 | 0.94 | 3.4 | 6 | 12.5 |
| albumine_creatinine_ratio | 1460 | mean | 1945 | nan | 34.57 | 0.93 | 3.5 | 6 | 13 |
| albumine_creatinine_ratio | 30 | latest | 294 | nan | 88.13 | 1 | 4 | 7 | 16 |
| albumine_creatinine_ratio | 365 | mean | 971 | nan | 45.09 | 0.97 | 3.4 | 6 | 13 |

| | | | | | | | | | |
|---|---|---|---|---|---|---|---|---|---|
| albumine_creatinine_ratio | 365 | latest | 459 | nan | 42.56 | 0.97 | 3.4 | 6 | 12 |
| albumine_creatinine_ratio | 730 | latest | 478 | nan | 36.07 | 0.95 | 3 | 6 | 12 |
| albumine_creatinine_ratio | 1095 | latest | 489 | nan | 33.44 | 0.94 | 3 | 6 | 12 |
| albumine_creatinine_ratio | 1460 | latest | 497 | nan | 32.5 | 0.93 | 3 | 6 | 12 |
| albumine_creatinine_ratio | 180 | mean | 707 | nan | 59.86 | 0.98 | 3.4 | 6.7 | 14 |
| antihypertensives | 180 | mean | 2 | 0 | 0 | 1 | 0 | 0 | 0 |
| antihypertensives | 30 | mean | 2 | 0 | 0 | 1 | 0 | 0 | 0 |
| antihypertensives | 1460 | minimum | 2 | 0 | 0.01 | 0.99 | 0 | 0 | 0 |
| antihypertensives | 1095 | minimum | 2 | 0 | 0.01 | 0.99 | 0 | 0 | 0 |
| antihypertensives | 365 | minimum | 2 | 0 | 0 | 1 | 0 | 0 | 0 |
| antihypertensives | 1460 | mean | 2 | 0 | 0.01 | 0.99 | 0 | 0 | 0 |
| antihypertensives | 30 | minimum | 2 | 0 | 0 | 1 | 0 | 0 | 0 |
| antihypertensives | 365 | mean | 2 | 0 | 0 | 1 | 0 | 0 | 0 |
| antihypertensives | 30 | maximum | 2 | 0 | 0 | 1 | 0 | 0 | 0 |
| antihypertensives | 1460 | maximum | 2 | 0 | 0.01 | 0.99 | 0 | 0 | 0 |
| antihypertensives | 1825 | maximum | 2 | 0 | 0.01 | 0.99 | 0 | 0 | 0 |
| antihypertensives | 180 | minimum | 2 | 0 | 0 | 1 | 0 | 0 | 0 |
| antihypertensives | 730 | mean | 2 | 0 | 0.01 | 0.99 | 0 | 0 | 0 |
| antihypertensives | 730 | minimum | 2 | 0 | 0.01 | 0.99 | 0 | 0 | 0 |
| antihypertensives | 1825 | minimum | 2 | 0 | 0.01 | 0.99 | 0 | 0 | 0 |
| antihypertensives | 365 | latest | 2 | 0 | 0 | 1 | 0 | 0 | 0 |
| antihypertensives | 730 | latest | 2 | 0 | 0.01 | 0.99 | 0 | 0 | 0 |
| antihypertensives | 1095 | latest | 2 | 0 | 0.01 | 0.99 | 0 | 0 | 0 |
| antihypertensives | 180 | maximum | 2 | 0 | 0 | 1 | 0 | 0 | 0 |
| antihypertensives | 1825 | latest | 2 | 0 | 0.01 | 0.99 | 0 | 0 | 0 |
| antihypertensives | 1095 | maximum | 2 | 0 | 0.01 | 0.99 | 0 | 0 | 0 |
| antihypertensives | 180 | latest | 2 | 0 | 0 | 1 | 0 | 0 | 0 |
| antihypertensives | 730 | maximum | 2 | 0 | 0.01 | 0.99 | 0 | 0 | 0 |

| | | | | | | | | | |
|---|---|---|---|---|---|---|---|---|---|
| antihypertensives | 365 | maximum | 2 | 0 | 0 | 1 | 0 | 0 | 0 |
| antihypertensives | 30 | latest | 2 | 0 | 0 | 1 | 0 | 0 | 0 |
| antihypertensives | 1460 | latest | 2 | 0 | 0.01 | 0.99 | 0 | 0 | 0 |
| antihypertensives | 1825 | mean | 2 | 0 | 0.01 | 0.99 | 0 | 0 | 0 |
| antihypertensives | 1095 | mean | 2 | 0 | 0.01 | 0.99 | 0 | 0 | 0 |
| antipsychotics | 1095 | maximum | 2 | 0 | 0.3 | 0.7 | 0 | 0 | 1 |
| antipsychotics | 730 | maximum | 2 | 0 | 0.27 | 0.73 | 0 | 0 | 1 |
| antipsychotics | 365 | maximum | 2 | 0 | 0.22 | 0.78 | 0 | 0 | 0 |
| antipsychotics | 180 | maximum | 2 | 0 | 0.17 | 0.83 | 0 | 0 | 0 |
| antipsychotics | 30 | maximum | 2 | 0 | 0.08 | 0.92 | 0 | 0 | 0 |
| antipsychotics | 1825 | minimum | 2 | 0 | 0.32 | 0.68 | 0 | 0 | 1 |
| antipsychotics | 1460 | latest | 2 | 0 | 0.31 | 0.69 | 0 | 0 | 1 |
| antipsychotics | 1095 | latest | 2 | 0 | 0.3 | 0.7 | 0 | 0 | 1 |
| antipsychotics | 730 | latest | 2 | 0 | 0.27 | 0.73 | 0 | 0 | 1 |
| antipsychotics | 365 | latest | 2 | 0 | 0.22 | 0.78 | 0 | 0 | 0 |
| antipsychotics | 1460 | maximum | 2 | 0 | 0.31 | 0.69 | 0 | 0 | 1 |
| antipsychotics | 30 | latest | 2 | 0 | 0.08 | 0.92 | 0 | 0 | 0 |
| antipsychotics | 1825 | maximum | 2 | 0 | 0.32 | 0.68 | 0 | 0 | 1 |
| antipsychotics | 180 | latest | 2 | 0 | 0.17 | 0.83 | 0 | 0 | 0 |
| antipsychotics | 180 | minimum | 2 | 0 | 0.17 | 0.83 | 0 | 0 | 0 |
| antipsychotics | 1825 | latest | 2 | 0 | 0.32 | 0.68 | 0 | 0 | 1 |
| antipsychotics | 1825 | mean | 2 | 0 | 0.32 | 0.68 | 0 | 0 | 1 |
| antipsychotics | 1460 | mean | 2 | 0 | 0.31 | 0.69 | 0 | 0 | 1 |
| antipsychotics | 1095 | mean | 2 | 0 | 0.3 | 0.7 | 0 | 0 | 1 |
| antipsychotics | 730 | mean | 2 | 0 | 0.27 | 0.73 | 0 | 0 | 1 |
| antipsychotics | 365 | mean | 2 | 0 | 0.22 | 0.78 | 0 | 0 | 0 |
| antipsychotics | 30 | minimum | 2 | 0 | 0.08 | 0.92 | 0 | 0 | 0 |
| antipsychotics | 30 | mean | 2 | 0 | 0.08 | 0.92 | 0 | 0 | 0 |

| | | | | | | | | | |
|---|---|---|---|---|---|---|---|---|---|
| antipsychotics | 1460 | minimum | 2 | 0 | 0.31 | 0.69 | 0 | 0 | 1 |
| antipsychotics | 1095 | minimum | 2 | 0 | 0.3 | 0.7 | 0 | 0 | 1 |
| antipsychotics | 730 | minimum | 2 | 0 | 0.27 | 0.73 | 0 | 0 | 1 |
| antipsychotics | 365 | minimum | 2 | 0 | 0.22 | 0.78 | 0 | 0 | 0 |
| antipsychotics | 180 | mean | 2 | 0 | 0.17 | 0.83 | 0 | 0 | 0 |
| arterial_p_glc | 1460 | minimum | 119 | nan | 5.88 | 0.9 | 5.1 | 5.6 | 6.4 |
| arterial_p_glc | 1095 | minimum | 116 | nan | 5.91 | 0.91 | 5.1 | 5.6 | 6.5 |
| arterial_p_glc | 730 | minimum | 118 | nan | 5.94 | 0.93 | 5.1 | 5.7 | 6.5 |
| arterial_p_glc | 365 | minimum | 114 | nan | 5.97 | 0.95 | 5.2 | 5.7 | 6.6 |
| arterial_p_glc | 30 | maximum | 136 | nan | 6.8 | 0.99 | 5.5 | 6.3 | 7.5 |
| arterial_p_glc | 180 | maximum | 162 | nan | 7.02 | 0.97 | 5.6 | 6.5 | 7.8 |
| arterial_p_glc | 180 | minimum | 111 | nan | 5.98 | 0.97 | 5.2 | 5.7 | 6.6 |
| arterial_p_glc | 730 | maximum | 178 | nan | 7.19 | 0.93 | 5.7 | 6.7 | 8.1 |
| arterial_p_glc | 1460 | maximum | 179 | nan | 7.26 | 0.9 | 5.8 | 6.7 | 8.2 |
| arterial_p_glc | 730 | mean | 1625 | nan | 6.47 | 0.93 | 5.6 | 6.2 | 7.2 |
| arterial_p_glc | 1095 | maximum | 179 | nan | 7.24 | 0.91 | 5.7 | 6.7 | 8.1 |
| arterial_p_glc | 1825 | minimum | 120 | nan | 5.87 | 0.89 | 5.1 | 5.6 | 6.4 |
| arterial_p_glc | 365 | maximum | 172 | nan | 7.12 | 0.95 | 5.7 | 6.6 | 7.9 |
| arterial_p_glc | 365 | mean | 1340 | nan | 6.46 | 0.95 | 5.5 | 6.2 | 7.2 |
| arterial_p_glc | 30 | mean | 664 | nan | 6.36 | 0.99 | 5.4 | 6.1 | 7 |
| arterial_p_glc | 1095 | mean | 1772 | nan | 6.47 | 0.91 | 5.6 | 6.2 | 7.1 |
| arterial_p_glc | 180 | mean | 1068 | nan | 6.43 | 0.97 | 5.5 | 6.2 | 7.1 |
| arterial_p_glc | 1825 | maximum | 179 | nan | 7.28 | 0.89 | 5.8 | 6.7 | 8.2 |
| arterial_p_glc | 1825 | latest | 128 | nan | 6.43 | 0.89 | 5.4 | 6.1 | 7.1 |
| arterial_p_glc | 1460 | latest | 128 | nan | 6.42 | 0.9 | 5.4 | 6.1 | 7.1 |
| arterial_p_glc | 730 | latest | 127 | nan | 6.42 | 0.93 | 5.4 | 6.1 | 7.1 |
| arterial_p_glc | 1095 | latest | 127 | nan | 6.43 | 0.91 | 5.4 | 6.1 | 7.1 |
| arterial_p_glc | 180 | latest | 118 | nan | 6.38 | 0.97 | 5.4 | 6.1 | 7 |

| | | | | | | | | | |
|---|---|---|---|---|---|---|---|---|---|
| arterial_p_glc | 30 | latest | 108 | nan | 6.31 | 0.99 | 5.4 | 6 | 6.9 |
| arterial_p_glc | 1825 | mean | 1914 | nan | 6.47 | 0.89 | 5.6 | 6.2 | 7.1 |
| arterial_p_glc | 1460 | mean | 1865 | nan | 6.46 | 0.9 | 5.6 | 6.2 | 7.1 |
| arterial_p_glc | 30 | minimum | 104 | nan | 6.02 | 0.99 | 5.2 | 5.8 | 6.6 |
| arterial_p_glc | 365 | latest | 124 | nan | 6.41 | 0.95 | 5.4 | 6.1 | 7.1 |
| benzodiazepine_related_sleeping_agents | 730 | maximum | 2 | 0 | 0.17 | 0.83 | 0 | 0 | 0 |
| benzodiazepine_related_sleeping_agents | 30 | maximum | 2 | 0 | 0.03 | 0.97 | 0 | 0 | 0 |
| benzodiazepine_related_sleeping_agents | 180 | maximum | 2 | 0 | 0.09 | 0.91 | 0 | 0 | 0 |
| benzodiazepine_related_sleeping_agents | 365 | maximum | 2 | 0 | 0.13 | 0.87 | 0 | 0 | 0 |
| benzodiazepine_related_sleeping_agents | 1095 | maximum | 2 | 0 | 0.2 | 0.8 | 0 | 0 | 0 |
| benzodiazepine_related_sleeping_agents | 1095 | minimum | 2 | 0 | 0.2 | 0.8 | 0 | 0 | 0 |
| benzodiazepine_related_sleeping_agents | 1460 | maximum | 2 | 0 | 0.21 | 0.79 | 0 | 0 | 0 |
| benzodiazepine_related_sleeping_agents | 180 | minimum | 2 | 0 | 0.09 | 0.91 | 0 | 0 | 0 |
| benzodiazepine_related_sleeping_agents | 365 | minimum | 2 | 0 | 0.13 | 0.87 | 0 | 0 | 0 |
| benzodiazepine_related_sleeping_agents | 730 | minimum | 2 | 0 | 0.17 | 0.83 | 0 | 0 | 0 |
| benzodiazepine_related_sleeping_agents | 1825 | maximum | 2 | 0 | 0.22 | 0.78 | 0 | 0 | 0 |
| benzodiazepine_related_sleeping_agents | 1460 | minimum | 2 | 0 | 0.21 | 0.79 | 0 | 0 | 0 |
| benzodiazepine_related_sleeping_agents | 180 | mean | 2 | 0 | 0.09 | 0.91 | 0 | 0 | 0 |

| | | | | | | | | | |
|---|---|---|---|---|---|---|---|---|---|
| benzodiazepine_related_sleeping_agents | 30 | minimum | 2 | 0 | 0.03 | 0.97 | 0 | 0 | 0 |
| benzodiazepine_related_sleeping_agents | 30 | mean | 2 | 0 | 0.03 | 0.97 | 0 | 0 | 0 |
| benzodiazepine_related_sleeping_agents | 1825 | latest | 2 | 0 | 0.22 | 0.78 | 0 | 0 | 0 |
| benzodiazepine_related_sleeping_agents | 1460 | latest | 2 | 0 | 0.21 | 0.79 | 0 | 0 | 0 |
| benzodiazepine_related_sleeping_agents | 1095 | latest | 2 | 0 | 0.2 | 0.8 | 0 | 0 | 0 |
| benzodiazepine_related_sleeping_agents | 1825 | minimum | 2 | 0 | 0.22 | 0.78 | 0 | 0 | 0 |
| benzodiazepine_related_sleeping_agents | 730 | latest | 2 | 0 | 0.17 | 0.83 | 0 | 0 | 0 |
| benzodiazepine_related_sleeping_agents | 365 | latest | 2 | 0 | 0.13 | 0.87 | 0 | 0 | 0 |
| benzodiazepine_related_sleeping_agents | 1460 | mean | 2 | 0 | 0.21 | 0.79 | 0 | 0 | 0 |
| benzodiazepine_related_sleeping_agents | 30 | latest | 2 | 0 | 0.03 | 0.97 | 0 | 0 | 0 |
| benzodiazepine_related_sleeping_agents | 1825 | mean | 2 | 0 | 0.22 | 0.78 | 0 | 0 | 0 |
| benzodiazepine_related_sleeping_agents | 1095 | mean | 2 | 0 | 0.2 | 0.8 | 0 | 0 | 0 |
| benzodiazepine_related_sleeping_agents | 730 | mean | 2 | 0 | 0.17 | 0.83 | 0 | 0 | 0 |
| benzodiazepine_related_sleeping_agents | 365 | mean | 2 | 0 | 0.13 | 0.87 | 0 | 0 | 0 |
| benzodiazepine_related_sleeping_agents | 180 | latest | 2 | 0 | 0.09 | 0.91 | 0 | 0 | 0 |
| benzodiazepines | 30 | maximum | 2 | 0 | 0.03 | 0.97 | 0 | 0 | 0 |
| benzodiazepines | 180 | maximum | 2 | 0 | 0.1 | 0.9 | 0 | 0 | 0 |

| | | | | | | | | | |
|---|---|---|---|---|---|---|---|---|---|
| benzodiazepines | 730 | maximum | 2 | 0 | 0.19 | 0.81 | 0 | 0 | 0 |
| benzodiazepines | 1095 | maximum | 2 | 0 | 0.21 | 0.79 | 0 | 0 | 0 |
| benzodiazepines | 1460 | maximum | 2 | 0 | 0.23 | 0.77 | 0 | 0 | 0 |
| benzodiazepines | 1460 | latest | 2 | 0 | 0.23 | 0.77 | 0 | 0 | 0 |
| benzodiazepines | 730 | minimum | 2 | 0 | 0.19 | 0.81 | 0 | 0 | 0 |
| benzodiazepines | 1825 | maximum | 2 | 0 | 0.24 | 0.76 | 0 | 0 | 0 |
| benzodiazepines | 1095 | minimum | 2 | 0 | 0.21 | 0.79 | 0 | 0 | 0 |
| benzodiazepines | 180 | minimum | 2 | 0 | 0.1 | 0.9 | 0 | 0 | 0 |
| benzodiazepines | 365 | minimum | 2 | 0 | 0.15 | 0.85 | 0 | 0 | 0 |
| benzodiazepines | 1825 | minimum | 2 | 0 | 0.24 | 0.76 | 0 | 0 | 0 |
| benzodiazepines | 30 | minimum | 2 | 0 | 0.03 | 0.97 | 0 | 0 | 0 |
| benzodiazepines | 1460 | minimum | 2 | 0 | 0.23 | 0.77 | 0 | 0 | 0 |
| benzodiazepines | 730 | latest | 2 | 0 | 0.19 | 0.81 | 0 | 0 | 0 |
| benzodiazepines | 30 | mean | 2 | 0 | 0.03 | 0.97 | 0 | 0 | 0 |
| benzodiazepines | 30 | latest | 2 | 0 | 0.03 | 0.97 | 0 | 0 | 0 |
| benzodiazepines | 180 | mean | 2 | 0 | 0.1 | 0.9 | 0 | 0 | 0 |
| benzodiazepines | 1095 | latest | 2 | 0 | 0.21 | 0.79 | 0 | 0 | 0 |
| benzodiazepines | 365 | latest | 2 | 0 | 0.15 | 0.85 | 0 | 0 | 0 |
| benzodiazepines | 180 | latest | 2 | 0 | 0.1 | 0.9 | 0 | 0 | 0 |
| benzodiazepines | 365 | maximum | 2 | 0 | 0.15 | 0.85 | 0 | 0 | 0 |
| benzodiazepines | 1825 | mean | 2 | 0 | 0.24 | 0.76 | 0 | 0 | 0 |
| benzodiazepines | 1095 | mean | 2 | 0 | 0.21 | 0.79 | 0 | 0 | 0 |
| benzodiazepines | 1825 | latest | 2 | 0 | 0.24 | 0.76 | 0 | 0 | 0 |
| benzodiazepines | 1460 | mean | 2 | 0 | 0.23 | 0.77 | 0 | 0 | 0 |
| benzodiazepines | 365 | mean | 2 | 0 | 0.15 | 0.85 | 0 | 0 | 0 |
| benzodiazepines | 730 | mean | 2 | 0 | 0.19 | 0.81 | 0 | 0 | 0 |
| bmi | 1825 | latest | 520 | nan | 26.36 | 0.22 | 21.6 | 25.2 | 29.8 |
| bmi | 1095 | latest | 520 | nan | 26.4 | 0.27 | 21.6 | 25.2 | 29.8 |

| | | | | | | | | | |
|---|---|---|---|---|---|---|---|---|---|
| bmi | 1460 | latest | 520 | nan | 26.38 | 0.24 | 21.6 | 25.2 | 29.8 |
| bmi | 730 | latest | 519 | nan | 26.41 | 0.32 | 21.6 | 25.2 | 29.9 |
| bmi | 365 | latest | 517 | nan | 26.38 | 0.44 | 21.6 | 25.2 | 29.9 |
| bmi | 180 | latest | 515 | nan | 26.22 | 0.59 | 21.3 | 25 | 29.8 |
| bmi | 30 | latest | 499 | nan | 24.89 | 0.87 | 20 | 23.5 | 28.5 |
| clozapine | 30 | latest | 2 | 0 | 0 | 1 | 0 | 0 | 0 |
| clozapine | 1460 | minimum | 2 | 0 | 0.02 | 0.98 | 0 | 0 | 0 |
| clozapine | 1825 | latest | 2 | 0 | 0.02 | 0.98 | 0 | 0 | 0 |
| clozapine | 1460 | latest | 2 | 0 | 0.02 | 0.98 | 0 | 0 | 0 |
| clozapine | 1095 | latest | 2 | 0 | 0.02 | 0.98 | 0 | 0 | 0 |
| clozapine | 730 | latest | 2 | 0 | 0.02 | 0.98 | 0 | 0 | 0 |
| clozapine | 365 | latest | 2 | 0 | 0.01 | 0.99 | 0 | 0 | 0 |
| clozapine | 180 | latest | 2 | 0 | 0.01 | 0.99 | 0 | 0 | 0 |
| clozapine | 1460 | mean | 2 | 0 | 0.02 | 0.98 | 0 | 0 | 0 |
| clozapine | 1095 | mean | 2 | 0 | 0.02 | 0.98 | 0 | 0 | 0 |
| clozapine | 365 | mean | 2 | 0 | 0.01 | 0.99 | 0 | 0 | 0 |
| clozapine | 180 | mean | 2 | 0 | 0.01 | 0.99 | 0 | 0 | 0 |
| clozapine | 30 | mean | 2 | 0 | 0 | 1 | 0 | 0 | 0 |
| clozapine | 1825 | mean | 2 | 0 | 0.02 | 0.98 | 0 | 0 | 0 |
| clozapine | 730 | mean | 2 | 0 | 0.02 | 0.98 | 0 | 0 | 0 |
| clozapine | 1825 | minimum | 2 | 0 | 0.02 | 0.98 | 0 | 0 | 0 |
| clozapine | 365 | minimum | 2 | 0 | 0.01 | 0.99 | 0 | 0 | 0 |
| clozapine | 180 | minimum | 2 | 0 | 0.01 | 0.99 | 0 | 0 | 0 |
| clozapine | 30 | minimum | 2 | 0 | 0 | 1 | 0 | 0 | 0 |
| clozapine | 180 | maximum | 2 | 0 | 0.01 | 0.99 | 0 | 0 | 0 |
| clozapine | 1095 | maximum | 2 | 0 | 0.02 | 0.98 | 0 | 0 | 0 |
| clozapine | 1825 | maximum | 2 | 0 | 0.02 | 0.98 | 0 | 0 | 0 |
| clozapine | 365 | maximum | 2 | 0 | 0.01 | 0.99 | 0 | 0 | 0 |

| | | | | | | | | | |
|---|---|---|---|---|---|---|---|---|---|
| clozapine | 730 | minimum | 2 | 0 | 0.02 | 0.98 | 0 | 0 | 0 |
| clozapine | 30 | maximum | 2 | 0 | 0 | 1 | 0 | 0 | 0 |
| clozapine | 1460 | maximum | 2 | 0 | 0.02 | 0.98 | 0 | 0 | 0 |
| clozapine | 1095 | minimum | 2 | 0 | 0.02 | 0.98 | 0 | 0 | 0 |
| clozapine | 730 | maximum | 2 | 0 | 0.02 | 0.98 | 0 | 0 | 0 |
| crp | 730 | minimum | 867 | nan | 2.96 | 0.19 | 0.7 | 1.6 | 2.7 |
| crp | 365 | minimum | 913 | nan | 3.45 | 0.31 | 0.7 | 2.3 | 2.7 |
| crp | 1095 | latest | 1462 | nan | 5.15 | 0.15 | 0.7 | 2.7 | 4.1 |
| crp | 180 | minimum | 960 | nan | 3.88 | 0.49 | 0.7 | 2.7 | 3.1 |
| crp | 30 | minimum | 957 | nan | 4.82 | 0.86 | 0.9 | 2.7 | 4.1 |
| crp | 1825 | maximum | 3053 | nan | 22.81 | 0.12 | 1.8 | 3.7 | 14.1 |
| crp | 730 | maximum | 2807 | nan | 15.82 | 0.19 | 1.4 | 2.7 | 8.9 |
| crp | 1095 | maximum | 2940 | nan | 18.95 | 0.15 | 1.6 | 3 | 10.9 |
| crp | 365 | maximum | 2521 | nan | 11.68 | 0.31 | 1.2 | 2.7 | 6.6 |
| crp | 180 | maximum | 2292 | nan | 9.47 | 0.49 | 1.1 | 2.7 | 5.6 |
| crp | 1460 | latest | 1480 | nan | 5.18 | 0.13 | 0.7 | 2.7 | 4.1 |
| crp | 1095 | minimum | 838 | nan | 2.67 | 0.15 | 0.6 | 1.2 | 2.7 |
| crp | 1460 | maximum | 3012 | nan | 21.22 | 0.13 | 1.7 | 3.4 | 12.6 |
| crp | 1460 | minimum | 816 | nan | 2.48 | 0.13 | 0.5 | 1 | 2.7 |
| crp | 1095 | mean | 45240 | nan | 7.85 | 0.15 | 1.1 | 2.7 | 6.1 |
| crp | 180 | mean | 16737 | nan | 6.1 | 0.49 | 1 | 2.7 | 4.7 |
| crp | 30 | maximum | 1671 | nan | 7.68 | 0.86 | 1.1 | 2.7 | 4.9 |
| crp | 30 | mean | 5346 | nan | 6.12 | 0.86 | 1 | 2.7 | 4.6 |
| crp | 1825 | latest | 1484 | nan | 5.18 | 0.12 | 0.7 | 2.7 | 4.1 |
| crp | 730 | latest | 1434 | nan | 5.09 | 0.19 | 0.8 | 2.7 | 4.1 |
| crp | 365 | latest | 1360 | nan | 5.02 | 0.31 | 0.8 | 2.7 | 4.1 |
| crp | 1825 | minimum | 793 | nan | 2.34 | 0.12 | 0.4 | 0.9 | 2.7 |
| crp | 30 | latest | 1143 | nan | 5.56 | 0.86 | 1 | 2.7 | 4.5 |

| | | | | | | | | | |
|---|---|---|---|---|---|---|---|---|---|
| crp | 1825 | mean | 52604 | nan | 8.39 | 0.12 | 1.2 | 2.7 | 6.7 |
| crp | 1460 | mean | 50002 | nan | 8.21 | 0.13 | 1.2 | 2.7 | 6.5 |
| crp | 730 | mean | 38051 | nan | 7.3 | 0.19 | 1.1 | 2.7 | 5.6 |
| crp | 365 | mean | 25833 | nan | 6.49 | 0.31 | 1.1 | 2.7 | 5 |
| crp | 180 | latest | 1300 | nan | 5.04 | 0.49 | 0.9 | 2.7 | 4.1 |
| diuretics | 365 | mean | 2 | 0 | 0.02 | 0.98 | 0 | 0 | 0 |
| diuretics | 730 | minimum | 2 | 0 | 0.03 | 0.97 | 0 | 0 | 0 |
| diuretics | 365 | maximum | 2 | 0 | 0.02 | 0.98 | 0 | 0 | 0 |
| diuretics | 1095 | minimum | 2 | 0 | 0.03 | 0.97 | 0 | 0 | 0 |
| diuretics | 30 | maximum | 2 | 0 | 0 | 1 | 0 | 0 | 0 |
| diuretics | 730 | maximum | 2 | 0 | 0.03 | 0.97 | 0 | 0 | 0 |
| diuretics | 1460 | maximum | 2 | 0 | 0.03 | 0.97 | 0 | 0 | 0 |
| diuretics | 30 | minimum | 2 | 0 | 0 | 1 | 0 | 0 | 0 |
| diuretics | 180 | minimum | 2 | 0 | 0.01 | 0.99 | 0 | 0 | 0 |
| diuretics | 365 | minimum | 2 | 0 | 0.02 | 0.98 | 0 | 0 | 0 |
| diuretics | 1460 | minimum | 2 | 0 | 0.03 | 0.97 | 0 | 0 | 0 |
| diuretics | 1095 | maximum | 2 | 0 | 0.03 | 0.97 | 0 | 0 | 0 |
| diuretics | 1825 | minimum | 2 | 0 | 0.04 | 0.96 | 0 | 0 | 0 |
| diuretics | 1825 | maximum | 2 | 0 | 0.04 | 0.96 | 0 | 0 | 0 |
| diuretics | 180 | mean | 2 | 0 | 0.01 | 0.99 | 0 | 0 | 0 |
| diuretics | 1460 | latest | 2 | 0 | 0.03 | 0.97 | 0 | 0 | 0 |
| diuretics | 1825 | latest | 2 | 0 | 0.04 | 0.96 | 0 | 0 | 0 |
| diuretics | 1095 | latest | 2 | 0 | 0.03 | 0.97 | 0 | 0 | 0 |
| diuretics | 730 | latest | 2 | 0 | 0.03 | 0.97 | 0 | 0 | 0 |
| diuretics | 180 | latest | 2 | 0 | 0.01 | 0.99 | 0 | 0 | 0 |
| diuretics | 180 | maximum | 2 | 0 | 0.01 | 0.99 | 0 | 0 | 0 |
| diuretics | 30 | mean | 2 | 0 | 0 | 1 | 0 | 0 | 0 |
| diuretics | 1825 | mean | 2 | 0 | 0.04 | 0.96 | 0 | 0 | 0 |

| | | | | | | | | | |
|---|---|---|---|---|---|---|---|---|---|
| diuretics | 1460 | mean | 2 | 0 | 0.03 | 0.97 | 0 | 0 | 0 |
| diuretics | 1095 | mean | 2 | 0 | 0.03 | 0.97 | 0 | 0 | 0 |
| diuretics | 730 | mean | 2 | 0 | 0.03 | 0.97 | 0 | 0 | 0 |
| diuretics | 365 | latest | 2 | 0 | 0.02 | 0.98 | 0 | 0 | 0 |
| diuretics | 30 | latest | 2 | 0 | 0 | 1 | 0 | 0 | 0 |
| egfr | 730 | minimum | 162 | nan | 105.44 | 0.09 | 80 | 108 | 135 |
| egfr | 365 | minimum | 167 | nan | 108.96 | 0.18 | 83 | 108 | 135 |
| egfr | 180 | minimum | 163 | nan | 110.8 | 0.35 | 84 | 135 | 135 |
| egfr | 30 | minimum | 150 | nan | 111.89 | 0.8 | 85 | 135 | 135 |
| egfr | 1825 | maximum | 220 | nan | 123.27 | 0.06 | 108 | 135 | 135 |
| egfr | 30 | maximum | 153 | nan | 114.03 | 0.8 | 87 | 135 | 135 |
| egfr | 1460 | latest | 174 | nan | 114.81 | 0.06 | 88 | 135 | 135 |
| egfr | 730 | maximum | 213 | nan | 121.3 | 0.09 | 108 | 135 | 135 |
| egfr | 30 | mean | 1645 | nan | 113.01 | 0.8 | 86.5 | 135 | 135 |
| egfr | 1825 | latest | 174 | nan | 114.8 | 0.06 | 88 | 135 | 135 |
| egfr | 1460 | maximum | 220 | nan | 122.93 | 0.06 | 108 | 135 | 135 |
| egfr | 1095 | minimum | 161 | nan | 102.68 | 0.07 | 78 | 108 | 135 |
| egfr | 180 | maximum | 187 | nan | 117.18 | 0.35 | 102 | 135 | 135 |
| egfr | 1095 | maximum | 211 | nan | 122.35 | 0.07 | 108 | 135 | 135 |
| egfr | 1460 | minimum | 161 | nan | 100.64 | 0.06 | 77 | 99 | 135 |
| egfr | 365 | maximum | 197 | nan | 119.16 | 0.18 | 108 | 135 | 135 |
| egfr | 1825 | minimum | 160 | nan | 99.02 | 0.06 | 77 | 90 | 135 |
| egfr | 730 | latest | 174 | nan | 114.84 | 0.09 | 88 | 135 | 135 |
| egfr | 1095 | latest | 174 | nan | 114.83 | 0.07 | 88 | 135 | 135 |
| egfr | 365 | latest | 171 | nan | 114.75 | 0.18 | 88 | 135 | 135 |
| egfr | 30 | latest | 151 | nan | 113.15 | 0.8 | 86 | 135 | 135 |
| egfr | 180 | latest | 167 | nan | 114.36 | 0.35 | 87 | 135 | 135 |
| egfr | 1460 | mean | 22825 | nan | 112.97 | 0.06 | 97.4 | 118 | 135 |

| | | | | | | | | | |
|---|---|---|---|---|---|---|---|---|---|
| egfr | 1095 | mean | 19821 | nan | 113.49 | 0.07 | 97.8 | 119.5 | 135 |
| egfr | 730 | mean | 15869 | nan | 114.06 | 0.09 | 97.8 | 121.5 | 135 |
| egfr | 365 | mean | 10031 | nan | 114.44 | 0.18 | 96 | 128.2 | 135 |
| egfr | 180 | mean | 6058 | nan | 114.2 | 0.35 | 90 | 135 | 135 |
| egfr | 1825 | mean | 24949 | nan | 112.52 | 0.06 | 97 | 117 | 135 |
| essential_hypertension | 1095 | maximum | 2 | 0 | 0.02 | 0.98 | 0 | 0 | 0 |
| essential_hypertension | 30 | minimum | 2 | 0 | 0 | 1 | 0 | 0 | 0 |
| essential_hypertension | 1825 | maximum | 2 | 0 | 0.02 | 0.98 | 0 | 0 | 0 |
| essential_hypertension | 1460 | maximum | 2 | 0 | 0.02 | 0.98 | 0 | 0 | 0 |
| essential_hypertension | 730 | maximum | 2 | 0 | 0.01 | 0.99 | 0 | 0 | 0 |
| essential_hypertension | 180 | minimum | 2 | 0 | 0.01 | 0.99 | 0 | 0 | 0 |
| essential_hypertension | 180 | maximum | 2 | 0 | 0.01 | 0.99 | 0 | 0 | 0 |
| essential_hypertension | 30 | maximum | 2 | 0 | 0 | 1 | 0 | 0 | 0 |
| essential_hypertension | 1095 | latest | 2 | 0 | 0.02 | 0.98 | 0 | 0 | 0 |
| essential_hypertension | 1460 | minimum | 2 | 0 | 0.02 | 0.98 | 0 | 0 | 0 |
| essential_hypertension | 1825 | latest | 2 | 0 | 0.02 | 0.98 | 0 | 0 | 0 |
| essential_hypertension | 365 | maximum | 2 | 0 | 0.01 | 0.99 | 0 | 0 | 0 |
| essential_hypertension | 365 | minimum | 2 | 0 | 0.01 | 0.99 | 0 | 0 | 0 |
| essential_hypertension | 730 | latest | 2 | 0 | 0.01 | 0.99 | 0 | 0 | 0 |
| essential_hypertension | 1095 | minimum | 2 | 0 | 0.02 | 0.98 | 0 | 0 | 0 |
| essential_hypertension | 365 | latest | 2 | 0 | 0.01 | 0.99 | 0 | 0 | 0 |
| essential_hypertension | 180 | latest | 2 | 0 | 0.01 | 0.99 | 0 | 0 | 0 |
| essential_hypertension | 730 | minimum | 2 | 0 | 0.01 | 0.99 | 0 | 0 | 0 |
| essential_hypertension | 30 | latest | 2 | 0 | 0 | 1 | 0 | 0 | 0 |
| essential_hypertension | 1825 | mean | 2 | 0 | 0.02 | 0.98 | 0 | 0 | 0 |
| essential_hypertension | 1460 | mean | 2 | 0 | 0.02 | 0.98 | 0 | 0 | 0 |
| essential_hypertension | 1460 | latest | 2 | 0 | 0.02 | 0.98 | 0 | 0 | 0 |
| essential_hypertension | 730 | mean | 2 | 0 | 0.01 | 0.99 | 0 | 0 | 0 |

| | | | | | | | | | |
|---|---|---|---|---|---|---|---|---|---|
| essential_hypertension | 365 | mean | 2 | 0 | 0.01 | 0.99 | 0 | 0 | 0 |
| essential_hypertension | 180 | mean | 2 | 0 | 0.01 | 0.99 | 0 | 0 | 0 |
| essential_hypertension | 30 | mean | 2 | 0 | 0 | 1 | 0 | 0 | 0 |
| essential_hypertension | 1825 | minimum | 2 | 0 | 0.02 | 0.98 | 0 | 0 | 0 |
| essential_hypertension | 1095 | mean | 2 | 0 | 0.02 | 0.98 | 0 | 0 | 0 |
| f0 - Organic disorders | 30 | maximum | 2 | 0 | 0 | 1 | 0 | 0 | 0 |
| f0 - Organic disorders | 180 | maximum | 2 | 0 | 0.01 | 0.99 | 0 | 0 | 0 |
| f0 - Organic disorders | 365 | maximum | 2 | 0 | 0.02 | 0.98 | 0 | 0 | 0 |
| f0 - Organic disorders | 730 | maximum | 2 | 0 | 0.02 | 0.98 | 0 | 0 | 0 |
| f0 - Organic disorders | 1095 | maximum | 2 | 0 | 0.03 | 0.97 | 0 | 0 | 0 |
| f0 - Organic disorders | 30 | minimum | 2 | 0 | 0 | 1 | 0 | 0 | 0 |
| f0 - Organic disorders | 1825 | maximum | 2 | 0 | 0.03 | 0.97 | 0 | 0 | 0 |
| f0 - Organic disorders | 180 | minimum | 2 | 0 | 0.01 | 0.99 | 0 | 0 | 0 |
| f0 - Organic disorders | 730 | minimum | 2 | 0 | 0.02 | 0.98 | 0 | 0 | 0 |
| f0 - Organic disorders | 1095 | minimum | 2 | 0 | 0.03 | 0.97 | 0 | 0 | 0 |
| f0 - Organic disorders | 30 | mean | 2 | 0 | 0 | 1 | 0 | 0 | 0 |
| f0 - Organic disorders | 1460 | maximum | 2 | 0 | 0.03 | 0.97 | 0 | 0 | 0 |
| f0 - Organic disorders | 180 | mean | 2 | 0 | 0.01 | 0.99 | 0 | 0 | 0 |
| f0 - Organic disorders | 1825 | minimum | 2 | 0 | 0.03 | 0.97 | 0 | 0 | 0 |
| f0 - Organic disorders | 730 | mean | 2 | 0 | 0.02 | 0.98 | 0 | 0 | 0 |
| f0 - Organic disorders | 365 | minimum | 2 | 0 | 0.02 | 0.98 | 0 | 0 | 0 |
| f0 - Organic disorders | 1460 | minimum | 2 | 0 | 0.03 | 0.97 | 0 | 0 | 0 |
| f0 - Organic disorders | 1825 | latest | 2 | 0 | 0.03 | 0.97 | 0 | 0 | 0 |
| f0 - Organic disorders | 1460 | latest | 2 | 0 | 0.03 | 0.97 | 0 | 0 | 0 |
| f0 - Organic disorders | 1095 | latest | 2 | 0 | 0.03 | 0.97 | 0 | 0 | 0 |
| f0 - Organic disorders | 730 | latest | 2 | 0 | 0.02 | 0.98 | 0 | 0 | 0 |
| f0 - Organic disorders | 365 | mean | 2 | 0 | 0.02 | 0.98 | 0 | 0 | 0 |
| f0 - Organic disorders | 180 | latest | 2 | 0 | 0.01 | 0.99 | 0 | 0 | 0 |

| | | | | | | | | | |
|---|---|---|---|---|---|---|---|---|---|
| f0 - Organic disorders | 30 | latest | 2 | 0 | 0 | 1 | 0 | 0 | 0 |
| f0 - Organic disorders | 1825 | mean | 2 | 0 | 0.03 | 0.97 | 0 | 0 | 0 |
| f0 - Organic disorders | 1460 | mean | 2 | 0 | 0.03 | 0.97 | 0 | 0 | 0 |
| f0 - Organic disorders | 1095 | mean | 2 | 0 | 0.03 | 0.97 | 0 | 0 | 0 |
| f0 - Organic disorders | 365 | latest | 2 | 0 | 0.02 | 0.98 | 0 | 0 | 0 |
| f1 - Substance abuse | 1095 | minimum | 2 | 0 | 0.12 | 0.88 | 0 | 0 | 0 |
| f1 - Substance abuse | 730 | minimum | 2 | 0 | 0.11 | 0.89 | 0 | 0 | 0 |
| f1 - Substance abuse | 365 | minimum | 2 | 0 | 0.09 | 0.91 | 0 | 0 | 0 |
| f1 - Substance abuse | 180 | minimum | 2 | 0 | 0.06 | 0.94 | 0 | 0 | 0 |
| f1 - Substance abuse | 30 | minimum | 2 | 0 | 0.03 | 0.97 | 0 | 0 | 0 |
| f1 - Substance abuse | 1825 | maximum | 2 | 0 | 0.14 | 0.86 | 0 | 0 | 0 |
| f1 - Substance abuse | 730 | maximum | 2 | 0 | 0.11 | 0.89 | 0 | 0 | 0 |
| f1 - Substance abuse | 1095 | maximum | 2 | 0 | 0.12 | 0.88 | 0 | 0 | 0 |
| f1 - Substance abuse | 365 | maximum | 2 | 0 | 0.09 | 0.91 | 0 | 0 | 0 |
| f1 - Substance abuse | 180 | maximum | 2 | 0 | 0.06 | 0.94 | 0 | 0 | 0 |
| f1 - Substance abuse | 30 | maximum | 2 | 0 | 0.03 | 0.97 | 0 | 0 | 0 |
| f1 - Substance abuse | 180 | mean | 2 | 0 | 0.06 | 0.94 | 0 | 0 | 0 |
| f1 - Substance abuse | 1460 | maximum | 2 | 0 | 0.13 | 0.87 | 0 | 0 | 0 |
| f1 - Substance abuse | 1460 | minimum | 2 | 0 | 0.13 | 0.87 | 0 | 0 | 0 |
| f1 - Substance abuse | 1825 | minimum | 2 | 0 | 0.14 | 0.86 | 0 | 0 | 0 |
| f1 - Substance abuse | 1460 | latest | 2 | 0 | 0.13 | 0.87 | 0 | 0 | 0 |
| f1 - Substance abuse | 365 | mean | 2 | 0 | 0.09 | 0.91 | 0 | 0 | 0 |
| f1 - Substance abuse | 1825 | latest | 2 | 0 | 0.14 | 0.86 | 0 | 0 | 0 |
| f1 - Substance abuse | 730 | mean | 2 | 0 | 0.11 | 0.89 | 0 | 0 | 0 |
| f1 - Substance abuse | 1460 | mean | 2 | 0 | 0.13 | 0.87 | 0 | 0 | 0 |
| f1 - Substance abuse | 1825 | mean | 2 | 0 | 0.14 | 0.86 | 0 | 0 | 0 |
| f1 - Substance abuse | 1095 | mean | 2 | 0 | 0.12 | 0.88 | 0 | 0 | 0 |
| f1 - Substance abuse | 180 | latest | 2 | 0 | 0.06 | 0.94 | 0 | 0 | 0 |

| | | | | | | | | | |
|---|---|---|---|---|---|---|---|---|---|
| f1 - Substance abuse | 365 | latest | 2 | 0 | 0.09 | 0.91 | 0 | 0 | 0 |
| f1 - Substance abuse | 730 | latest | 2 | 0 | 0.11 | 0.89 | 0 | 0 | 0 |
| f1 - Substance abuse | 1095 | latest | 2 | 0 | 0.12 | 0.88 | 0 | 0 | 0 |
| f1 - Substance abuse | 30 | mean | 2 | 0 | 0.03 | 0.97 | 0 | 0 | 0 |
| f1 - Substance abuse | 30 | latest | 2 | 0 | 0.03 | 0.97 | 0 | 0 | 0 |
| f2 - Psychotic disorders | 730 | latest | 2 | 0 | 0.16 | 0.84 | 0 | 0 | 0 |
| f2 - Psychotic disorders | 1825 | mean | 2 | 0 | 0.18 | 0.82 | 0 | 0 | 0 |
| f2 - Psychotic disorders | 30 | latest | 2 | 0 | 0.06 | 0.94 | 0 | 0 | 0 |
| f2 - Psychotic disorders | 180 | latest | 2 | 0 | 0.11 | 0.89 | 0 | 0 | 0 |
| f2 - Psychotic disorders | 365 | latest | 2 | 0 | 0.13 | 0.87 | 0 | 0 | 0 |
| f2 - Psychotic disorders | 1460 | minimum | 2 | 0 | 0.17 | 0.83 | 0 | 0 | 0 |
| f2 - Psychotic disorders | 180 | maximum | 2 | 0 | 0.11 | 0.89 | 0 | 0 | 0 |
| f2 - Psychotic disorders | 1825 | latest | 2 | 0 | 0.18 | 0.82 | 0 | 0 | 0 |
| f2 - Psychotic disorders | 365 | maximum | 2 | 0 | 0.13 | 0.87 | 0 | 0 | 0 |
| f2 - Psychotic disorders | 30 | maximum | 2 | 0 | 0.06 | 0.94 | 0 | 0 | 0 |
| f2 - Psychotic disorders | 1460 | latest | 2 | 0 | 0.17 | 0.83 | 0 | 0 | 0 |
| f2 - Psychotic disorders | 1460 | mean | 2 | 0 | 0.17 | 0.83 | 0 | 0 | 0 |
| f2 - Psychotic disorders | 1095 | latest | 2 | 0 | 0.17 | 0.83 | 0 | 0 | 0 |
| f2 - Psychotic disorders | 1095 | mean | 2 | 0 | 0.17 | 0.83 | 0 | 0 | 0 |
| f2 - Psychotic disorders | 730 | mean | 2 | 0 | 0.16 | 0.84 | 0 | 0 | 0 |
| f2 - Psychotic disorders | 365 | mean | 2 | 0 | 0.13 | 0.87 | 0 | 0 | 0 |
| f2 - Psychotic disorders | 180 | minimum | 2 | 0 | 0.11 | 0.89 | 0 | 0 | 0 |
| f2 - Psychotic disorders | 30 | mean | 2 | 0 | 0.06 | 0.94 | 0 | 0 | 0 |
| f2 - Psychotic disorders | 1825 | maximum | 2 | 0 | 0.18 | 0.82 | 0 | 0 | 0 |
| f2 - Psychotic disorders | 730 | minimum | 2 | 0 | 0.16 | 0.84 | 0 | 0 | 0 |
| f2 - Psychotic disorders | 1460 | maximum | 2 | 0 | 0.17 | 0.83 | 0 | 0 | 0 |
| f2 - Psychotic disorders | 365 | minimum | 2 | 0 | 0.13 | 0.87 | 0 | 0 | 0 |
| f2 - Psychotic disorders | 730 | maximum | 2 | 0 | 0.16 | 0.84 | 0 | 0 | 0 |

| | | | | | | | | | |
|---|---|---|---|---|---|---|---|---|---|
| f2 - Psychotic disorders | 30 | minimum | 2 | 0 | 0.06 | 0.94 | 0 | 0 | 0 |
| f2 - Psychotic disorders | 1095 | minimum | 2 | 0 | 0.17 | 0.83 | 0 | 0 | 0 |
| f2 - Psychotic disorders | 180 | mean | 2 | 0 | 0.11 | 0.89 | 0 | 0 | 0 |
| f2 - Psychotic disorders | 1825 | minimum | 2 | 0 | 0.18 | 0.82 | 0 | 0 | 0 |
| f2 - Psychotic disorders | 1095 | maximum | 2 | 0 | 0.17 | 0.83 | 0 | 0 | 0 |
| f3 - Mood disorders | 730 | mean | 2 | 0 | 0.39 | 0.61 | 0 | 0 | 1 |
| f3 - Mood disorders | 1095 | mean | 2 | 0 | 0.42 | 0.58 | 0 | 0 | 1 |
| f3 - Mood disorders | 730 | maximum | 2 | 0 | 0.39 | 0.61 | 0 | 0 | 1 |
| f3 - Mood disorders | 1825 | mean | 2 | 0 | 0.45 | 0.55 | 0 | 0 | 1 |
| f3 - Mood disorders | 1460 | mean | 2 | 0 | 0.44 | 0.56 | 0 | 0 | 1 |
| f3 - Mood disorders | 1095 | latest | 2 | 0 | 0.42 | 0.58 | 0 | 0 | 1 |
| f3 - Mood disorders | 1460 | latest | 2 | 0 | 0.44 | 0.56 | 0 | 0 | 1 |
| f3 - Mood disorders | 365 | latest | 2 | 0 | 0.32 | 0.68 | 0 | 0 | 1 |
| f3 - Mood disorders | 180 | latest | 2 | 0 | 0.25 | 0.75 | 0 | 0 | 0 |
| f3 - Mood disorders | 730 | latest | 2 | 0 | 0.39 | 0.61 | 0 | 0 | 1 |
| f3 - Mood disorders | 30 | maximum | 2 | 0 | 0.12 | 0.88 | 0 | 0 | 0 |
| f3 - Mood disorders | 1825 | latest | 2 | 0 | 0.45 | 0.55 | 0 | 0 | 1 |
| f3 - Mood disorders | 180 | maximum | 2 | 0 | 0.25 | 0.75 | 0 | 0 | 0 |
| f3 - Mood disorders | 30 | latest | 2 | 0 | 0.12 | 0.88 | 0 | 0 | 0 |
| f3 - Mood disorders | 1095 | maximum | 2 | 0 | 0.42 | 0.58 | 0 | 0 | 1 |
| f3 - Mood disorders | 365 | mean | 2 | 0 | 0.32 | 0.68 | 0 | 0 | 1 |
| f3 - Mood disorders | 180 | mean | 2 | 0 | 0.25 | 0.75 | 0 | 0 | 0 |
| f3 - Mood disorders | 30 | mean | 2 | 0 | 0.12 | 0.88 | 0 | 0 | 0 |
| f3 - Mood disorders | 1825 | minimum | 2 | 0 | 0.45 | 0.55 | 0 | 0 | 1 |
| f3 - Mood disorders | 1460 | minimum | 2 | 0 | 0.44 | 0.56 | 0 | 0 | 1 |
| f3 - Mood disorders | 1095 | minimum | 2 | 0 | 0.42 | 0.58 | 0 | 0 | 1 |
| f3 - Mood disorders | 365 | maximum | 2 | 0 | 0.32 | 0.68 | 0 | 0 | 1 |
| f3 - Mood disorders | 365 | minimum | 2 | 0 | 0.32 | 0.68 | 0 | 0 | 1 |

| | | | | | | | | | |
|---|---|---|---|---|---|---|---|---|---|
| f3 - Mood disorders | 180 | minimum | 2 | 0 | 0.25 | 0.75 | 0 | 0 | 0 |
| f3 - Mood disorders | 30 | minimum | 2 | 0 | 0.12 | 0.88 | 0 | 0 | 0 |
| f3 - Mood disorders | 1825 | maximum | 2 | 0 | 0.45 | 0.55 | 0 | 0 | 1 |
| f3 - Mood disorders | 1460 | maximum | 2 | 0 | 0.44 | 0.56 | 0 | 0 | 1 |
| f3 - Mood disorders | 730 | minimum | 2 | 0 | 0.39 | 0.61 | 0 | 0 | 1 |
| f4 - Neurotic & stress-related | 180 | maximum | 2 | 0 | 0.19 | 0.81 | 0 | 0 | 0 |
| f4 - Neurotic & stress-related | 365 | maximum | 2 | 0 | 0.26 | 0.74 | 0 | 0 | 1 |
| f4 - Neurotic & stress-related | 1825 | minimum | 2 | 0 | 0.4 | 0.6 | 0 | 0 | 1 |
| f4 - Neurotic & stress-related | 730 | maximum | 2 | 0 | 0.33 | 0.67 | 0 | 0 | 1 |
| f4 - Neurotic & stress-related | 1095 | maximum | 2 | 0 | 0.36 | 0.64 | 0 | 0 | 1 |
| f4 - Neurotic & stress-related | 1095 | latest | 2 | 0 | 0.36 | 0.64 | 0 | 0 | 1 |
| f4 - Neurotic & stress-related | 30 | minimum | 2 | 0 | 0.09 | 0.91 | 0 | 0 | 0 |
| f4 - Neurotic & stress-related | 1095 | minimum | 2 | 0 | 0.36 | 0.64 | 0 | 0 | 1 |
| f4 - Neurotic & stress-related | 180 | minimum | 2 | 0 | 0.19 | 0.81 | 0 | 0 | 0 |
| f4 - Neurotic & stress-related | 730 | minimum | 2 | 0 | 0.33 | 0.67 | 0 | 0 | 1 |
| f4 - Neurotic & stress-related | 365 | minimum | 2 | 0 | 0.26 | 0.74 | 0 | 0 | 1 |
| f4 - Neurotic & stress-related | 30 | maximum | 2 | 0 | 0.09 | 0.91 | 0 | 0 | 0 |
| f4 - Neurotic & stress-related | 1825 | maximum | 2 | 0 | 0.4 | 0.6 | 0 | 0 | 1 |
| f4 - Neurotic & stress-related | 1460 | maximum | 2 | 0 | 0.38 | 0.62 | 0 | 0 | 1 |
| f4 - Neurotic & stress-related | 1460 | minimum | 2 | 0 | 0.38 | 0.62 | 0 | 0 | 1 |
| f4 - Neurotic & stress-related | 30 | mean | 2 | 0 | 0.09 | 0.91 | 0 | 0 | 0 |
| f4 - Neurotic & stress-related | 1460 | mean | 2 | 0 | 0.38 | 0.62 | 0 | 0 | 1 |
| f4 - Neurotic & stress-related | 365 | latest | 2 | 0 | 0.26 | 0.74 | 0 | 0 | 1 |
| f4 - Neurotic & stress-related | 1460 | latest | 2 | 0 | 0.38 | 0.62 | 0 | 0 | 1 |
| f4 - Neurotic & stress-related | 1825 | latest | 2 | 0 | 0.4 | 0.6 | 0 | 0 | 1 |
| f4 - Neurotic & stress-related | 30 | latest | 2 | 0 | 0.09 | 0.91 | 0 | 0 | 0 |
| f4 - Neurotic & stress-related | 180 | latest | 2 | 0 | 0.19 | 0.81 | 0 | 0 | 0 |
| f4 - Neurotic & stress-related | 730 | latest | 2 | 0 | 0.33 | 0.67 | 0 | 0 | 1 |

| | | | | | | | | | |
|---|---|---|---|---|---|---|---|---|---|
| f4 - Neurotic & stress-related | 1095 | mean | 2 | 0 | 0.36 | 0.64 | 0 | 0 | 1 |
| f4 - Neurotic & stress-related | 730 | mean | 2 | 0 | 0.33 | 0.67 | 0 | 0 | 1 |
| f4 - Neurotic & stress-related | 365 | mean | 2 | 0 | 0.26 | 0.74 | 0 | 0 | 1 |
| f4 - Neurotic & stress-related | 180 | mean | 2 | 0 | 0.19 | 0.81 | 0 | 0 | 0 |
| f4 - Neurotic & stress-related | 1825 | mean | 2 | 0 | 0.4 | 0.6 | 0 | 0 | 1 |
| f5 - Eating & sleeping disorders | 1460 | mean | 2 | 0 | 0.07 | 0.93 | 0 | 0 | 0 |
| f5 - Eating & sleeping disorders | 180 | latest | 2 | 0 | 0.04 | 0.96 | 0 | 0 | 0 |
| f5 - Eating & sleeping disorders | 30 | latest | 2 | 0 | 0.02 | 0.98 | 0 | 0 | 0 |
| f5 - Eating & sleeping disorders | 1825 | mean | 2 | 0 | 0.07 | 0.93 | 0 | 0 | 0 |
| f5 - Eating & sleeping disorders | 1095 | minimum | 2 | 0 | 0.06 | 0.94 | 0 | 0 | 0 |
| f5 - Eating & sleeping disorders | 365 | latest | 2 | 0 | 0.05 | 0.95 | 0 | 0 | 0 |
| f5 - Eating & sleeping disorders | 180 | mean | 2 | 0 | 0.04 | 0.96 | 0 | 0 | 0 |
| f5 - Eating & sleeping disorders | 1825 | minimum | 2 | 0 | 0.07 | 0.93 | 0 | 0 | 0 |
| f5 - Eating & sleeping disorders | 1825 | latest | 2 | 0 | 0.07 | 0.93 | 0 | 0 | 0 |
| f5 - Eating & sleeping disorders | 30 | mean | 2 | 0 | 0.02 | 0.98 | 0 | 0 | 0 |
| f5 - Eating & sleeping disorders | 1460 | minimum | 2 | 0 | 0.07 | 0.93 | 0 | 0 | 0 |
| f5 - Eating & sleeping disorders | 730 | mean | 2 | 0 | 0.06 | 0.94 | 0 | 0 | 0 |
| f5 - Eating & sleeping disorders | 730 | latest | 2 | 0 | 0.06 | 0.94 | 0 | 0 | 0 |
| f5 - Eating & sleeping disorders | 1095 | mean | 2 | 0 | 0.06 | 0.94 | 0 | 0 | 0 |
| f5 - Eating & sleeping disorders | 1460 | latest | 2 | 0 | 0.07 | 0.93 | 0 | 0 | 0 |
| f5 - Eating & sleeping disorders | 730 | minimum | 2 | 0 | 0.06 | 0.94 | 0 | 0 | 0 |
| f5 - Eating & sleeping disorders | 30 | maximum | 2 | 0 | 0.02 | 0.98 | 0 | 0 | 0 |
| f5 - Eating & sleeping disorders | 180 | maximum | 2 | 0 | 0.04 | 0.96 | 0 | 0 | 0 |
| f5 - Eating & sleeping disorders | 1095 | maximum | 2 | 0 | 0.06 | 0.94 | 0 | 0 | 0 |
| f5 - Eating & sleeping disorders | 730 | maximum | 2 | 0 | 0.06 | 0.94 | 0 | 0 | 0 |
| f5 - Eating & sleeping disorders | 1460 | maximum | 2 | 0 | 0.07 | 0.93 | 0 | 0 | 0 |
| f5 - Eating & sleeping disorders | 1095 | latest | 2 | 0 | 0.06 | 0.94 | 0 | 0 | 0 |
| f5 - Eating & sleeping disorders | 30 | minimum | 2 | 0 | 0.02 | 0.98 | 0 | 0 | 0 |

| | | | | | | | | | |
|---|---|---|---|---|---|---|---|---|---|
| f5 - Eating & sleeping disorders | 365 | minimum | 2 | 0 | 0.05 | 0.95 | 0 | 0 | 0 |
| f5 - Eating & sleeping disorders | 365 | maximum | 2 | 0 | 0.05 | 0.95 | 0 | 0 | 0 |
| f5 - Eating & sleeping disorders | 180 | minimum | 2 | 0 | 0.04 | 0.96 | 0 | 0 | 0 |
| f5 - Eating & sleeping disorders | 365 | mean | 2 | 0 | 0.05 | 0.95 | 0 | 0 | 0 |
| f5 - Eating & sleeping disorders | 1825 | maximum | 2 | 0 | 0.07 | 0.93 | 0 | 0 | 0 |
| f6 - Perosnality disorders | 730 | latest | 2 | 0 | 0.18 | 0.82 | 0 | 0 | 0 |
| f6 - Perosnality disorders | 730 | maximum | 2 | 0 | 0.18 | 0.82 | 0 | 0 | 0 |
| f6 - Perosnality disorders | 180 | latest | 2 | 0 | 0.12 | 0.88 | 0 | 0 | 0 |
| f6 - Perosnality disorders | 365 | latest | 2 | 0 | 0.15 | 0.85 | 0 | 0 | 0 |
| f6 - Perosnality disorders | 30 | latest | 2 | 0 | 0.07 | 0.93 | 0 | 0 | 0 |
| f6 - Perosnality disorders | 730 | mean | 2 | 0 | 0.18 | 0.82 | 0 | 0 | 0 |
| f6 - Perosnality disorders | 1095 | mean | 2 | 0 | 0.2 | 0.8 | 0 | 0 | 0 |
| f6 - Perosnality disorders | 365 | mean | 2 | 0 | 0.15 | 0.85 | 0 | 0 | 0 |
| f6 - Perosnality disorders | 180 | mean | 2 | 0 | 0.12 | 0.88 | 0 | 0 | 0 |
| f6 - Perosnality disorders | 30 | mean | 2 | 0 | 0.07 | 0.93 | 0 | 0 | 0 |
| f6 - Perosnality disorders | 1095 | latest | 2 | 0 | 0.2 | 0.8 | 0 | 0 | 0 |
| f6 - Perosnality disorders | 1825 | mean | 2 | 0 | 0.21 | 0.79 | 0 | 0 | 0 |
| f6 - Perosnality disorders | 1825 | latest | 2 | 0 | 0.21 | 0.79 | 0 | 0 | 0 |
| f6 - Perosnality disorders | 1825 | minimum | 2 | 0 | 0.21 | 0.79 | 0 | 0 | 0 |
| f6 - Perosnality disorders | 30 | maximum | 2 | 0 | 0.07 | 0.93 | 0 | 0 | 0 |
| f6 - Perosnality disorders | 1460 | mean | 2 | 0 | 0.2 | 0.8 | 0 | 0 | 0 |
| f6 - Perosnality disorders | 1460 | minimum | 2 | 0 | 0.2 | 0.8 | 0 | 0 | 0 |
| f6 - Perosnality disorders | 1095 | minimum | 2 | 0 | 0.2 | 0.8 | 0 | 0 | 0 |
| f6 - Perosnality disorders | 730 | minimum | 2 | 0 | 0.18 | 0.82 | 0 | 0 | 0 |
| f6 - Perosnality disorders | 365 | minimum | 2 | 0 | 0.15 | 0.85 | 0 | 0 | 0 |
| f6 - Perosnality disorders | 180 | minimum | 2 | 0 | 0.12 | 0.88 | 0 | 0 | 0 |
| f6 - Perosnality disorders | 1460 | latest | 2 | 0 | 0.2 | 0.8 | 0 | 0 | 0 |
| f6 - Perosnality disorders | 1825 | maximum | 2 | 0 | 0.21 | 0.79 | 0 | 0 | 0 |

| | | | | | | | | | |
|---|---|---|---|---|---|---|---|---|---|
| f6 - Perosnality disorders | 1460 | maximum | 2 | 0 | 0.2 | 0.8 | 0 | 0 | 0 |
| f6 - Perosnality disorders | 1095 | maximum | 2 | 0 | 0.2 | 0.8 | 0 | 0 | 0 |
| f6 - Perosnality disorders | 365 | maximum | 2 | 0 | 0.15 | 0.85 | 0 | 0 | 0 |
| f6 - Perosnality disorders | 180 | maximum | 2 | 0 | 0.12 | 0.88 | 0 | 0 | 0 |
| f6 - Perosnality disorders | 30 | minimum | 2 | 0 | 0.07 | 0.93 | 0 | 0 | 0 |
| f7 - Mental retardation | 1095 | maximum | 2 | 0 | 0.01 | 0.99 | 0 | 0 | 0 |
| f7 - Mental retardation | 365 | maximum | 2 | 0 | 0.01 | 0.99 | 0 | 0 | 0 |
| f7 - Mental retardation | 180 | maximum | 2 | 0 | 0.01 | 0.99 | 0 | 0 | 0 |
| f7 - Mental retardation | 30 | maximum | 2 | 0 | 0 | 1 | 0 | 0 | 0 |
| f7 - Mental retardation | 1825 | minimum | 2 | 0 | 0.02 | 0.98 | 0 | 0 | 0 |
| f7 - Mental retardation | 30 | mean | 2 | 0 | 0 | 1 | 0 | 0 | 0 |
| f7 - Mental retardation | 730 | mean | 2 | 0 | 0.01 | 0.99 | 0 | 0 | 0 |
| f7 - Mental retardation | 365 | mean | 2 | 0 | 0.01 | 0.99 | 0 | 0 | 0 |
| f7 - Mental retardation | 1460 | maximum | 2 | 0 | 0.02 | 0.98 | 0 | 0 | 0 |
| f7 - Mental retardation | 1095 | mean | 2 | 0 | 0.01 | 0.99 | 0 | 0 | 0 |
| f7 - Mental retardation | 1460 | mean | 2 | 0 | 0.02 | 0.98 | 0 | 0 | 0 |
| f7 - Mental retardation | 730 | maximum | 2 | 0 | 0.01 | 0.99 | 0 | 0 | 0 |
| f7 - Mental retardation | 180 | mean | 2 | 0 | 0.01 | 0.99 | 0 | 0 | 0 |
| f7 - Mental retardation | 1825 | mean | 2 | 0 | 0.02 | 0.98 | 0 | 0 | 0 |
| f7 - Mental retardation | 1825 | maximum | 2 | 0 | 0.02 | 0.98 | 0 | 0 | 0 |
| f7 - Mental retardation | 1825 | latest | 2 | 0 | 0.02 | 0.98 | 0 | 0 | 0 |
| f7 - Mental retardation | 30 | latest | 2 | 0 | 0 | 1 | 0 | 0 | 0 |
| f7 - Mental retardation | 180 | minimum | 2 | 0 | 0.01 | 0.99 | 0 | 0 | 0 |
| f7 - Mental retardation | 365 | minimum | 2 | 0 | 0.01 | 0.99 | 0 | 0 | 0 |
| f7 - Mental retardation | 730 | minimum | 2 | 0 | 0.01 | 0.99 | 0 | 0 | 0 |
| f7 - Mental retardation | 1460 | minimum | 2 | 0 | 0.02 | 0.98 | 0 | 0 | 0 |
| f7 - Mental retardation | 1095 | minimum | 2 | 0 | 0.01 | 0.99 | 0 | 0 | 0 |
| f7 - Mental retardation | 180 | latest | 2 | 0 | 0.01 | 0.99 | 0 | 0 | 0 |

| | | | | | | | | | |
|---|---|---|---|---|---|---|---|---|---|
| f7 - Mental retardation | 365 | latest | 2 | 0 | 0.01 | 0.99 | 0 | 0 | 0 |
| f7 - Mental retardation | 730 | latest | 2 | 0 | 0.01 | 0.99 | 0 | 0 | 0 |
| f7 - Mental retardation | 30 | minimum | 2 | 0 | 0 | 1 | 0 | 0 | 0 |
| f7 - Mental retardation | 1460 | latest | 2 | 0 | 0.02 | 0.98 | 0 | 0 | 0 |
| f7 - Mental retardation | 1095 | latest | 2 | 0 | 0.01 | 0.99 | 0 | 0 | 0 |
| f8 - Developmental disorders | 1095 | latest | 2 | 0 | 0.05 | 0.95 | 0 | 0 | 0 |
| f8 - Developmental disorders | 180 | latest | 2 | 0 | 0.02 | 0.98 | 0 | 0 | 0 |
| f8 - Developmental disorders | 1460 | minimum | 2 | 0 | 0.05 | 0.95 | 0 | 0 | 0 |
| f8 - Developmental disorders | 730 | latest | 2 | 0 | 0.04 | 0.96 | 0 | 0 | 0 |
| f8 - Developmental disorders | 180 | minimum | 2 | 0 | 0.02 | 0.98 | 0 | 0 | 0 |
| f8 - Developmental disorders | 30 | latest | 2 | 0 | 0.01 | 0.99 | 0 | 0 | 0 |
| f8 - Developmental disorders | 365 | mean | 2 | 0 | 0.03 | 0.97 | 0 | 0 | 0 |
| f8 - Developmental disorders | 1095 | minimum | 2 | 0 | 0.05 | 0.95 | 0 | 0 | 0 |
| f8 - Developmental disorders | 730 | minimum | 2 | 0 | 0.04 | 0.96 | 0 | 0 | 0 |
| f8 - Developmental disorders | 365 | minimum | 2 | 0 | 0.03 | 0.97 | 0 | 0 | 0 |
| f8 - Developmental disorders | 365 | latest | 2 | 0 | 0.03 | 0.97 | 0 | 0 | 0 |
| f8 - Developmental disorders | 1825 | latest | 2 | 0 | 0.05 | 0.95 | 0 | 0 | 0 |
| f8 - Developmental disorders | 1825 | mean | 2 | 0 | 0.05 | 0.95 | 0 | 0 | 0 |
| f8 - Developmental disorders | 1460 | latest | 2 | 0 | 0.05 | 0.95 | 0 | 0 | 0 |
| f8 - Developmental disorders | 1095 | mean | 2 | 0 | 0.05 | 0.95 | 0 | 0 | 0 |
| f8 - Developmental disorders | 730 | maximum | 2 | 0 | 0.04 | 0.96 | 0 | 0 | 0 |
| f8 - Developmental disorders | 365 | maximum | 2 | 0 | 0.03 | 0.97 | 0 | 0 | 0 |
| f8 - Developmental disorders | 1095 | maximum | 2 | 0 | 0.05 | 0.95 | 0 | 0 | 0 |
| f8 - Developmental disorders | 30 | maximum | 2 | 0 | 0.01 | 0.99 | 0 | 0 | 0 |
| f8 - Developmental disorders | 180 | maximum | 2 | 0 | 0.02 | 0.98 | 0 | 0 | 0 |
| f8 - Developmental disorders | 1460 | maximum | 2 | 0 | 0.05 | 0.95 | 0 | 0 | 0 |
| f8 - Developmental disorders | 1460 | mean | 2 | 0 | 0.05 | 0.95 | 0 | 0 | 0 |
| f8 - Developmental disorders | 30 | minimum | 2 | 0 | 0.01 | 0.99 | 0 | 0 | 0 |

| | | | | | | | | | |
|---|---|---|---|---|---|---|---|---|---|
| f8 - Developmental disorders | 1825 | maximum | 2 | 0 | 0.05 | 0.95 | 0 | 0 | 0 |
| f8 - Developmental disorders | 30 | mean | 2 | 0 | 0.01 | 0.99 | 0 | 0 | 0 |
| f8 - Developmental disorders | 1825 | minimum | 2 | 0 | 0.05 | 0.95 | 0 | 0 | 0 |
| f8 - Developmental disorders | 730 | mean | 2 | 0 | 0.04 | 0.96 | 0 | 0 | 0 |
| f8 - Developmental disorders | 180 | mean | 2 | 0 | 0.02 | 0.98 | 0 | 0 | 0 |
| f9 - Child & adolescent disorders | 1095 | latest | 2 | 0 | 0.11 | 0.89 | 0 | 0 | 0 |
| f9 - Child & adolescent disorders | 730 | latest | 2 | 0 | 0.1 | 0.9 | 0 | 0 | 0 |
| f9 - Child & adolescent disorders | 365 | latest | 2 | 0 | 0.08 | 0.92 | 0 | 0 | 0 |
| f9 - Child & adolescent disorders | 1460 | latest | 2 | 0 | 0.12 | 0.88 | 0 | 0 | 0 |
| f9 - Child & adolescent disorders | 730 | maximum | 2 | 0 | 0.1 | 0.9 | 0 | 0 | 0 |
| f9 - Child & adolescent disorders | 365 | maximum | 2 | 0 | 0.08 | 0.92 | 0 | 0 | 0 |
| f9 - Child & adolescent disorders | 1825 | latest | 2 | 0 | 0.13 | 0.87 | 0 | 0 | 0 |
| f9 - Child & adolescent disorders | 1095 | maximum | 2 | 0 | 0.11 | 0.89 | 0 | 0 | 0 |
| f9 - Child & adolescent disorders | 1460 | maximum | 2 | 0 | 0.12 | 0.88 | 0 | 0 | 0 |
| f9 - Child & adolescent disorders | 30 | maximum | 2 | 0 | 0.03 | 0.97 | 0 | 0 | 0 |
| f9 - Child & adolescent disorders | 180 | maximum | 2 | 0 | 0.06 | 0.94 | 0 | 0 | 0 |
| f9 - Child & adolescent disorders | 30 | latest | 2 | 0 | 0.03 | 0.97 | 0 | 0 | 0 |
| f9 - Child & adolescent disorders | 180 | minimum | 2 | 0 | 0.06 | 0.94 | 0 | 0 | 0 |
| f9 - Child & adolescent disorders | 180 | latest | 2 | 0 | 0.06 | 0.94 | 0 | 0 | 0 |
| f9 - Child & adolescent disorders | 365 | minimum | 2 | 0 | 0.08 | 0.92 | 0 | 0 | 0 |
| f9 - Child & adolescent disorders | 1825 | mean | 2 | 0 | 0.13 | 0.87 | 0 | 0 | 0 |
| f9 - Child & adolescent disorders | 1460 | mean | 2 | 0 | 0.12 | 0.88 | 0 | 0 | 0 |
| f9 - Child & adolescent disorders | 30 | minimum | 2 | 0 | 0.03 | 0.97 | 0 | 0 | 0 |
| f9 - Child & adolescent disorders | 730 | mean | 2 | 0 | 0.1 | 0.9 | 0 | 0 | 0 |
| f9 - Child & adolescent disorders | 365 | mean | 2 | 0 | 0.08 | 0.92 | 0 | 0 | 0 |
| f9 - Child & adolescent disorders | 180 | mean | 2 | 0 | 0.06 | 0.94 | 0 | 0 | 0 |
| f9 - Child & adolescent disorders | 30 | mean | 2 | 0 | 0.03 | 0.97 | 0 | 0 | 0 |
| f9 - Child & adolescent disorders | 1825 | minimum | 2 | 0 | 0.13 | 0.87 | 0 | 0 | 0 |

| | | | | | | | | | |
|---|---|---|---|---|---|---|---|---|---|
| f9 - Child & adolescent disorders | 1095 | minimum | 2 | 0 | 0.11 | 0.89 | 0 | 0 | 0 |
| f9 - Child & adolescent disorders | 1825 | maximum | 2 | 0 | 0.13 | 0.87 | 0 | 0 | 0 |
| f9 - Child & adolescent disorders | 1095 | mean | 2 | 0 | 0.11 | 0.89 | 0 | 0 | 0 |
| f9 - Child & adolescent disorders | 1460 | minimum | 2 | 0 | 0.12 | 0.88 | 0 | 0 | 0 |
| f9 - Child & adolescent disorders | 730 | minimum | 2 | 0 | 0.1 | 0.9 | 0 | 0 | 0 |
| fasting_ldl | 365 | latest | 77 | nan | 2.88 | 0.91 | 2.2 | 2.8 | 3.4 |
| fasting_ldl | 1095 | latest | 79 | nan | 2.88 | 0.82 | 2.2 | 2.8 | 3.4 |
| fasting_ldl | 730 | latest | 79 | nan | 2.88 | 0.85 | 2.2 | 2.8 | 3.4 |
| fasting_ldl | 180 | latest | 77 | nan | 2.9 | 0.95 | 2.2 | 2.8 | 3.5 |
| fasting_ldl | 30 | latest | 74 | nan | 2.89 | 0.99 | 2.2 | 2.8 | 3.4 |
| fasting_ldl | 1825 | mean | 876 | nan | 2.89 | 0.77 | 2.2 | 2.8 | 3.4 |
| fasting_ldl | 1460 | mean | 848 | nan | 2.89 | 0.79 | 2.2 | 2.8 | 3.4 |
| fasting_ldl | 1095 | mean | 753 | nan | 2.89 | 0.82 | 2.2 | 2.8 | 3.4 |
| fasting_ldl | 365 | mean | 426 | nan | 2.88 | 0.91 | 2.2 | 2.8 | 3.4 |
| fasting_ldl | 180 | mean | 283 | nan | 2.9 | 0.95 | 2.2 | 2.8 | 3.5 |
| fasting_ldl | 30 | mean | 107 | nan | 2.89 | 0.99 | 2.2 | 2.8 | 3.4 |
| fasting_ldl | 730 | mean | 622 | nan | 2.89 | 0.85 | 2.2 | 2.8 | 3.4 |
| fasting_ldl | 1460 | minimum | 75 | nan | 2.76 | 0.79 | 2.1 | 2.7 | 3.3 |
| fasting_ldl | 1460 | maximum | 82 | nan | 3.02 | 0.79 | 2.3 | 2.9 | 3.6 |
| fasting_ldl | 30 | maximum | 74 | nan | 2.89 | 0.99 | 2.2 | 2.8 | 3.4 |
| fasting_ldl | 180 | maximum | 77 | nan | 2.92 | 0.95 | 2.2 | 2.8 | 3.5 |
| fasting_ldl | 365 | maximum | 77 | nan | 2.93 | 0.91 | 2.2 | 2.8 | 3.5 |
| fasting_ldl | 1095 | minimum | 75 | nan | 2.78 | 0.82 | 2.1 | 2.7 | 3.3 |
| fasting_ldl | 1095 | maximum | 81 | nan | 3 | 0.82 | 2.3 | 2.9 | 3.6 |
| fasting_ldl | 730 | maximum | 81 | nan | 2.98 | 0.85 | 2.3 | 2.9 | 3.6 |
| fasting_ldl | 1825 | latest | 79 | nan | 2.88 | 0.77 | 2.2 | 2.8 | 3.4 |
| fasting_ldl | 1825 | maximum | 82 | nan | 3.03 | 0.77 | 2.3 | 2.9 | 3.6 |
| fasting_ldl | 30 | minimum | 74 | nan | 2.89 | 0.99 | 2.2 | 2.8 | 3.4 |

| | | | | | | | | | |
|---|---|---|---|---|---|---|---|---|---|
| fasting_ldl | 180 | minimum | 76 | nan | 2.87 | 0.95 | 2.2 | 2.8 | 3.4 |
| fasting_ldl | 365 | minimum | 76 | nan | 2.83 | 0.91 | 2.2 | 2.8 | 3.4 |
| fasting_ldl | 730 | minimum | 77 | nan | 2.8 | 0.85 | 2.1 | 2.7 | 3.3 |
| fasting_ldl | 1460 | latest | 79 | nan | 2.88 | 0.79 | 2.2 | 2.8 | 3.4 |
| fasting_ldl | 1825 | minimum | 75 | nan | 2.75 | 0.77 | 2.1 | 2.7 | 3.3 |
| fasting_p_glc | 1460 | mean | 437 | nan | 5.4 | 0.82 | 5 | 5.4 | 5.7 |
| fasting_p_glc | 30 | mean | 62 | nan | 5.41 | 0.99 | 5 | 5.4 | 5.8 |
| fasting_p_glc | 1825 | latest | 40 | nan | 5.41 | 0.8 | 5 | 5.4 | 5.8 |
| fasting_p_glc | 1460 | latest | 40 | nan | 5.4 | 0.82 | 5 | 5.4 | 5.8 |
| fasting_p_glc | 180 | latest | 39 | nan | 5.41 | 0.95 | 5 | 5.4 | 5.8 |
| fasting_p_glc | 1095 | minimum | 42 | nan | 5.32 | 0.84 | 5 | 5.3 | 5.7 |
| fasting_p_glc | 730 | minimum | 41 | nan | 5.34 | 0.87 | 5 | 5.3 | 5.7 |
| fasting_p_glc | 365 | minimum | 40 | nan | 5.37 | 0.92 | 5 | 5.3 | 5.7 |
| fasting_p_glc | 30 | minimum | 39 | nan | 5.41 | 0.99 | 5 | 5.4 | 5.8 |
| fasting_p_glc | 1825 | maximum | 39 | nan | 5.5 | 0.8 | 5.1 | 5.5 | 5.9 |
| fasting_p_glc | 1460 | maximum | 39 | nan | 5.49 | 0.82 | 5.1 | 5.5 | 5.9 |
| fasting_p_glc | 1095 | maximum | 39 | nan | 5.48 | 0.84 | 5.1 | 5.4 | 5.8 |
| fasting_p_glc | 365 | maximum | 40 | nan | 5.44 | 0.92 | 5.1 | 5.4 | 5.8 |
| fasting_p_glc | 730 | maximum | 40 | nan | 5.46 | 0.87 | 5.1 | 5.4 | 5.8 |
| fasting_p_glc | 180 | maximum | 39 | nan | 5.43 | 0.95 | 5.1 | 5.4 | 5.8 |
| fasting_p_glc | 365 | mean | 236 | nan | 5.41 | 0.92 | 5 | 5.4 | 5.8 |
| fasting_p_glc | 730 | mean | 335 | nan | 5.4 | 0.87 | 5 | 5.4 | 5.7 |
| fasting_p_glc | 1095 | mean | 402 | nan | 5.4 | 0.84 | 5 | 5.4 | 5.7 |
| fasting_p_glc | 1460 | minimum | 42 | nan | 5.31 | 0.82 | 5 | 5.3 | 5.7 |
| fasting_p_glc | 1825 | minimum | 42 | nan | 5.3 | 0.8 | 5 | 5.3 | 5.6 |
| fasting_p_glc | 1095 | latest | 40 | nan | 5.41 | 0.84 | 5 | 5.4 | 5.8 |
| fasting_p_glc | 730 | latest | 40 | nan | 5.4 | 0.87 | 5 | 5.4 | 5.8 |
| fasting_p_glc | 365 | latest | 40 | nan | 5.4 | 0.92 | 5 | 5.4 | 5.8 |

| | | | | | | | | | |
|---|---|---|---|---|---|---|---|---|---|
| fasting_p_glc | 30 | latest | 39 | nan | 5.41 | 0.99 | 5 | 5.4 | 5.8 |
| fasting_p_glc | 1825 | mean | 451 | nan | 5.4 | 0.8 | 5 | 5.4 | 5.7 |
| fasting_p_glc | 180 | mean | 159 | nan | 5.41 | 0.95 | 5 | 5.4 | 5.8 |
| fasting_p_glc | 30 | maximum | 39 | nan | 5.41 | 0.99 | 5 | 5.4 | 5.8 |
| fasting_p_glc | 180 | minimum | 39 | nan | 5.39 | 0.95 | 5 | 5.4 | 5.7 |
| gerd | 1460 | maximum | 2 | 0 | 0.01 | 0.99 | 0 | 0 | 0 |
| gerd | 1825 | latest | 2 | 0 | 0.01 | 0.99 | 0 | 0 | 0 |
| gerd | 180 | mean | 2 | 0 | 0 | 1 | 0 | 0 | 0 |
| gerd | 1460 | minimum | 2 | 0 | 0.01 | 0.99 | 0 | 0 | 0 |
| gerd | 1095 | minimum | 2 | 0 | 0.01 | 0.99 | 0 | 0 | 0 |
| gerd | 30 | maximum | 2 | 0 | 0 | 1 | 0 | 0 | 0 |
| gerd | 180 | maximum | 2 | 0 | 0 | 1 | 0 | 0 | 0 |
| gerd | 365 | maximum | 2 | 0 | 0 | 1 | 0 | 0 | 0 |
| gerd | 730 | maximum | 2 | 0 | 0.01 | 0.99 | 0 | 0 | 0 |
| gerd | 1095 | maximum | 2 | 0 | 0.01 | 0.99 | 0 | 0 | 0 |
| gerd | 1825 | maximum | 2 | 0 | 0.01 | 0.99 | 0 | 0 | 0 |
| gerd | 30 | minimum | 2 | 0 | 0 | 1 | 0 | 0 | 0 |
| gerd | 180 | minimum | 2 | 0 | 0 | 1 | 0 | 0 | 0 |
| gerd | 365 | mean | 2 | 0 | 0 | 1 | 0 | 0 | 0 |
| gerd | 1825 | minimum | 2 | 0 | 0.01 | 0.99 | 0 | 0 | 0 |
| gerd | 1460 | latest | 2 | 0 | 0.01 | 0.99 | 0 | 0 | 0 |
| gerd | 1095 | latest | 2 | 0 | 0.01 | 0.99 | 0 | 0 | 0 |
| gerd | 730 | latest | 2 | 0 | 0.01 | 0.99 | 0 | 0 | 0 |
| gerd | 365 | latest | 2 | 0 | 0 | 1 | 0 | 0 | 0 |
| gerd | 180 | latest | 2 | 0 | 0 | 1 | 0 | 0 | 0 |
| gerd | 30 | latest | 2 | 0 | 0 | 1 | 0 | 0 | 0 |
| gerd | 1825 | mean | 2 | 0 | 0.01 | 0.99 | 0 | 0 | 0 |
| gerd | 1460 | mean | 2 | 0 | 0.01 | 0.99 | 0 | 0 | 0 |

| | | | | | | | | | |
|---|---|---|---|---|---|---|---|---|---|
| gerd | 1095 | mean | 2 | 0 | 0.01 | 0.99 | 0 | 0 | 0 |
| gerd | 730 | mean | 2 | 0 | 0.01 | 0.99 | 0 | 0 | 0 |
| gerd | 365 | minimum | 2 | 0 | 0 | 1 | 0 | 0 | 0 |
| gerd | 730 | minimum | 2 | 0 | 0.01 | 0.99 | 0 | 0 | 0 |
| gerd | 30 | mean | 2 | 0 | 0 | 1 | 0 | 0 | 0 |
| gerd_drugs | 1095 | latest | 2 | 0 | 0.15 | 0.85 | 0 | 0 | 0 |
| gerd_drugs | 30 | mean | 2 | 0 | 0.02 | 0.98 | 0 | 0 | 0 |
| gerd_drugs | 365 | maximum | 2 | 0 | 0.09 | 0.91 | 0 | 0 | 0 |
| gerd_drugs | 1825 | maximum | 2 | 0 | 0.18 | 0.82 | 0 | 0 | 0 |
| gerd_drugs | 1460 | maximum | 2 | 0 | 0.17 | 0.83 | 0 | 0 | 0 |
| gerd_drugs | 730 | maximum | 2 | 0 | 0.13 | 0.87 | 0 | 0 | 0 |
| gerd_drugs | 1095 | maximum | 2 | 0 | 0.15 | 0.85 | 0 | 0 | 0 |
| gerd_drugs | 30 | minimum | 2 | 0 | 0.02 | 0.98 | 0 | 0 | 0 |
| gerd_drugs | 1825 | latest | 2 | 0 | 0.18 | 0.82 | 0 | 0 | 0 |
| gerd_drugs | 30 | maximum | 2 | 0 | 0.02 | 0.98 | 0 | 0 | 0 |
| gerd_drugs | 730 | mean | 2 | 0 | 0.13 | 0.87 | 0 | 0 | 0 |
| gerd_drugs | 180 | minimum | 2 | 0 | 0.06 | 0.94 | 0 | 0 | 0 |
| gerd_drugs | 365 | minimum | 2 | 0 | 0.09 | 0.91 | 0 | 0 | 0 |
| gerd_drugs | 730 | minimum | 2 | 0 | 0.13 | 0.87 | 0 | 0 | 0 |
| gerd_drugs | 180 | maximum | 2 | 0 | 0.06 | 0.94 | 0 | 0 | 0 |
| gerd_drugs | 1095 | minimum | 2 | 0 | 0.15 | 0.85 | 0 | 0 | 0 |
| gerd_drugs | 1825 | minimum | 2 | 0 | 0.18 | 0.82 | 0 | 0 | 0 |
| gerd_drugs | 180 | mean | 2 | 0 | 0.06 | 0.94 | 0 | 0 | 0 |
| gerd_drugs | 1460 | latest | 2 | 0 | 0.17 | 0.83 | 0 | 0 | 0 |
| gerd_drugs | 365 | mean | 2 | 0 | 0.09 | 0.91 | 0 | 0 | 0 |
| gerd_drugs | 1095 | mean | 2 | 0 | 0.15 | 0.85 | 0 | 0 | 0 |
| gerd_drugs | 1460 | mean | 2 | 0 | 0.17 | 0.83 | 0 | 0 | 0 |
| gerd_drugs | 1825 | mean | 2 | 0 | 0.18 | 0.82 | 0 | 0 | 0 |

| | | | | | | | | | |
|---|---|---|---|---|---|---|---|---|---|
| gerd_drugs | 30 | latest | 2 | 0 | 0.02 | 0.98 | 0 | 0 | 0 |
| gerd_drugs | 180 | latest | 2 | 0 | 0.06 | 0.94 | 0 | 0 | 0 |
| gerd_drugs | 365 | latest | 2 | 0 | 0.09 | 0.91 | 0 | 0 | 0 |
| gerd_drugs | 730 | latest | 2 | 0 | 0.13 | 0.87 | 0 | 0 | 0 |
| gerd_drugs | 1460 | minimum | 2 | 0 | 0.17 | 0.83 | 0 | 0 | 0 |
| hba1c | 180 | maximum | 57 | nan | 34.18 | 0.53 | 32 | 34 | 37 |
| hba1c | 30 | maximum | 53 | nan | 33.99 | 0.89 | 32 | 34 | 36 |
| hba1c | 365 | maximum | 56 | nan | 34.36 | 0.34 | 32 | 34 | 37 |
| hba1c | 1095 | minimum | 61 | nan | 32.73 | 0.18 | 31 | 33 | 35 |
| hba1c | 1825 | minimum | 64 | nan | 32.46 | 0.16 | 30 | 33 | 35 |
| hba1c | 1095 | maximum | 48 | nan | 34.91 | 0.18 | 32 | 35 | 37 |
| hba1c | 30 | latest | 54 | nan | 33.96 | 0.89 | 31 | 34 | 36 |
| hba1c | 365 | latest | 62 | nan | 33.9 | 0.34 | 31 | 34 | 36 |
| hba1c | 30 | mean | 135 | nan | 33.97 | 0.89 | 31 | 34 | 36 |
| hba1c | 1460 | minimum | 64 | nan | 32.57 | 0.17 | 30 | 33 | 35 |
| hba1c | 1825 | mean | 2829 | nan | 33.83 | 0.16 | 31.6 | 33.8 | 36 |
| hba1c | 1460 | mean | 2461 | nan | 33.83 | 0.17 | 31.6 | 33.8 | 36 |
| hba1c | 1095 | mean | 2003 | nan | 33.84 | 0.18 | 31.5 | 34 | 36 |
| hba1c | 730 | mean | 1437 | nan | 33.85 | 0.22 | 31.5 | 34 | 36 |
| hba1c | 180 | mean | 456 | nan | 33.95 | 0.53 | 31.5 | 34 | 36 |
| hba1c | 365 | mean | 785 | nan | 33.9 | 0.34 | 31.5 | 34 | 36 |
| hba1c | 180 | latest | 60 | nan | 33.95 | 0.53 | 31 | 34 | 36 |
| hba1c | 1825 | latest | 65 | nan | 33.84 | 0.16 | 31 | 34 | 36 |
| hba1c | 1460 | maximum | 47 | nan | 35.07 | 0.17 | 33 | 35 | 37 |
| hba1c | 730 | minimum | 60 | nan | 33 | 0.22 | 31 | 33 | 35 |
| hba1c | 365 | minimum | 62 | nan | 33.43 | 0.34 | 31 | 33 | 36 |
| hba1c | 180 | minimum | 60 | nan | 33.73 | 0.53 | 31 | 34 | 36 |
| hba1c | 30 | minimum | 54 | nan | 33.94 | 0.89 | 31 | 34 | 36 |

| | | | | | | | | | |
|---|---|---|---|---|---|---|---|---|---|
| hba1c | 1825 | maximum | 45 | nan | 35.16 | 0.16 | 33 | 35 | 38 |
| hba1c | 1460 | latest | 65 | nan | 33.84 | 0.17 | 31 | 34 | 36 |
| hba1c | 1095 | latest | 64 | nan | 33.85 | 0.18 | 31 | 34 | 36 |
| hba1c | 730 | latest | 63 | nan | 33.86 | 0.22 | 31 | 34 | 36 |
| hba1c | 730 | maximum | 51 | nan | 34.68 | 0.22 | 32 | 35 | 37 |
| hdl | 730 | maximum | 273 | nan | 1.47 | 0.23 | 1.2 | 1.4 | 1.7 |
| hdl | 30 | maximum | 234 | nan | 1.37 | 0.89 | 1.1 | 1.3 | 1.6 |
| hdl | 365 | maximum | 265 | nan | 1.43 | 0.34 | 1.1 | 1.4 | 1.7 |
| hdl | 180 | maximum | 257 | nan | 1.4 | 0.53 | 1.1 | 1.3 | 1.6 |
| hdl | 1095 | latest | 278 | nan | 1.38 | 0.19 | 1.1 | 1.3 | 1.6 |
| hdl | 1460 | latest | 279 | nan | 1.38 | 0.17 | 1.1 | 1.3 | 1.6 |
| hdl | 30 | latest | 234 | nan | 1.37 | 0.89 | 1.1 | 1.3 | 1.6 |
| hdl | 365 | latest | 270 | nan | 1.37 | 0.34 | 1.1 | 1.3 | 1.6 |
| hdl | 1095 | minimum | 258 | nan | 1.28 | 0.19 | 1 | 1.2 | 1.5 |
| hdl | 730 | latest | 275 | nan | 1.38 | 0.23 | 1.1 | 1.3 | 1.6 |
| hdl | 1825 | minimum | 261 | nan | 1.26 | 0.16 | 1 | 1.2 | 1.5 |
| hdl | 30 | mean | 440 | nan | 1.37 | 0.89 | 1.1 | 1.3 | 1.6 |
| hdl | 180 | mean | 1415 | nan | 1.37 | 0.53 | 1.1 | 1.3 | 1.6 |
| hdl | 1460 | minimum | 257 | nan | 1.27 | 0.17 | 1 | 1.2 | 1.5 |
| hdl | 730 | mean | 4382 | nan | 1.38 | 0.23 | 1.1 | 1.3 | 1.6 |
| hdl | 1460 | maximum | 280 | nan | 1.52 | 0.17 | 1.2 | 1.4 | 1.8 |
| hdl | 1095 | mean | 6109 | nan | 1.39 | 0.19 | 1.1 | 1.3 | 1.6 |
| hdl | 1825 | mean | 8513 | nan | 1.39 | 0.16 | 1.1 | 1.3 | 1.6 |
| hdl | 180 | latest | 262 | nan | 1.37 | 0.53 | 1.1 | 1.3 | 1.6 |
| hdl | 365 | mean | 2451 | nan | 1.38 | 0.34 | 1.1 | 1.3 | 1.6 |
| hdl | 730 | minimum | 262 | nan | 1.3 | 0.23 | 1 | 1.2 | 1.5 |
| hdl | 365 | minimum | 265 | nan | 1.33 | 0.34 | 1 | 1.3 | 1.5 |
| hdl | 1095 | maximum | 280 | nan | 1.5 | 0.19 | 1.2 | 1.4 | 1.7 |

| | | | | | | | | | |
|---|---|---|---|---|---|---|---|---|---|
| hdl | 30 | minimum | 233 | nan | 1.37 | 0.89 | 1.1 | 1.3 | 1.6 |
| hdl | 1825 | maximum | 284 | nan | 1.53 | 0.16 | 1.2 | 1.5 | 1.8 |
| hdl | 180 | minimum | 259 | nan | 1.35 | 0.53 | 1.1 | 1.3 | 1.6 |
| hdl | 1460 | mean | 7421 | nan | 1.39 | 0.17 | 1.1 | 1.3 | 1.6 |
| hdl | 1825 | latest | 280 | nan | 1.38 | 0.16 | 1.1 | 1.3 | 1.6 |
| height_in_cm | 730 | latest | 785 | nan | 170.45 | 0.27 | 165 | 171 | 178 |
| height_in_cm | 1095 | latest | 798 | nan | 170.45 | 0.22 | 165 | 171 | 178 |
| height_in_cm | 1460 | latest | 803 | nan | 170.47 | 0.2 | 165 | 171 | 178 |
| height_in_cm | 365 | latest | 764 | nan | 170.49 | 0.39 | 165 | 171 | 178 |
| height_in_cm | 180 | latest | 719 | nan | 170.44 | 0.55 | 165 | 171 | 178 |
| height_in_cm | 30 | latest | 550 | nan | 169.86 | 0.86 | 165 | 170 | 177 |
| height_in_cm | 1825 | latest | 808 | nan | 170.49 | 0.18 | 165 | 171 | 178 |
| hyperlipidemia | 365 | maximum | 2 | 0 | 0 | 1 | 0 | 0 | 0 |
| hyperlipidemia | 180 | maximum | 2 | 0 | 0 | 1 | 0 | 0 | 0 |
| hyperlipidemia | 30 | maximum | 2 | 0 | 0 | 1 | 0 | 0 | 0 |
| hyperlipidemia | 730 | minimum | 2 | 0 | 0 | 1 | 0 | 0 | 0 |
| hyperlipidemia | 30 | latest | 2 | 0 | 0 | 1 | 0 | 0 | 0 |
| hyperlipidemia | 365 | minimum | 2 | 0 | 0 | 1 | 0 | 0 | 0 |
| hyperlipidemia | 180 | latest | 2 | 0 | 0 | 1 | 0 | 0 | 0 |
| hyperlipidemia | 730 | maximum | 2 | 0 | 0 | 1 | 0 | 0 | 0 |
| hyperlipidemia | 730 | latest | 2 | 0 | 0 | 1 | 0 | 0 | 0 |
| hyperlipidemia | 1095 | latest | 2 | 0 | 0 | 1 | 0 | 0 | 0 |
| hyperlipidemia | 1460 | latest | 2 | 0 | 0 | 1 | 0 | 0 | 0 |
| hyperlipidemia | 1825 | latest | 2 | 0 | 0 | 1 | 0 | 0 | 0 |
| hyperlipidemia | 365 | latest | 2 | 0 | 0 | 1 | 0 | 0 | 0 |
| hyperlipidemia | 1825 | mean | 2 | 0 | 0 | 1 | 0 | 0 | 0 |
| hyperlipidemia | 365 | mean | 2 | 0 | 0 | 1 | 0 | 0 | 0 |
| hyperlipidemia | 1825 | maximum | 2 | 0 | 0 | 1 | 0 | 0 | 0 |

| | | | | | | | | | |
|---|---|---|---|---|---|---|---|---|---|
| hyperlipidemia | 1460 | maximum | 2 | 0 | 0 | 1 | 0 | 0 | 0 |
| hyperlipidemia | 1460 | mean | 2 | 0 | 0 | 1 | 0 | 0 | 0 |
| hyperlipidemia | 1095 | minimum | 2 | 0 | 0 | 1 | 0 | 0 | 0 |
| hyperlipidemia | 1095 | mean | 2 | 0 | 0 | 1 | 0 | 0 | 0 |
| hyperlipidemia | 180 | mean | 2 | 0 | 0 | 1 | 0 | 0 | 0 |
| hyperlipidemia | 730 | mean | 2 | 0 | 0 | 1 | 0 | 0 | 0 |
| hyperlipidemia | 1825 | minimum | 2 | 0 | 0 | 1 | 0 | 0 | 0 |
| hyperlipidemia | 1460 | minimum | 2 | 0 | 0 | 1 | 0 | 0 | 0 |
| hyperlipidemia | 1095 | maximum | 2 | 0 | 0 | 1 | 0 | 0 | 0 |
| hyperlipidemia | 180 | minimum | 2 | 0 | 0 | 1 | 0 | 0 | 0 |
| hyperlipidemia | 30 | minimum | 2 | 0 | 0 | 1 | 0 | 0 | 0 |
| hyperlipidemia | 30 | mean | 2 | 0 | 0 | 1 | 0 | 0 | 0 |
| lamotrigine | 180 | latest | 2 | 0 | 0.03 | 0.97 | 0 | 0 | 0 |
| lamotrigine | 1095 | mean | 2 | 0 | 0.06 | 0.94 | 0 | 0 | 0 |
| lamotrigine | 1825 | mean | 2 | 0 | 0.07 | 0.93 | 0 | 0 | 0 |
| lamotrigine | 30 | latest | 2 | 0 | 0.01 | 0.99 | 0 | 0 | 0 |
| lamotrigine | 365 | latest | 2 | 0 | 0.04 | 0.96 | 0 | 0 | 0 |
| lamotrigine | 730 | mean | 2 | 0 | 0.05 | 0.95 | 0 | 0 | 0 |
| lamotrigine | 1095 | latest | 2 | 0 | 0.06 | 0.94 | 0 | 0 | 0 |
| lamotrigine | 1460 | latest | 2 | 0 | 0.07 | 0.93 | 0 | 0 | 0 |
| lamotrigine | 1825 | latest | 2 | 0 | 0.07 | 0.93 | 0 | 0 | 0 |
| lamotrigine | 30 | maximum | 2 | 0 | 0.01 | 0.99 | 0 | 0 | 0 |
| lamotrigine | 180 | maximum | 2 | 0 | 0.03 | 0.97 | 0 | 0 | 0 |
| lamotrigine | 180 | mean | 2 | 0 | 0.03 | 0.97 | 0 | 0 | 0 |
| lamotrigine | 730 | latest | 2 | 0 | 0.05 | 0.95 | 0 | 0 | 0 |
| lamotrigine | 1460 | mean | 2 | 0 | 0.07 | 0.93 | 0 | 0 | 0 |
| lamotrigine | 365 | maximum | 2 | 0 | 0.04 | 0.96 | 0 | 0 | 0 |
| lamotrigine | 1460 | maximum | 2 | 0 | 0.07 | 0.93 | 0 | 0 | 0 |

| | | | | | | | | | |
|---|---|---|---|---|---|---|---|---|---|
| lamotrigine | 30 | mean | 2 | 0 | 0.01 | 0.99 | 0 | 0 | 0 |
| lamotrigine | 1825 | minimum | 2 | 0 | 0.07 | 0.93 | 0 | 0 | 0 |
| lamotrigine | 1825 | maximum | 2 | 0 | 0.07 | 0.93 | 0 | 0 | 0 |
| lamotrigine | 1095 | minimum | 2 | 0 | 0.06 | 0.94 | 0 | 0 | 0 |
| lamotrigine | 1460 | minimum | 2 | 0 | 0.07 | 0.93 | 0 | 0 | 0 |
| lamotrigine | 1095 | maximum | 2 | 0 | 0.06 | 0.94 | 0 | 0 | 0 |
| lamotrigine | 730 | minimum | 2 | 0 | 0.05 | 0.95 | 0 | 0 | 0 |
| lamotrigine | 365 | minimum | 2 | 0 | 0.04 | 0.96 | 0 | 0 | 0 |
| lamotrigine | 365 | mean | 2 | 0 | 0.04 | 0.96 | 0 | 0 | 0 |
| lamotrigine | 180 | minimum | 2 | 0 | 0.03 | 0.97 | 0 | 0 | 0 |
| lamotrigine | 30 | minimum | 2 | 0 | 0.01 | 0.99 | 0 | 0 | 0 |
| lamotrigine | 730 | maximum | 2 | 0 | 0.05 | 0.95 | 0 | 0 | 0 |
| ldl | 1095 | latest | 87 | nan | 2.84 | 0.24 | 2.2 | 2.8 | 3.4 |
| ldl | 1460 | latest | 87 | nan | 2.84 | 0.22 | 2.2 | 2.8 | 3.4 |
| ldl | 1825 | latest | 87 | nan | 2.84 | 0.21 | 2.2 | 2.8 | 3.4 |
| ldl | 1825 | minimum | 79 | nan | 2.53 | 0.21 | 1.9 | 2.5 | 3 |
| ldl | 30 | maximum | 82 | nan | 2.87 | 0.9 | 2.2 | 2.8 | 3.4 |
| ldl | 180 | maximum | 86 | nan | 2.93 | 0.57 | 2.3 | 2.8 | 3.5 |
| ldl | 1460 | maximum | 95 | nan | 3.18 | 0.22 | 2.5 | 3.1 | 3.8 |
| ldl | 1095 | maximum | 94 | nan | 3.14 | 0.24 | 2.4 | 3 | 3.7 |
| ldl | 1825 | maximum | 93 | nan | 3.2 | 0.21 | 2.5 | 3.1 | 3.8 |
| ldl | 180 | minimum | 84 | nan | 2.8 | 0.57 | 2.2 | 2.7 | 3.4 |
| ldl | 1460 | minimum | 82 | nan | 2.55 | 0.22 | 2 | 2.5 | 3.1 |
| ldl | 730 | latest | 87 | nan | 2.84 | 0.28 | 2.2 | 2.8 | 3.4 |
| ldl | 730 | maximum | 93 | nan | 3.08 | 0.28 | 2.4 | 3 | 3.7 |
| ldl | 365 | latest | 87 | nan | 2.85 | 0.4 | 2.2 | 2.8 | 3.4 |
| ldl | 365 | maximum | 91 | nan | 2.99 | 0.4 | 2.3 | 2.9 | 3.6 |
| ldl | 30 | latest | 82 | nan | 2.86 | 0.9 | 2.2 | 2.8 | 3.4 |

| | | | | | | | | | |
|---|---|---|---|---|---|---|---|---|---|
| ldl | 730 | minimum | 86 | nan | 2.64 | 0.28 | 2 | 2.6 | 3.2 |
| ldl | 1095 | minimum | 85 | nan | 2.58 | 0.24 | 2 | 2.5 | 3.1 |
| ldl | 365 | minimum | 87 | nan | 2.73 | 0.4 | 2.1 | 2.7 | 3.3 |
| ldl | 30 | minimum | 83 | nan | 2.86 | 0.9 | 2.2 | 2.8 | 3.4 |
| ldl | 30 | mean | 247 | nan | 2.86 | 0.9 | 2.2 | 2.8 | 3.4 |
| ldl | 180 | latest | 85 | nan | 2.86 | 0.57 | 2.2 | 2.8 | 3.4 |
| ldl | 365 | mean | 1403 | nan | 2.86 | 0.4 | 2.2 | 2.8 | 3.4 |
| ldl | 730 | mean | 2329 | nan | 2.86 | 0.28 | 2.2 | 2.8 | 3.4 |
| ldl | 1095 | mean | 3124 | nan | 2.86 | 0.24 | 2.2 | 2.8 | 3.4 |
| ldl | 1460 | mean | 3751 | nan | 2.86 | 0.22 | 2.3 | 2.8 | 3.4 |
| ldl | 1825 | mean | 4228 | nan | 2.86 | 0.21 | 2.3 | 2.8 | 3.4 |
| ldl | 180 | mean | 798 | nan | 2.87 | 0.57 | 2.2 | 2.8 | 3.4 |
| lithium | 180 | minimum | 2 | 0 | 0.02 | 0.98 | 0 | 0 | 0 |
| lithium | 1460 | minimum | 2 | 0 | 0.04 | 0.96 | 0 | 0 | 0 |
| lithium | 1095 | minimum | 2 | 0 | 0.03 | 0.97 | 0 | 0 | 0 |
| lithium | 730 | minimum | 2 | 0 | 0.03 | 0.97 | 0 | 0 | 0 |
| lithium | 365 | minimum | 2 | 0 | 0.02 | 0.98 | 0 | 0 | 0 |
| lithium | 30 | minimum | 2 | 0 | 0.01 | 0.99 | 0 | 0 | 0 |
| lithium | 180 | maximum | 2 | 0 | 0.02 | 0.98 | 0 | 0 | 0 |
| lithium | 1460 | maximum | 2 | 0 | 0.04 | 0.96 | 0 | 0 | 0 |
| lithium | 1095 | maximum | 2 | 0 | 0.03 | 0.97 | 0 | 0 | 0 |
| lithium | 730 | maximum | 2 | 0 | 0.03 | 0.97 | 0 | 0 | 0 |
| lithium | 30 | mean | 2 | 0 | 0.01 | 0.99 | 0 | 0 | 0 |
| lithium | 1825 | minimum | 2 | 0 | 0.04 | 0.96 | 0 | 0 | 0 |
| lithium | 1825 | maximum | 2 | 0 | 0.04 | 0.96 | 0 | 0 | 0 |
| lithium | 180 | mean | 2 | 0 | 0.02 | 0.98 | 0 | 0 | 0 |
| lithium | 30 | maximum | 2 | 0 | 0.01 | 0.99 | 0 | 0 | 0 |
| lithium | 365 | mean | 2 | 0 | 0.02 | 0.98 | 0 | 0 | 0 |

| | | | | | | | | | |
|---|---|---|---|---|---|---|---|---|---|
| lithium | 365 | maximum | 2 | 0 | 0.02 | 0.98 | 0 | 0 | 0 |
| lithium | 1825 | latest | 2 | 0 | 0.04 | 0.96 | 0 | 0 | 0 |
| lithium | 1460 | latest | 2 | 0 | 0.04 | 0.96 | 0 | 0 | 0 |
| lithium | 1095 | latest | 2 | 0 | 0.03 | 0.97 | 0 | 0 | 0 |
| lithium | 730 | latest | 2 | 0 | 0.03 | 0.97 | 0 | 0 | 0 |
| lithium | 1825 | mean | 2 | 0 | 0.04 | 0.96 | 0 | 0 | 0 |
| lithium | 180 | latest | 2 | 0 | 0.02 | 0.98 | 0 | 0 | 0 |
| lithium | 30 | latest | 2 | 0 | 0.01 | 0.99 | 0 | 0 | 0 |
| lithium | 1460 | mean | 2 | 0 | 0.04 | 0.96 | 0 | 0 | 0 |
| lithium | 1095 | mean | 2 | 0 | 0.03 | 0.97 | 0 | 0 | 0 |
| lithium | 730 | mean | 2 | 0 | 0.03 | 0.97 | 0 | 0 | 0 |
| lithium | 365 | latest | 2 | 0 | 0.02 | 0.98 | 0 | 0 | 0 |
| ogtt | 1825 | minimum | 78 | nan | 6.54 | 0.99 | 5.6 | 6.4 | 7.4 |
| ogtt | 730 | latest | 76 | nan | 6.86 | 0.99 | 5.8 | 6.7 | 7.8 |
| ogtt | 365 | latest | 70 | nan | 6.79 | 1 | 5.8 | 6.7 | 7.8 |
| ogtt | 180 | latest | 70 | nan | 6.72 | 1 | 5.8 | 6.7 | 7.8 |
| ogtt | 1095 | latest | 78 | nan | 6.88 | 0.99 | 5.9 | 6.7 | 7.8 |
| ogtt | 1460 | mean | 167 | nan | 6.78 | 0.99 | 5.8 | 6.7 | 7.7 |
| ogtt | 30 | latest | 54 | nan | 6.78 | 1 | 5.9 | 6.6 | 7.8 |
| ogtt | 1095 | mean | 159 | nan | 6.84 | 0.99 | 5.9 | 6.7 | 7.8 |
| ogtt | 730 | mean | 138 | nan | 6.81 | 0.99 | 5.8 | 6.7 | 7.8 |
| ogtt | 365 | mean | 108 | nan | 6.74 | 1 | 5.8 | 6.7 | 7.8 |
| ogtt | 30 | mean | 56 | nan | 6.79 | 1 | 5.9 | 6.6 | 7.8 |
| ogtt | 1825 | mean | 177 | nan | 6.74 | 0.99 | 5.8 | 6.6 | 7.6 |
| ogtt | 1460 | maximum | 77 | nan | 6.96 | 0.99 | 5.9 | 6.8 | 7.9 |
| ogtt | 180 | mean | 96 | nan | 6.68 | 1 | 5.8 | 6.6 | 7.7 |
| ogtt | 1095 | minimum | 78 | nan | 6.68 | 0.99 | 5.7 | 6.6 | 7.6 |
| ogtt | 365 | minimum | 69 | nan | 6.64 | 1 | 5.7 | 6.6 | 7.6 |

| | | | | | | | | | |
|---|---|---|---|---|---|---|---|---|---|
| ogtt | 1460 | latest | 78 | nan | 6.81 | 0.99 | 5.8 | 6.7 | 7.7 |
| ogtt | 1825 | latest | 78 | nan | 6.76 | 0.99 | 5.7 | 6.6 | 7.7 |
| ogtt | 30 | maximum | 54 | nan | 6.79 | 1 | 5.9 | 6.6 | 7.8 |
| ogtt | 180 | maximum | 70 | nan | 6.75 | 1 | 5.8 | 6.7 | 7.8 |
| ogtt | 365 | maximum | 70 | nan | 6.84 | 1 | 5.9 | 6.7 | 7.8 |
| ogtt | 1460 | minimum | 78 | nan | 6.59 | 0.99 | 5.6 | 6.5 | 7.5 |
| ogtt | 1095 | maximum | 77 | nan | 7.01 | 0.99 | 5.9 | 6.8 | 7.9 |
| ogtt | 1825 | maximum | 77 | nan | 6.94 | 0.99 | 5.9 | 6.8 | 7.8 |
| ogtt | 30 | minimum | 54 | nan | 6.78 | 1 | 5.9 | 6.6 | 7.8 |
| ogtt | 180 | minimum | 69 | nan | 6.61 | 1 | 5.7 | 6.5 | 7.6 |
| ogtt | 730 | minimum | 76 | nan | 6.7 | 0.99 | 5.7 | 6.6 | 7.7 |
| ogtt | 730 | maximum | 76 | nan | 6.93 | 0.99 | 5.8 | 6.7 | 7.9 |
| polycystic_ovarian_syndrome | 1095 | maximum | 2 | 0 | 0 | 1 | 0 | 0 | 0 |
| polycystic_ovarian_syndrome | 180 | latest | 2 | 0 | 0 | 1 | 0 | 0 | 0 |
| polycystic_ovarian_syndrome | 365 | maximum | 2 | 0 | 0 | 1 | 0 | 0 | 0 |
| polycystic_ovarian_syndrome | 180 | maximum | 2 | 0 | 0 | 1 | 0 | 0 | 0 |
| polycystic_ovarian_syndrome | 30 | maximum | 2 | 0 | 0 | 1 | 0 | 0 | 0 |
| polycystic_ovarian_syndrome | 1095 | latest | 2 | 0 | 0 | 1 | 0 | 0 | 0 |
| polycystic_ovarian_syndrome | 730 | latest | 2 | 0 | 0 | 1 | 0 | 0 | 0 |
| polycystic_ovarian_syndrome | 1825 | minimum | 2 | 0 | 0.01 | 0.99 | 0 | 0 | 0 |
| polycystic_ovarian_syndrome | 1460 | latest | 2 | 0 | 0 | 1 | 0 | 0 | 0 |
| polycystic_ovarian_syndrome | 1825 | latest | 2 | 0 | 0.01 | 0.99 | 0 | 0 | 0 |
| polycystic_ovarian_syndrome | 730 | maximum | 2 | 0 | 0 | 1 | 0 | 0 | 0 |
| polycystic_ovarian_syndrome | 365 | latest | 2 | 0 | 0 | 1 | 0 | 0 | 0 |
| polycystic_ovarian_syndrome | 1825 | mean | 2 | 0 | 0.01 | 0.99 | 0 | 0 | 0 |
| polycystic_ovarian_syndrome | 30 | latest | 2 | 0 | 0 | 1 | 0 | 0 | 0 |
| polycystic_ovarian_syndrome | 1460 | mean | 2 | 0 | 0 | 1 | 0 | 0 | 0 |
| polycystic_ovarian_syndrome | 1825 | maximum | 2 | 0 | 0.01 | 0.99 | 0 | 0 | 0 |

| | | | | | | | | | |
|---|---|---|---|---|---|---|---|---|---|
| polycystic_ovarian_syndrome | 180 | minimum | 2 | 0 | 0 | 1 | 0 | 0 | 0 |
| polycystic_ovarian_syndrome | 365 | minimum | 2 | 0 | 0 | 1 | 0 | 0 | 0 |
| polycystic_ovarian_syndrome | 730 | minimum | 2 | 0 | 0 | 1 | 0 | 0 | 0 |
| polycystic_ovarian_syndrome | 1095 | minimum | 2 | 0 | 0 | 1 | 0 | 0 | 0 |
| polycystic_ovarian_syndrome | 30 | minimum | 2 | 0 | 0 | 1 | 0 | 0 | 0 |
| polycystic_ovarian_syndrome | 1460 | maximum | 2 | 0 | 0 | 1 | 0 | 0 | 0 |
| polycystic_ovarian_syndrome | 1095 | mean | 2 | 0 | 0 | 1 | 0 | 0 | 0 |
| polycystic_ovarian_syndrome | 30 | mean | 2 | 0 | 0 | 1 | 0 | 0 | 0 |
| polycystic_ovarian_syndrome | 180 | mean | 2 | 0 | 0 | 1 | 0 | 0 | 0 |
| polycystic_ovarian_syndrome | 365 | mean | 2 | 0 | 0 | 1 | 0 | 0 | 0 |
| polycystic_ovarian_syndrome | 730 | mean | 2 | 0 | 0 | 1 | 0 | 0 | 0 |
| polycystic_ovarian_syndrome | 1460 | minimum | 2 | 0 | 0 | 1 | 0 | 0 | 0 |
| pregabaline | 1095 | mean | 2 | 0 | 0.05 | 0.95 | 0 | 0 | 0 |
| pregabaline | 730 | mean | 2 | 0 | 0.04 | 0.96 | 0 | 0 | 0 |
| pregabaline | 365 | mean | 2 | 0 | 0.03 | 0.97 | 0 | 0 | 0 |
| pregabaline | 180 | mean | 2 | 0 | 0.02 | 0.98 | 0 | 0 | 0 |
| pregabaline | 30 | mean | 2 | 0 | 0.01 | 0.99 | 0 | 0 | 0 |
| pregabaline | 1825 | minimum | 2 | 0 | 0.06 | 0.94 | 0 | 0 | 0 |
| pregabaline | 730 | minimum | 2 | 0 | 0.04 | 0.96 | 0 | 0 | 0 |
| pregabaline | 1095 | minimum | 2 | 0 | 0.05 | 0.95 | 0 | 0 | 0 |
| pregabaline | 180 | maximum | 2 | 0 | 0.02 | 0.98 | 0 | 0 | 0 |
| pregabaline | 30 | maximum | 2 | 0 | 0.01 | 0.99 | 0 | 0 | 0 |
| pregabaline | 365 | minimum | 2 | 0 | 0.03 | 0.97 | 0 | 0 | 0 |
| pregabaline | 1460 | mean | 2 | 0 | 0.06 | 0.94 | 0 | 0 | 0 |
| pregabaline | 1460 | minimum | 2 | 0 | 0.06 | 0.94 | 0 | 0 | 0 |
| pregabaline | 180 | minimum | 2 | 0 | 0.02 | 0.98 | 0 | 0 | 0 |
| pregabaline | 1825 | mean | 2 | 0 | 0.06 | 0.94 | 0 | 0 | 0 |
| pregabaline | 180 | latest | 2 | 0 | 0.02 | 0.98 | 0 | 0 | 0 |

| | | | | | | | | | |
|---|---|---|---|---|---|---|---|---|---|
| pregabaline | 30 | latest | 2 | 0 | 0.01 | 0.99 | 0 | 0 | 0 |
| pregabaline | 365 | maximum | 2 | 0 | 0.03 | 0.97 | 0 | 0 | 0 |
| pregabaline | 730 | maximum | 2 | 0 | 0.04 | 0.96 | 0 | 0 | 0 |
| pregabaline | 1095 | maximum | 2 | 0 | 0.05 | 0.95 | 0 | 0 | 0 |
| pregabaline | 1825 | maximum | 2 | 0 | 0.06 | 0.94 | 0 | 0 | 0 |
| pregabaline | 1460 | latest | 2 | 0 | 0.06 | 0.94 | 0 | 0 | 0 |
| pregabaline | 1095 | latest | 2 | 0 | 0.05 | 0.95 | 0 | 0 | 0 |
| pregabaline | 1460 | maximum | 2 | 0 | 0.06 | 0.94 | 0 | 0 | 0 |
| pregabaline | 30 | minimum | 2 | 0 | 0.01 | 0.99 | 0 | 0 | 0 |
| pregabaline | 730 | latest | 2 | 0 | 0.04 | 0.96 | 0 | 0 | 0 |
| pregabaline | 365 | latest | 2 | 0 | 0.03 | 0.97 | 0 | 0 | 0 |
| pregabaline | 1825 | latest | 2 | 0 | 0.06 | 0.94 | 0 | 0 | 0 |
| scheduled_glc | 180 | latest | 106 | nan | 6.06 | 0.99 | 5.1 | 5.7 | 6.6 |
| scheduled_glc | 365 | latest | 114 | nan | 5.99 | 0.98 | 5.1 | 5.6 | 6.5 |
| scheduled_glc | 730 | latest | 119 | nan | 5.94 | 0.96 | 5 | 5.6 | 6.4 |
| scheduled_glc | 1095 | latest | 121 | nan | 5.92 | 0.95 | 5 | 5.6 | 6.4 |
| scheduled_glc | 1460 | latest | 122 | nan | 5.92 | 0.94 | 5 | 5.6 | 6.5 |
| scheduled_glc | 365 | maximum | 156 | nan | 6.51 | 0.98 | 5.2 | 5.8 | 7.1 |
| scheduled_glc | 180 | maximum | 138 | nan | 6.57 | 0.99 | 5.2 | 5.9 | 7.2 |
| scheduled_glc | 1825 | minimum | 116 | nan | 5.52 | 0.93 | 4.8 | 5.3 | 6.1 |
| scheduled_glc | 1095 | maximum | 166 | nan | 6.44 | 0.95 | 5.2 | 5.8 | 7.1 |
| scheduled_glc | 1460 | maximum | 167 | nan | 6.43 | 0.94 | 5.2 | 5.8 | 7.1 |
| scheduled_glc | 730 | maximum | 163 | nan | 6.45 | 0.96 | 5.2 | 5.8 | 7.1 |
| scheduled_glc | 1825 | latest | 122 | nan | 5.92 | 0.93 | 5 | 5.6 | 6.5 |
| scheduled_glc | 30 | latest | 95 | nan | 6.11 | 1 | 5.1 | 5.7 | 6.7 |
| scheduled_glc | 1825 | mean | 927 | nan | 5.88 | 0.93 | 5.1 | 5.6 | 6.4 |
| scheduled_glc | 1095 | mean | 892 | nan | 5.89 | 0.95 | 5.1 | 5.6 | 6.4 |
| scheduled_glc | 30 | maximum | 115 | nan | 6.54 | 1 | 5.2 | 5.9 | 7.2 |

| | | | | | | | | | |
|---|---|---|---|---|---|---|---|---|---|
| scheduled_glc | 1825 | maximum | 167 | nan | 6.42 | 0.93 | 5.2 | 5.8 | 7.1 |
| scheduled_glc | 30 | minimum | 99 | nan | 5.86 | 1 | 4.9 | 5.6 | 6.4 |
| scheduled_glc | 180 | minimum | 110 | nan | 5.76 | 0.99 | 4.9 | 5.4 | 6.3 |
| scheduled_glc | 365 | minimum | 114 | nan | 5.63 | 0.98 | 4.8 | 5.4 | 6.2 |
| scheduled_glc | 730 | minimum | 116 | nan | 5.57 | 0.96 | 4.8 | 5.3 | 6.1 |
| scheduled_glc | 1460 | mean | 910 | nan | 5.89 | 0.94 | 5.1 | 5.6 | 6.4 |
| scheduled_glc | 1460 | minimum | 115 | nan | 5.53 | 0.94 | 4.8 | 5.3 | 6.1 |
| scheduled_glc | 30 | mean | 322 | nan | 6.13 | 1 | 5.2 | 5.8 | 6.8 |
| scheduled_glc | 180 | mean | 592 | nan | 6.08 | 0.99 | 5.2 | 5.7 | 6.7 |
| scheduled_glc | 365 | mean | 717 | nan | 5.98 | 0.98 | 5.1 | 5.6 | 6.6 |
| scheduled_glc | 730 | mean | 834 | nan | 5.92 | 0.96 | 5.1 | 5.6 | 6.5 |
| scheduled_glc | 1095 | minimum | 115 | nan | 5.54 | 0.95 | 4.8 | 5.3 | 6.1 |
| selected_nassa | 1460 | maximum | 2 | 0 | 0.06 | 0.94 | 0 | 0 | 0 |
| selected_nassa | 1095 | maximum | 2 | 0 | 0.06 | 0.94 | 0 | 0 | 0 |
| selected_nassa | 730 | maximum | 2 | 0 | 0.05 | 0.95 | 0 | 0 | 0 |
| selected_nassa | 365 | maximum | 2 | 0 | 0.04 | 0.96 | 0 | 0 | 0 |
| selected_nassa | 180 | maximum | 2 | 0 | 0.02 | 0.98 | 0 | 0 | 0 |
| selected_nassa | 1460 | latest | 2 | 0 | 0.06 | 0.94 | 0 | 0 | 0 |
| selected_nassa | 1825 | latest | 2 | 0 | 0.07 | 0.93 | 0 | 0 | 0 |
| selected_nassa | 1095 | latest | 2 | 0 | 0.06 | 0.94 | 0 | 0 | 0 |
| selected_nassa | 730 | latest | 2 | 0 | 0.05 | 0.95 | 0 | 0 | 0 |
| selected_nassa | 365 | latest | 2 | 0 | 0.04 | 0.96 | 0 | 0 | 0 |
| selected_nassa | 1825 | maximum | 2 | 0 | 0.07 | 0.93 | 0 | 0 | 0 |
| selected_nassa | 30 | maximum | 2 | 0 | 0.01 | 0.99 | 0 | 0 | 0 |
| selected_nassa | 180 | minimum | 2 | 0 | 0.02 | 0.98 | 0 | 0 | 0 |
| selected_nassa | 180 | mean | 2 | 0 | 0.02 | 0.98 | 0 | 0 | 0 |
| selected_nassa | 730 | minimum | 2 | 0 | 0.05 | 0.95 | 0 | 0 | 0 |
| selected_nassa | 1095 | minimum | 2 | 0 | 0.06 | 0.94 | 0 | 0 | 0 |

| | | | | | | | | | |
|---|---|---|---|---|---|---|---|---|---|
| selected_nassa | 1460 | minimum | 2 | 0 | 0.06 | 0.94 | 0 | 0 | 0 |
| selected_nassa | 1825 | minimum | 2 | 0 | 0.07 | 0.93 | 0 | 0 | 0 |
| selected_nassa | 30 | mean | 2 | 0 | 0.01 | 0.99 | 0 | 0 | 0 |
| selected_nassa | 30 | minimum | 2 | 0 | 0.01 | 0.99 | 0 | 0 | 0 |
| selected_nassa | 365 | mean | 2 | 0 | 0.04 | 0.96 | 0 | 0 | 0 |
| selected_nassa | 730 | mean | 2 | 0 | 0.05 | 0.95 | 0 | 0 | 0 |
| selected_nassa | 1095 | mean | 2 | 0 | 0.06 | 0.94 | 0 | 0 | 0 |
| selected_nassa | 1460 | mean | 2 | 0 | 0.06 | 0.94 | 0 | 0 | 0 |
| selected_nassa | 1825 | mean | 2 | 0 | 0.07 | 0.93 | 0 | 0 | 0 |
| selected_nassa | 30 | latest | 2 | 0 | 0.01 | 0.99 | 0 | 0 | 0 |
| selected_nassa | 365 | minimum | 2 | 0 | 0.04 | 0.96 | 0 | 0 | 0 |
| selected_nassa | 180 | latest | 2 | 0 | 0.02 | 0.98 | 0 | 0 | 0 |
| sex_female | nan | nan | 2 | nan | 0.62 | nan | nan | nan | nan |
| sleep_apnea | 1460 | latest | 2 | 0 | 0.01 | 0.99 | 0 | 0 | 0 |
| sleep_apnea | 1825 | mean | 2 | 0 | 0.01 | 0.99 | 0 | 0 | 0 |
| sleep_apnea | 180 | mean | 2 | 0 | 0 | 1 | 0 | 0 | 0 |
| sleep_apnea | 365 | mean | 2 | 0 | 0.01 | 0.99 | 0 | 0 | 0 |
| sleep_apnea | 730 | mean | 2 | 0 | 0.01 | 0.99 | 0 | 0 | 0 |
| sleep_apnea | 1095 | mean | 2 | 0 | 0.01 | 0.99 | 0 | 0 | 0 |
| sleep_apnea | 1460 | mean | 2 | 0 | 0.01 | 0.99 | 0 | 0 | 0 |
| sleep_apnea | 1095 | latest | 2 | 0 | 0.01 | 0.99 | 0 | 0 | 0 |
| sleep_apnea | 180 | latest | 2 | 0 | 0 | 1 | 0 | 0 | 0 |
| sleep_apnea | 365 | latest | 2 | 0 | 0.01 | 0.99 | 0 | 0 | 0 |
| sleep_apnea | 730 | latest | 2 | 0 | 0.01 | 0.99 | 0 | 0 | 0 |
| sleep_apnea | 1825 | minimum | 2 | 0 | 0.01 | 0.99 | 0 | 0 | 0 |
| sleep_apnea | 30 | mean | 2 | 0 | 0 | 1 | 0 | 0 | 0 |
| sleep_apnea | 30 | maximum | 2 | 0 | 0 | 1 | 0 | 0 | 0 |
| sleep_apnea | 30 | latest | 2 | 0 | 0 | 1 | 0 | 0 | 0 |

| | | | | | | | | | |
|---|---|---|---|---|---|---|---|---|---|
| sleep_apnea | 1095 | minimum | 2 | 0 | 0.01 | 0.99 | 0 | 0 | 0 |
| sleep_apnea | 1460 | minimum | 2 | 0 | 0.01 | 0.99 | 0 | 0 | 0 |
| sleep_apnea | 1825 | latest | 2 | 0 | 0.01 | 0.99 | 0 | 0 | 0 |
| sleep_apnea | 180 | maximum | 2 | 0 | 0 | 1 | 0 | 0 | 0 |
| sleep_apnea | 365 | maximum | 2 | 0 | 0.01 | 0.99 | 0 | 0 | 0 |
| sleep_apnea | 1095 | maximum | 2 | 0 | 0.01 | 0.99 | 0 | 0 | 0 |
| sleep_apnea | 1460 | maximum | 2 | 0 | 0.01 | 0.99 | 0 | 0 | 0 |
| sleep_apnea | 730 | maximum | 2 | 0 | 0.01 | 0.99 | 0 | 0 | 0 |
| sleep_apnea | 30 | minimum | 2 | 0 | 0 | 1 | 0 | 0 | 0 |
| sleep_apnea | 180 | minimum | 2 | 0 | 0 | 1 | 0 | 0 | 0 |
| sleep_apnea | 365 | minimum | 2 | 0 | 0.01 | 0.99 | 0 | 0 | 0 |
| sleep_apnea | 730 | minimum | 2 | 0 | 0.01 | 0.99 | 0 | 0 | 0 |
| sleep_apnea | 1825 | maximum | 2 | 0 | 0.01 | 0.99 | 0 | 0 | 0 |
| snri | 1095 | mean | 2 | 0 | 0.09 | 0.91 | 0 | 0 | 0 |
| snri | 365 | latest | 2 | 0 | 0.06 | 0.94 | 0 | 0 | 0 |
| snri | 1825 | mean | 2 | 0 | 0.11 | 0.89 | 0 | 0 | 0 |
| snri | 30 | latest | 2 | 0 | 0.01 | 0.99 | 0 | 0 | 0 |
| snri | 180 | latest | 2 | 0 | 0.04 | 0.96 | 0 | 0 | 0 |
| snri | 730 | latest | 2 | 0 | 0.08 | 0.92 | 0 | 0 | 0 |
| snri | 1460 | latest | 2 | 0 | 0.1 | 0.9 | 0 | 0 | 0 |
| snri | 180 | maximum | 2 | 0 | 0.04 | 0.96 | 0 | 0 | 0 |
| snri | 30 | mean | 2 | 0 | 0.01 | 0.99 | 0 | 0 | 0 |
| snri | 30 | maximum | 2 | 0 | 0.01 | 0.99 | 0 | 0 | 0 |
| snri | 1825 | latest | 2 | 0 | 0.11 | 0.89 | 0 | 0 | 0 |
| snri | 730 | mean | 2 | 0 | 0.08 | 0.92 | 0 | 0 | 0 |
| snri | 1095 | latest | 2 | 0 | 0.09 | 0.91 | 0 | 0 | 0 |
| snri | 365 | mean | 2 | 0 | 0.06 | 0.94 | 0 | 0 | 0 |
| snri | 180 | mean | 2 | 0 | 0.04 | 0.96 | 0 | 0 | 0 |

| | | | | | | | | | |
|---|---|---|---|---|---|---|---|---|---|
| snri | 1460 | mean | 2 | 0 | 0.1 | 0.9 | 0 | 0 | 0 |
| snri | 730 | minimum | 2 | 0 | 0.08 | 0.92 | 0 | 0 | 0 |
| snri | 365 | minimum | 2 | 0 | 0.06 | 0.94 | 0 | 0 | 0 |
| snri | 180 | minimum | 2 | 0 | 0.04 | 0.96 | 0 | 0 | 0 |
| snri | 1460 | maximum | 2 | 0 | 0.1 | 0.9 | 0 | 0 | 0 |
| snri | 1825 | maximum | 2 | 0 | 0.11 | 0.89 | 0 | 0 | 0 |
| snri | 1095 | minimum | 2 | 0 | 0.09 | 0.91 | 0 | 0 | 0 |
| snri | 730 | maximum | 2 | 0 | 0.08 | 0.92 | 0 | 0 | 0 |
| snri | 1460 | minimum | 2 | 0 | 0.1 | 0.9 | 0 | 0 | 0 |
| snri | 365 | maximum | 2 | 0 | 0.06 | 0.94 | 0 | 0 | 0 |
| snri | 30 | minimum | 2 | 0 | 0.01 | 0.99 | 0 | 0 | 0 |
| snri | 1825 | minimum | 2 | 0 | 0.11 | 0.89 | 0 | 0 | 0 |
| snri | 1095 | maximum | 2 | 0 | 0.09 | 0.91 | 0 | 0 | 0 |
| ssri | 730 | mean | 2 | 0 | 0.12 | 0.88 | 0 | 0 | 0 |
| ssri | 1095 | mean | 2 | 0 | 0.15 | 0.85 | 0 | 0 | 0 |
| ssri | 1460 | mean | 2 | 0 | 0.16 | 0.84 | 0 | 0 | 0 |
| ssri | 1825 | mean | 2 | 0 | 0.17 | 0.83 | 0 | 0 | 0 |
| ssri | 30 | latest | 2 | 0 | 0.02 | 0.98 | 0 | 0 | 0 |
| ssri | 1095 | latest | 2 | 0 | 0.15 | 0.85 | 0 | 0 | 0 |
| ssri | 365 | latest | 2 | 0 | 0.09 | 0.91 | 0 | 0 | 0 |
| ssri | 1825 | latest | 2 | 0 | 0.17 | 0.83 | 0 | 0 | 0 |
| ssri | 1825 | minimum | 2 | 0 | 0.17 | 0.83 | 0 | 0 | 0 |
| ssri | 730 | latest | 2 | 0 | 0.12 | 0.88 | 0 | 0 | 0 |
| ssri | 365 | mean | 2 | 0 | 0.09 | 0.91 | 0 | 0 | 0 |
| ssri | 180 | latest | 2 | 0 | 0.06 | 0.94 | 0 | 0 | 0 |
| ssri | 180 | mean | 2 | 0 | 0.06 | 0.94 | 0 | 0 | 0 |
| ssri | 30 | mean | 2 | 0 | 0.02 | 0.98 | 0 | 0 | 0 |
| ssri | 1460 | latest | 2 | 0 | 0.16 | 0.84 | 0 | 0 | 0 |

| | | | | | | | | | |
|---|---|---|---|---|---|---|---|---|---|
| ssri | 1460 | minimum | 2 | 0 | 0.16 | 0.84 | 0 | 0 | 0 |
| ssri | 30 | maximum | 2 | 0 | 0.02 | 0.98 | 0 | 0 | 0 |
| ssri | 180 | maximum | 2 | 0 | 0.06 | 0.94 | 0 | 0 | 0 |
| ssri | 365 | maximum | 2 | 0 | 0.09 | 0.91 | 0 | 0 | 0 |
| ssri | 1095 | maximum | 2 | 0 | 0.15 | 0.85 | 0 | 0 | 0 |
| ssri | 1460 | maximum | 2 | 0 | 0.16 | 0.84 | 0 | 0 | 0 |
| ssri | 730 | maximum | 2 | 0 | 0.12 | 0.88 | 0 | 0 | 0 |
| ssri | 30 | minimum | 2 | 0 | 0.02 | 0.98 | 0 | 0 | 0 |
| ssri | 180 | minimum | 2 | 0 | 0.06 | 0.94 | 0 | 0 | 0 |
| ssri | 365 | minimum | 2 | 0 | 0.09 | 0.91 | 0 | 0 | 0 |
| ssri | 730 | minimum | 2 | 0 | 0.12 | 0.88 | 0 | 0 | 0 |
| ssri | 1095 | minimum | 2 | 0 | 0.15 | 0.85 | 0 | 0 | 0 |
| ssri | 1825 | maximum | 2 | 0 | 0.17 | 0.83 | 0 | 0 | 0 |
| statins | 180 | latest | 2 | 0 | 0.02 | 0.98 | 0 | 0 | 0 |
| statins | 1095 | mean | 2 | 0 | 0.03 | 0.97 | 0 | 0 | 0 |
| statins | 1460 | mean | 2 | 0 | 0.03 | 0.97 | 0 | 0 | 0 |
| statins | 1825 | mean | 2 | 0 | 0.03 | 0.97 | 0 | 0 | 0 |
| statins | 30 | latest | 2 | 0 | 0.01 | 0.99 | 0 | 0 | 0 |
| statins | 365 | latest | 2 | 0 | 0.02 | 0.98 | 0 | 0 | 0 |
| statins | 730 | mean | 2 | 0 | 0.03 | 0.97 | 0 | 0 | 0 |
| statins | 1095 | latest | 2 | 0 | 0.03 | 0.97 | 0 | 0 | 0 |
| statins | 1460 | latest | 2 | 0 | 0.03 | 0.97 | 0 | 0 | 0 |
| statins | 1825 | latest | 2 | 0 | 0.03 | 0.97 | 0 | 0 | 0 |
| statins | 365 | mean | 2 | 0 | 0.02 | 0.98 | 0 | 0 | 0 |
| statins | 180 | maximum | 2 | 0 | 0.02 | 0.98 | 0 | 0 | 0 |
| statins | 730 | latest | 2 | 0 | 0.03 | 0.97 | 0 | 0 | 0 |
| statins | 180 | mean | 2 | 0 | 0.02 | 0.98 | 0 | 0 | 0 |
| statins | 1825 | minimum | 2 | 0 | 0.03 | 0.97 | 0 | 0 | 0 |

| | | | | | | | | | |
|---|---|---|---|---|---|---|---|---|---|
| statins | 30 | mean | 2 | 0 | 0.01 | 0.99 | 0 | 0 | 0 |
| statins | 30 | maximum | 2 | 0 | 0.01 | 0.99 | 0 | 0 | 0 |
| statins | 365 | maximum | 2 | 0 | 0.02 | 0.98 | 0 | 0 | 0 |
| statins | 730 | maximum | 2 | 0 | 0.03 | 0.97 | 0 | 0 | 0 |
| statins | 1460 | maximum | 2 | 0 | 0.03 | 0.97 | 0 | 0 | 0 |
| statins | 1825 | maximum | 2 | 0 | 0.03 | 0.97 | 0 | 0 | 0 |
| statins | 1095 | maximum | 2 | 0 | 0.03 | 0.97 | 0 | 0 | 0 |
| statins | 180 | minimum | 2 | 0 | 0.02 | 0.98 | 0 | 0 | 0 |
| statins | 365 | minimum | 2 | 0 | 0.02 | 0.98 | 0 | 0 | 0 |
| statins | 730 | minimum | 2 | 0 | 0.03 | 0.97 | 0 | 0 | 0 |
| statins | 1095 | minimum | 2 | 0 | 0.03 | 0.97 | 0 | 0 | 0 |
| statins | 1460 | minimum | 2 | 0 | 0.03 | 0.97 | 0 | 0 | 0 |
| statins | 30 | minimum | 2 | 0 | 0.01 | 0.99 | 0 | 0 | 0 |
| tca | 30 | mean | 2 | 0 | 0.01 | 0.99 | 0 | 0 | 0 |
| tca | 180 | mean | 2 | 0 | 0.02 | 0.98 | 0 | 0 | 0 |
| tca | 1095 | latest | 2 | 0 | 0.05 | 0.95 | 0 | 0 | 0 |
| tca | 1460 | mean | 2 | 0 | 0.05 | 0.95 | 0 | 0 | 0 |
| tca | 730 | latest | 2 | 0 | 0.04 | 0.96 | 0 | 0 | 0 |
| tca | 180 | latest | 2 | 0 | 0.02 | 0.98 | 0 | 0 | 0 |
| tca | 365 | mean | 2 | 0 | 0.03 | 0.97 | 0 | 0 | 0 |
| tca | 1825 | mean | 2 | 0 | 0.05 | 0.95 | 0 | 0 | 0 |
| tca | 365 | latest | 2 | 0 | 0.03 | 0.97 | 0 | 0 | 0 |
| tca | 1095 | mean | 2 | 0 | 0.05 | 0.95 | 0 | 0 | 0 |
| tca | 730 | mean | 2 | 0 | 0.04 | 0.96 | 0 | 0 | 0 |
| tca | 365 | minimum | 2 | 0 | 0.03 | 0.97 | 0 | 0 | 0 |
| tca | 30 | latest | 2 | 0 | 0.01 | 0.99 | 0 | 0 | 0 |
| tca | 1825 | minimum | 2 | 0 | 0.05 | 0.95 | 0 | 0 | 0 |
| tca | 30 | minimum | 2 | 0 | 0.01 | 0.99 | 0 | 0 | 0 |

| | | | | | | | | | |
|---|---|---|---|---|---|---|---|---|---|
| tca | 1095 | minimum | 2 | 0 | 0.05 | 0.95 | 0 | 0 | 0 |
| tca | 30 | maximum | 2 | 0 | 0.01 | 0.99 | 0 | 0 | 0 |
| tca | 180 | maximum | 2 | 0 | 0.02 | 0.98 | 0 | 0 | 0 |
| tca | 1460 | latest | 2 | 0 | 0.05 | 0.95 | 0 | 0 | 0 |
| tca | 365 | maximum | 2 | 0 | 0.03 | 0.97 | 0 | 0 | 0 |
| tca | 730 | maximum | 2 | 0 | 0.04 | 0.96 | 0 | 0 | 0 |
| tca | 1460 | minimum | 2 | 0 | 0.05 | 0.95 | 0 | 0 | 0 |
| tca | 1095 | maximum | 2 | 0 | 0.05 | 0.95 | 0 | 0 | 0 |
| tca | 1460 | maximum | 2 | 0 | 0.05 | 0.95 | 0 | 0 | 0 |
| tca | 1825 | maximum | 2 | 0 | 0.05 | 0.95 | 0 | 0 | 0 |
| tca | 180 | minimum | 2 | 0 | 0.02 | 0.98 | 0 | 0 | 0 |
| tca | 730 | minimum | 2 | 0 | 0.04 | 0.96 | 0 | 0 | 0 |
| tca | 1825 | latest | 2 | 0 | 0.05 | 0.95 | 0 | 0 | 0 |
| top_10_weight_gaining_antipsychotics | 1825 | maximum | 2 | 0 | 0.28 | 0.72 | 0 | 0 | 1 |
| top_10_weight_gaining_antipsychotics | 1460 | maximum | 2 | 0 | 0.28 | 0.72 | 0 | 0 | 1 |
| top_10_weight_gaining_antipsychotics | 1095 | maximum | 2 | 0 | 0.26 | 0.74 | 0 | 0 | 1 |
| top_10_weight_gaining_antipsychotics | 730 | maximum | 2 | 0 | 0.24 | 0.76 | 0 | 0 | 0 |
| top_10_weight_gaining_antipsychotics | 365 | maximum | 2 | 0 | 0.19 | 0.81 | 0 | 0 | 0 |
| top_10_weight_gaining_antipsychotics | 1460 | minimum | 2 | 0 | 0.28 | 0.72 | 0 | 0 | 1 |
| top_10_weight_gaining_antipsychotics | 1095 | latest | 2 | 0 | 0.26 | 0.74 | 0 | 0 | 1 |
| top_10_weight_gaining_antipsychotics | 1460 | latest | 2 | 0 | 0.28 | 0.72 | 0 | 0 | 1 |

| | | | | | | | | | |
|---|---|---|---|---|---|---|---|---|---|
| top_10_weight_gaining_antipsychotics | 180 | minimum | 2 | 0 | 0.15 | 0.85 | 0 | 0 | 0 |
| top_10_weight_gaining_antipsychotics | 1825 | latest | 2 | 0 | 0.28 | 0.72 | 0 | 0 | 1 |
| top_10_weight_gaining_antipsychotics | 30 | minimum | 2 | 0 | 0.06 | 0.94 | 0 | 0 | 0 |
| top_10_weight_gaining_antipsychotics | 30 | maximum | 2 | 0 | 0.06 | 0.94 | 0 | 0 | 0 |
| top_10_weight_gaining_antipsychotics | 365 | minimum | 2 | 0 | 0.19 | 0.81 | 0 | 0 | 0 |
| top_10_weight_gaining_antipsychotics | 730 | minimum | 2 | 0 | 0.24 | 0.76 | 0 | 0 | 0 |
| top_10_weight_gaining_antipsychotics | 30 | mean | 2 | 0 | 0.06 | 0.94 | 0 | 0 | 0 |
| top_10_weight_gaining_antipsychotics | 1095 | minimum | 2 | 0 | 0.26 | 0.74 | 0 | 0 | 1 |
| top_10_weight_gaining_antipsychotics | 365 | latest | 2 | 0 | 0.19 | 0.81 | 0 | 0 | 0 |
| top_10_weight_gaining_antipsychotics | 180 | latest | 2 | 0 | 0.15 | 0.85 | 0 | 0 | 0 |
| top_10_weight_gaining_antipsychotics | 30 | latest | 2 | 0 | 0.06 | 0.94 | 0 | 0 | 0 |
| top_10_weight_gaining_antipsychotics | 730 | latest | 2 | 0 | 0.24 | 0.76 | 0 | 0 | 0 |
| top_10_weight_gaining_antipsychotics | 1460 | mean | 2 | 0 | 0.28 | 0.72 | 0 | 0 | 1 |
| top_10_weight_gaining_antipsychotics | 1825 | mean | 2 | 0 | 0.28 | 0.72 | 0 | 0 | 1 |
| top_10_weight_gaining_antipsychotics | 730 | mean | 2 | 0 | 0.24 | 0.76 | 0 | 0 | 0 |
| top_10_weight_gaining_antipsychotics | 365 | mean | 2 | 0 | 0.19 | 0.81 | 0 | 0 | 0 |

| | | | | | | | | | |
|---|---|---|---|---|---|---|---|---|---|
| top_10_weight_gaining_antipsychotics | 180 | mean | 2 | 0 | 0.15 | 0.85 | 0 | 0 | 0 |
| top_10_weight_gaining_antipsychotics | 1825 | minimum | 2 | 0 | 0.28 | 0.72 | 0 | 0 | 1 |
| top_10_weight_gaining_antipsychotics | 180 | maximum | 2 | 0 | 0.15 | 0.85 | 0 | 0 | 0 |
| top_10_weight_gaining_antipsychotics | 1095 | mean | 2 | 0 | 0.26 | 0.74 | 0 | 0 | 1 |
| triglycerides | 1460 | maximum | 650 | nan | 1.86 | 0.26 | 1 | 1.5 | 2.3 |
| triglycerides | 180 | maximum | 610 | nan | 1.63 | 0.6 | 0.9 | 1.3 | 2 |
| triglycerides | 365 | maximum | 615 | nan | 1.69 | 0.43 | 0.9 | 1.4 | 2 |
| triglycerides | 730 | maximum | 633 | nan | 1.77 | 0.32 | 1 | 1.4 | 2.1 |
| triglycerides | 1095 | maximum | 647 | nan | 1.83 | 0.28 | 1 | 1.5 | 2.2 |
| triglycerides | 30 | maximum | 556 | nan | 1.58 | 0.91 | 0.9 | 1.3 | 1.9 |
| triglycerides | 1460 | minimum | 504 | nan | 1.2 | 0.26 | 0.7 | 1 | 1.4 |
| triglycerides | 30 | minimum | 551 | nan | 1.56 | 0.91 | 0.9 | 1.3 | 1.9 |
| triglycerides | 180 | minimum | 577 | nan | 1.47 | 0.6 | 0.8 | 1.2 | 1.8 |
| triglycerides | 365 | minimum | 557 | nan | 1.4 | 0.43 | 0.8 | 1.1 | 1.7 |
| triglycerides | 730 | minimum | 536 | nan | 1.29 | 0.32 | 0.8 | 1.1 | 1.6 |
| triglycerides | 1825 | maximum | 662 | nan | 1.88 | 0.25 | 1 | 1.5 | 2.3 |
| triglycerides | 1825 | latest | 632 | nan | 1.51 | 0.25 | 0.9 | 1.2 | 1.8 |
| triglycerides | 1095 | minimum | 517 | nan | 1.23 | 0.28 | 0.7 | 1 | 1.5 |
| triglycerides | 1460 | latest | 629 | nan | 1.51 | 0.26 | 0.9 | 1.2 | 1.8 |
| triglycerides | 1825 | minimum | 509 | nan | 1.18 | 0.25 | 0.7 | 1 | 1.4 |
| triglycerides | 1460 | mean | 7251 | nan | 1.49 | 0.26 | 0.9 | 1.2 | 1.8 |
| triglycerides | 1095 | latest | 627 | nan | 1.51 | 0.28 | 0.9 | 1.2 | 1.8 |
| triglycerides | 730 | latest | 623 | nan | 1.52 | 0.32 | 0.9 | 1.2 | 1.8 |
| triglycerides | 180 | latest | 600 | nan | 1.55 | 0.6 | 0.9 | 1.3 | 1.9 |
| triglycerides | 30 | latest | 552 | nan | 1.57 | 0.91 | 0.9 | 1.3 | 1.9 |

| | | | | | | | | | |
|---|---|---|---|---|---|---|---|---|---|
| triglycerides | 365 | latest | 612 | nan | 1.53 | 0.43 | 0.9 | 1.2 | 1.9 |
| triglycerides | 1095 | mean | 6262 | nan | 1.5 | 0.28 | 0.9 | 1.2 | 1.8 |
| triglycerides | 730 | mean | 4834 | nan | 1.51 | 0.32 | 0.9 | 1.2 | 1.8 |
| triglycerides | 365 | mean | 3064 | nan | 1.53 | 0.43 | 0.9 | 1.3 | 1.8 |
| triglycerides | 180 | mean | 1944 | nan | 1.55 | 0.6 | 0.9 | 1.3 | 1.9 |
| triglycerides | 30 | mean | 760 | nan | 1.57 | 0.91 | 0.9 | 1.3 | 1.9 |
| triglycerides | 1825 | mean | 8123 | nan | 1.48 | 0.25 | 0.9 | 1.2 | 1.8 |
| unscheduled_p_glc | 180 | latest | 95 | nan | 5.61 | 0.67 | 5 | 5.4 | 6 |
| unscheduled_p_glc | 1095 | latest | 96 | nan | 5.61 | 0.37 | 5 | 5.4 | 6 |
| unscheduled_p_glc | 30 | latest | 95 | nan | 5.61 | 0.91 | 5 | 5.4 | 6 |
| unscheduled_p_glc | 730 | latest | 96 | nan | 5.61 | 0.42 | 5 | 5.4 | 6 |
| unscheduled_p_glc | 1825 | minimum | 98 | nan | 5.19 | 0.32 | 4.7 | 5.1 | 5.6 |
| unscheduled_p_glc | 365 | latest | 95 | nan | 5.61 | 0.54 | 5 | 5.4 | 6 |
| unscheduled_p_glc | 180 | mean | 2168 | nan | 5.62 | 0.67 | 5.1 | 5.5 | 6 |
| unscheduled_p_glc | 1460 | mean | 5526 | nan | 5.63 | 0.33 | 5.1 | 5.5 | 6 |
| unscheduled_p_glc | 1095 | mean | 5077 | nan | 5.63 | 0.37 | 5.1 | 5.5 | 6 |
| unscheduled_p_glc | 730 | mean | 4326 | nan | 5.63 | 0.42 | 5.1 | 5.5 | 6 |
| unscheduled_p_glc | 365 | mean | 3040 | nan | 5.62 | 0.54 | 5.1 | 5.5 | 6 |
| unscheduled_p_glc | 1460 | latest | 96 | nan | 5.61 | 0.33 | 5 | 5.5 | 6 |
| unscheduled_p_glc | 1825 | mean | 5717 | nan | 5.63 | 0.32 | 5.1 | 5.5 | 6 |
| unscheduled_p_glc | 1825 | latest | 96 | nan | 5.61 | 0.32 | 5 | 5.5 | 6 |
| unscheduled_p_glc | 1460 | maximum | 88 | nan | 6.16 | 0.33 | 5.3 | 5.9 | 6.8 |
| unscheduled_p_glc | 1460 | minimum | 97 | nan | 5.21 | 0.33 | 4.7 | 5.1 | 5.6 |
| unscheduled_p_glc | 30 | mean | 780 | nan | 5.61 | 0.91 | 5 | 5.5 | 6.1 |
| unscheduled_p_glc | 730 | minimum | 97 | nan | 5.3 | 0.42 | 4.8 | 5.2 | 5.7 |
| unscheduled_p_glc | 365 | minimum | 96 | nan | 5.38 | 0.54 | 4.9 | 5.3 | 5.8 |
| unscheduled_p_glc | 180 | minimum | 96 | nan | 5.45 | 0.67 | 4.9 | 5.3 | 5.9 |
| unscheduled_p_glc | 1825 | maximum | 88 | nan | 6.2 | 0.32 | 5.3 | 5.9 | 6.8 |

| | | | | | | | | | |
|---|---|---|---|---|---|---|---|---|---|
| unscheduled_p_glc | 30 | minimum | 96 | nan | 5.53 | 0.91 | 5 | 5.4 | 5.9 |
| unscheduled_p_glc | 1095 | maximum | 88 | nan | 6.11 | 0.37 | 5.3 | 5.8 | 6.7 |
| unscheduled_p_glc | 365 | maximum | 90 | nan | 5.91 | 0.54 | 5.2 | 5.7 | 6.4 |
| unscheduled_p_glc | 180 | maximum | 92 | nan | 5.82 | 0.67 | 5.1 | 5.6 | 6.3 |
| unscheduled_p_glc | 30 | maximum | 91 | nan | 5.7 | 0.91 | 5 | 5.5 | 6.2 |
| unscheduled_p_glc | 730 | maximum | 90 | nan | 6.03 | 0.42 | 5.2 | 5.7 | 6.6 |
| unscheduled_p_glc | 1095 | minimum | 97 | nan | 5.24 | 0.37 | 4.8 | 5.2 | 5.6 |
| urinary_glc | 1825 | mean | 96 | nan | 0.02 | 0.78 | 0 | 0 | 0 |
| urinary_glc | 365 | mean | 50 | nan | 0.02 | 0.9 | 0 | 0 | 0 |
| urinary_glc | 730 | mean | 70 | nan | 0.02 | 0.85 | 0 | 0 | 0 |
| urinary_glc | 1095 | mean | 83 | nan | 0.02 | 0.81 | 0 | 0 | 0 |
| urinary_glc | 30 | latest | 5 | nan | 0.03 | 0.98 | 0 | 0 | 0 |
| urinary_glc | 365 | latest | 5 | nan | 0.02 | 0.9 | 0 | 0 | 0 |
| urinary_glc | 730 | latest | 5 | nan | 0.02 | 0.85 | 0 | 0 | 0 |
| urinary_glc | 1095 | latest | 5 | nan | 0.02 | 0.81 | 0 | 0 | 0 |
| urinary_glc | 1825 | latest | 5 | nan | 0.02 | 0.78 | 0 | 0 | 0 |
| urinary_glc | 1825 | minimum | 5 | nan | 0.01 | 0.78 | 0 | 0 | 0 |
| urinary_glc | 180 | mean | 39 | nan | 0.02 | 0.94 | 0 | 0 | 0 |
| urinary_glc | 180 | latest | 5 | nan | 0.02 | 0.94 | 0 | 0 | 0 |
| urinary_glc | 30 | mean | 16 | nan | 0.03 | 0.98 | 0 | 0 | 0 |
| urinary_glc | 1460 | latest | 5 | nan | 0.02 | 0.79 | 0 | 0 | 0 |
| urinary_glc | 1460 | minimum | 5 | nan | 0.01 | 0.79 | 0 | 0 | 0 |
| urinary_glc | 1460 | mean | 95 | nan | 0.02 | 0.79 | 0 | 0 | 0 |
| urinary_glc | 730 | minimum | 5 | nan | 0.01 | 0.85 | 0 | 0 | 0 |
| urinary_glc | 365 | minimum | 5 | nan | 0.01 | 0.9 | 0 | 0 | 0 |
| urinary_glc | 180 | minimum | 4 | nan | 0.02 | 0.94 | 0 | 0 | 0 |
| urinary_glc | 1825 | maximum | 5 | nan | 0.04 | 0.78 | 0 | 0 | 0 |
| urinary_glc | 30 | maximum | 5 | nan | 0.03 | 0.98 | 0 | 0 | 0 |

| | | | | | | | | | |
|---|---|---|---|---|---|---|---|---|---|
| urinary_glc | 30 | minimum | 5 | nan | 0.02 | 0.98 | 0 | 0 | 0 |
| urinary_glc | 730 | maximum | 5 | nan | 0.04 | 0.85 | 0 | 0 | 0 |
| urinary_glc | 365 | maximum | 5 | nan | 0.03 | 0.9 | 0 | 0 | 0 |
| urinary_glc | 180 | maximum | 5 | nan | 0.03 | 0.94 | 0 | 0 | 0 |
| urinary_glc | 1095 | minimum | 5 | nan | 0.01 | 0.81 | 0 | 0 | 0 |
| urinary_glc | 1460 | maximum | 5 | nan | 0.04 | 0.79 | 0 | 0 | 0 |
| urinary_glc | 1095 | maximum | 5 | nan | 0.04 | 0.81 | 0 | 0 | 0 |
| valproate | 730 | latest | 2 | 0 | 0.02 | 0.98 | 0 | 0 | 0 |
| valproate | 180 | latest | 2 | 0 | 0.01 | 0.99 | 0 | 0 | 0 |
| valproate | 730 | mean | 2 | 0 | 0.02 | 0.98 | 0 | 0 | 0 |
| valproate | 1095 | latest | 2 | 0 | 0.03 | 0.97 | 0 | 0 | 0 |
| valproate | 365 | latest | 2 | 0 | 0.02 | 0.98 | 0 | 0 | 0 |
| valproate | 1460 | mean | 2 | 0 | 0.03 | 0.97 | 0 | 0 | 0 |
| valproate | 30 | mean | 2 | 0 | 0 | 1 | 0 | 0 | 0 |
| valproate | 180 | mean | 2 | 0 | 0.01 | 0.99 | 0 | 0 | 0 |
| valproate | 365 | mean | 2 | 0 | 0.02 | 0.98 | 0 | 0 | 0 |
| valproate | 1825 | mean | 2 | 0 | 0.03 | 0.97 | 0 | 0 | 0 |
| valproate | 1095 | mean | 2 | 0 | 0.03 | 0.97 | 0 | 0 | 0 |
| valproate | 1825 | latest | 2 | 0 | 0.03 | 0.97 | 0 | 0 | 0 |
| valproate | 30 | latest | 2 | 0 | 0 | 1 | 0 | 0 | 0 |
| valproate | 1825 | minimum | 2 | 0 | 0.03 | 0.97 | 0 | 0 | 0 |
| valproate | 365 | minimum | 2 | 0 | 0.02 | 0.98 | 0 | 0 | 0 |
| valproate | 1095 | minimum | 2 | 0 | 0.03 | 0.97 | 0 | 0 | 0 |
| valproate | 30 | maximum | 2 | 0 | 0 | 1 | 0 | 0 | 0 |
| valproate | 1460 | latest | 2 | 0 | 0.03 | 0.97 | 0 | 0 | 0 |
| valproate | 1460 | minimum | 2 | 0 | 0.03 | 0.97 | 0 | 0 | 0 |
| valproate | 365 | maximum | 2 | 0 | 0.02 | 0.98 | 0 | 0 | 0 |
| valproate | 730 | maximum | 2 | 0 | 0.02 | 0.98 | 0 | 0 | 0 |

| | | | | | | | | | |
|---|---|---|---|---|---|---|---|---|---|
| valproate | 180 | maximum | 2 | 0 | 0.01 | 0.99 | 0 | 0 | 0 |
| valproate | 1460 | maximum | 2 | 0 | 0.03 | 0.97 | 0 | 0 | 0 |
| valproate | 1825 | maximum | 2 | 0 | 0.03 | 0.97 | 0 | 0 | 0 |
| valproate | 30 | minimum | 2 | 0 | 0 | 1 | 0 | 0 | 0 |
| valproate | 180 | minimum | 2 | 0 | 0.01 | 0.99 | 0 | 0 | 0 |
| valproate | 730 | minimum | 2 | 0 | 0.02 | 0.98 | 0 | 0 | 0 |
| valproate | 1095 | maximum | 2 | 0 | 0.03 | 0.97 | 0 | 0 | 0 |
| weight_in_kg | 730 | latest | 2066 | nan | 78.93 | 0.25 | 63 | 76 | 91 |
| weight_in_kg | 365 | latest | 2052 | nan | 78.99 | 0.36 | 63 | 76 | 91 |
| weight_in_kg | 30 | latest | 1963 | nan | 75.46 | 0.83 | 58.5 | 71.9 | 88.9 |
| weight_in_kg | 180 | latest | 2047 | nan | 78.71 | 0.5 | 62.6 | 75.7 | 91 |
| weight_in_kg | 1825 | latest | 2077 | nan | 78.75 | 0.17 | 63 | 75.7 | 90.4 |
| weight_in_kg | 1460 | latest | 2074 | nan | 78.8 | 0.18 | 63 | 75.8 | 90.6 |
| weight_in_kg | 1095 | latest | 2071 | nan | 78.87 | 0.2 | 63 | 76 | 90.9 |

**ALAT**: Alanine aminotransferase. **p-Glc**: Plasma glucose. **BMI**: Body mass index. **CRP**: C-reactive protein. **EGFR**: Estimated glomerular filtration rate. **LDL**: Low-density lipoprotein. **GERD**: Gastro-esophageal reflux disorder. **HbA1c**: Hemoglobin A1c. **HDL**: High-density lipoprotein. **OGTT**: Oral glucose-tolerance test. **Glc**: Glucose. **NASSA**: Noradrenergic and specific serotonergic antidepressants. **SNRI**: Serotonin and noradrenaline reuptake inhibitor. **SSRI**: Selective serotonin reuptake inhibitor. **TCA**: Tricyclic antidepressants.

**Supplementary Table 1.** ICD-10 codes for diabetes

| | |
|---|---|
| E11.*, E12.*, E13.*, E14.*, E15.* | Diabetes |
| E16.0-E16.2 | Diabetic mononeuropathy |
| G63.2 | Diabetic polyneuropathy |
| H28.0 | Diabetic cataract |
| H33.4B | Diabetic retinal amotio |
| H36.0 | Diabetic retinopathy |
| M14.2 | Diabetic arthropathy |
| N08.3 | Diabetic glomerulonephropathy |
| O24.* | Diabetes mellitus in pregnancy |
| T38.3A | Insulin shock |

**Supplementary Table 3.** Hyperparameters for model selection. Parameters for the best performing model in bold.

| | |
|---|---|
| **Preprocessing** | |
| Imputation method | Either most frequent value, **mean**, median or no imputation. |
| Scaling | **z-score-normalisation** or no scaling |
| Predictor selection | Chi-squared or **no feature selection** |
| Predictor selection percentile[a] | Between 1 and 90 **(N/A)**. |
| Lookbehind combination (days)[b] | One of either<br>• [30, 180, 365, 730, 1095, 1460, 1825]<br>• [30, 1095, 1825]<br>• **[30, 365, 730]**<br>• [30, 365]<br>• [365] |
| **Model hyperparameters** | |
| **XGBoost** | |
| N estimators | [100; 1200], **238** |
| Alpha | [$10^{-8}$; 0.1], **$2.25 \cdot 10^{-6}$** |
| Lambda | [$10^{-8}$; 1.0], **$8.6 \cdot 10^{-7}$** |
| Max depth | [1; 10], 2 |
| Learning rate | [$10^{-8}$; 1], **0.052** |
| Gamma | [$10^{-8}$; $10^{-4}$], **0.00023** |
| Grow policy | Either depthwise or **lossguide** |
| **Logistic regression** | |
| Penalty solver | Elasticnet SAGA |
| C | [$10^{-5}$; 1] |
| L1 ratio | [$10^{-5}$; 1] |

[a]Feature selection rank-orders possible predictors by their chi-squared value. For e.g., the 10-th percentile, the top 10% of features with the highest chi-squared values were kept, the rest were dropped.

[b]Features were constructed for lookbehinds of 30, 180, 365, 730, 1095, 1460 and 1825 days. Which subset to train on was established as a hyperparameter, and one of the above combinations was selected for each training run.

**Supplementary Table 4.** The 100 most important features by information gain (XGBoost with 5 year lookahead)

| | Feature Name | Gain |
|---|---|---|
| 1 | 730-day mean HbA1c | 0.048 |
| **2** | 365-day latest weight (kg) | 0.042 |
| **3** | 730-day max HbA1c | 0.033 |
| **4** | 365-day latest triglycerides | 0.021 |
| **5** | Age (years) | 0.019 |
| **6** | 365-day max HbA1c | 0.019 |
| **7** | 730-day max HDL | 0.017 |
| **8** | 730-day mean urinary_glc | 0.017 |
| **9** | 365-day min arterial p-Glc | 0.017 |
| **10** | 365-day min HbA1c | 0.017 |
| **11** | 730-day mean ALAT | 0.016 |
| **12** | 365-day min scheduled p-Glc | 0.016 |
| **13** | 730-day min LDL | 0.016 |
| **14** | 730-day mean triglycerides | 0.016 |
| **15** | 730-day latest BMI | 0.015 |
| **16** | 365-day latest arterial p-Glc | 0.015 |
| **17** | 730-day min HbA1c | 0.015 |
| **18** | 365-day latest egfr | 0.014 |
| **19** | 730-day mean CRP | 0.014 |
| **20** | 730-day min egfr | 0.014 |
| **21** | 730-day min arterial p-Glc | 0.014 |
| **22** | 730-day max unscheduled p-Glc | 0.014 |
| **23** | 365-day mean HDL | 0.013 |
| **24** | 730-day max urinary_glc | 0.013 |
| **25** | 365-day max triglycerides | 0.013 |
| **26** | 730-day latest albumine creatinine ratio | 0.013 |
| **27** | 730-day max egfr | 0.013 |
| **28** | 730-day max albumine creatinine ratio | 0.013 |
| **29** | 730-day latest triglycerides | 0.013 |
| **30** | 730-day mean unscheduled p-Glc | 0.013 |
| **31** | 365-day latest height (cm) | 0.013 |
| **32** | 365-day mean HbA1c | 0.012 |
| **33** | 730-day min ALAT | 0.012 |
| **34** | 730-day max triglycerides | 0.012 |
| **35** | 365-day max HDL | 0.011 |
| **36** | 730-day min triglycerides | 0.011 |
| **37** | 365-day min triglycerides | 0.011 |

| | | |
|---|---|---|
| **38** | 730-day min HDL | 0.011 |
| **39** | 730-day mean LDL | 0.011 |
| **40** | 30-day max fasting LDL | 0.011 |
| **41** | 365-day max unscheduled p-Glc | 0.011 |
| **42** | 365-day min fasting p-Glc | 0.010 |
| **43** | 730-day min unscheduled p-Glc | 0.010 |
| **44** | 365-day max CRP | 0.010 |
| **45** | 365-day mean unscheduled p-Glc | 0.010 |
| **46** | 730-day min fasting p-Glc | 0.010 |
| **47** | 730-day mean HDL | 0.009 |
| **48** | 365-day max diuretics | 0.009 |
| **49** | 365-day latest fasting p-Glc | 0.009 |
| **50** | 365-day mean ALAT | 0.009 |
| **51** | 730-day max CRP | 0.009 |
| **52** | 730-day max scheduled p-Glc | 0.009 |
| **53** | 365-day min ALAT | 0.009 |
| **54** | 730-day max arterial p-Glc | 0.009 |
| **55** | 365-day max LDL | 0.009 |
| **56** | 730-day latest scheduled p-Glc | 0.009 |
| **57** | 365-day latest HDL | 0.008 |
| **58** | 730-day max LDL | 0.008 |
| **59** | 365-day latest ALAT | 0.008 |
| **60** | 730-day latest F2 disorders | 0.008 |
| **61** | 365-day latest LDL | 0.008 |
| **62** | 365-day mean F0 disorders | 0.008 |
| **63** | 730-day max fasting LDL | 0.007 |
| **64** | 365-day latest unscheduled p-Glc | 0.007 |
| **65** | 730-day latest height (cm) | 0.007 |
| **66** | 365-day min gerd_drugs | 0.007 |
| **67** | 730-day latest unscheduled p-Glc | 0.007 |
| **68** | 730-day latest HDL | 0.007 |
| **69** | 730-day latest LDL | 0.007 |
| **70** | 730-day latest weight (kg) | 0.006 |
| **71** | 365-day min HDL | 0.006 |
| **72** | 730-day latest fasting LDL | 0.006 |
| **73** | 365-day mean F3 disorders | 0.006 |
| **74** | 730-day min clozapine | 0.006 |
| **75** | 365-day max egfr | 0.006 |
| **76** | 730-day latest HbA1c | 0.006 |
| **77** | 730-day max F5 disorders | 0.005 |
| **78** | 730-day max ALAT | 0.005 |
| **79** | 730-day latest lithium | 0.005 |

| | | |
|---|---|---|
| **80** | 365-day min unscheduled p-Glc | 0.005 |
| **81** | 730-day latest ALAT | 0.005 |
| **82** | 365-day min egfr | 0.005 |
| **83** | 730-day min fasting LDL | 0.004 |
| **84** | 730-day latest F3 disorders | 0.004 |
| **85** | 365-day min LDL | 0.004 |
| **86** | 30-day latest height (cm) | 0.004 |
| **87** | 365-day min F1 disorders | 0.004 |
| **88** | 365-day mean statins | 0.003 |
| **89** | 365-day latest CRP | 0.003 |
| **90** | 365-day latest top 10 weight gaining antipsychotics | 0.003 |
| **91** | 730-day max Hyperkinetic disorders | 0.001 |
| **92** | 730-day latest F7 disorders | 0.000 |
| **93** | 730-day mean top 10 weight gaining antipsychotics | 0.000 |
| **94** | 365-day min sleep_apnea | 0.000 |
| **95** | 30-day latest albumine creatinine ratio | 0.000 |
| **96** | 30-day min gerd_drugs | 0.000 |
| **97** | 365-day mean pregabaline | 0.000 |
| **98** | 30-day min antipsychotics | 0.000 |
| **99** | 30-day mean sleep_apnea | 0.000 |
| **100** | 365-day max F6 disorders | 0.000 |

**Supplementary Table 5:** Performance by predicted positive rate of XGBoost with 1 years of lookahead.

| Predicted positive rate | True prevalence | PPV | NPV | Sens | Spec | FPR | FNR | Acc | TP | TN | FP | FN | % of all events captured | Median years from first positive to T2D |
|---|---|---|---|---|---|---|---|---|---|---|---|---|---|---|
| 5.0% | 0.1% | 1.0% | 99.9% | 45.4% | 95.0% | 5.0% | 54.6% | 95.0% | 93 | 179,566 | 9,370 | 112 | 42.9% | 2.5 |
| 4.0% | 0.1% | 1.2% | 99.9% | 43.9% | 96.0% | 4.0% | 56.1% | 96.0% | 90 | 181,460 | 7,476 | 115 | 38.1% | 2.4 |
| 3.0% | 0.1% | 1.4% | 99.9% | 39.5% | 97.0% | 3.0% | 60.5% | 97.0% | 81 | 183,342 | 5,594 | 124 | 33.3% | 2.7 |
| 2.0% | 0.1% | 2.0% | 99.9% | 36.6% | 98.0% | 2.0% | 63.4% | 98.0% | 75 | 185,226 | 3,710 | 130 | 30.2% | 2.4 |
| 1.0% | 0.1% | 3.0% | 99.9% | 28.3% | 99.0% | 1.0% | 71.7% | 98.9% | 58 | 187,089 | 1,847 | 147 | 22.2% | 2.3 |

**Predicted positive rate**: The proportion of contacts predicted positive by the model. Since the model outputs a predicted probability, this is a threshold set by us. **True prevalence**: The proportion of contacts that qualified for type 2 diabetes within the lookahead window. **PPV**: Positive predictive value. **NPV**: Negative predictive value. **FPR**: False positive rate. **FNR**: False negative rate. **TP**: True positives. Numbers are service contacts. **TN**: True negatives. Numbers are service contacts. **FP**: False positives. Numbers are service contacts. **FN**: False negatives. Numbers are service contacts. **% of all T2D captured**: Percentage of all patients who developed T2D, who had at least one positive prediction. **Median years from first positive to T2D**: For all patients with at least one true positive, the number of days from their first positive prediction to being labelled as T2D.

**Supplementary Table 6:** Performance by predicted positive rate of XGBoost with 2 years of lookahead.

| Predicted positive rate | True prevalence | PPV | NPV | Sens | Spec | FPR | FNR | Acc | TP | TN | FP | FN | % of all events captured | Median years from first positive to T2D |
|---|---|---|---|---|---|---|---|---|---|---|---|---|---|---|
| 5.0% | 0.3% | 3.6% | 99.8% | 52.6% | 95.2% | 4.8% | 47.4% | 95.0% | 251 | 132,260 | 6,722 | 226 | 43.0% | 2.4 |
| 4.0% | 0.3% | 4.4% | 99.8% | 51.8% | 96.2% | 3.8% | 48.2% | 96.0% | 247 | 133,641 | 5,341 | 230 | 41.9% | 2.4 |
| 3.0% | 0.3% | 5.1% | 99.8% | 44.4% | 97.1% | 2.9% | 55.6% | 97.0% | 212 | 135,008 | 3,974 | 265 | 33.7% | 2.3 |
| 2.0% | 0.3% | 6.7% | 99.8% | 39.6% | 98.1% | 1.9% | 60.4% | 97.9% | 189 | 136,357 | 2,625 | 288 | 30.2% | 2.1 |
| 1.0% | 0.3% | 6.5% | 99.7% | 19.1% | 99.1% | 0.9% | 80.9% | 98.8% | 91 | 137,675 | 1,307 | 386 | 18.6% | 2.1 |

**Predicted positive rate**: The proportion of contacts predicted positive by the model. Since the model outputs a predicted probability, this is a threshold set by us. **True prevalence**: The proportion of contacts that qualified for type 2 diabetes within the lookahead window. **PPV**: Positive predictive value. **NPV**: Negative predictive value. **FPR**: False positive rate. **FNR**: False negative rate. **TP**: True positives. Numbers are service contacts. **TN**: True negatives. Numbers are service contacts. **FP**: False positives. Numbers are service contacts. **FN**: False negatives. Numbers are service contacts. **% of all T2D captured**: Percentage of all patients who developed T2D, who had at least one positive prediction. **Median years from first positive to T2D**: For all patients with at least one true positive, the number of days from their first positive prediction to being labelled as T2D.

**Supplementary Table 7:** Performance by predicted positive rate of XGBoost with 3 years of lookahead.

| Predicted positive rate | True prevalence | PPV | NPV | Sens | Spec | FPR | FNR | Acc | TP | TN | FP | FN | % of all events captured | Median years from first positive to T2D |
|---|---|---|---|---|---|---|---|---|---|---|---|---|---|---|
| 5.0% | 0.7% | 5.7% | 99.6% | 43.4% | 95.3% | 4.7% | 56.6% | 94.9% | 401 | 133,374 | 6,648 | 522 | 37.9% | 2.9 |
| 4.0% | 0.7% | 6.7% | 99.6% | 41.0% | 96.2% | 3.8% | 59.0% | 95.9% | 378 | 134,760 | 5,262 | 545 | 34.8% | 2.9 |
| 3.0% | 0.7% | 8.0% | 99.6% | 36.8% | 97.2% | 2.8% | 63.2% | 96.8% | 340 | 136,128 | 3,894 | 583 | 31.8% | 2.7 |
| 2.0% | 0.7% | 9.3% | 99.5% | 28.3% | 98.2% | 1.8% | 71.7% | 97.7% | 261 | 137,464 | 2,558 | 662 | 25.8% | 2.7 |
| 1.0% | 0.7% | 10.0% | 99.4% | 15.3% | 99.1% | 0.9% | 84.7% | 98.5% | 141 | 138,752 | 1,270 | 782 | 13.6% | 2.4 |

**Predicted positive rate**: The proportion of contacts predicted positive by the model. Since the model outputs a predicted probability, this is a threshold set by us. **True prevalence**: The proportion of contacts that qualified for type 2 diabetes within the lookahead window. **PPV**: Positive predictive value. **NPV**: Negative predictive value. **FPR**: False positive rate. **FNR**: False negative rate. **TP**: True positives. Numbers are service contacts. **TN**: True negatives. Numbers are service contacts. **FP**: False positives. Numbers are service contacts. **FN**: False negatives. Numbers are service contacts. **% of all T2D captured**: Percentage of all patients who developed T2D, who had at least one positive prediction. **Median years from first positive to T2D**: For all patients with at least one true positive, the number of days from their first positive prediction to being labelled as T2D.

**Supplementary Table 8:** Performance by predicted positive rate of XGBoost with 4 years of lookahead.

| **Predicted positive rate** | **True prevalence** | **PPV** | **NPV** | **Sens** | **Spec** | **FPR** | **FNR** | **Acc** | **TP** | **TN** | **FP** | **FN** | **% of all events captured** | **Median years from first positive to T2D** |
|---|---|---|---|---|---|---|---|---|---|---|---|---|---|---|
| 5.0% | 1.1% | 10.6% | 99.4% | 48.5% | 95.5% | 4.5% | 51.5% | 95.0% | 450 | 80,458 | 3,810 | 478 | 43.0% | 3 |
| 4.0% | 1.1% | 12.4% | 99.4% | 45.7% | 96.5% | 3.5% | 54.3% | 95.9% | 424 | 81,283 | 2,985 | 504 | 39.3% | 3 |
| 3.0% | 1.1% | 14.4% | 99.3% | 39.8% | 97.4% | 2.6% | 60.2% | 96.8% | 369 | 82,080 | 2,188 | 559 | 32.7% | 2.7 |
| 2.0% | 1.1% | 11.5% | 99.1% | 21.1% | 98.2% | 1.8% | 78.9% | 97.4% | 196 | 82,759 | 1,509 | 732 | 21.5% | 2.7 |
| 1.0% | 1.1% | 15.0% | 99.1% | 13.8% | 99.1% | 0.9% | 86.2% | 98.2% | 128 | 83,544 | 724 | 800 | 15.0% | 1.9 |

**Predicted positive rate**: The proportion of contacts predicted positive by the model. Since the model outputs a predicted probability, this is a threshold set by us. **True prevalence**: The proportion of contacts that qualified for type 2 diabetes within the lookahead window. **PPV**: Positive predictive value. **NPV**: Negative predictive value. **FPR**: False positive rate. **FNR**: False negative rate. **TP**: True positives. **TN**: True negatives. **FP**: False positives. **FN**: False negatives. **% of all T2D captured**: Percentage of all patients who developed T2D, who had at least one positive prediction. **Median years from first positive to T2D**: For all patients with at least one true positive, the number of days from their first positive prediction to being labelled as T2D.

### Supplementary Discussion

Selection of wash-in periods

Since Danish guidelines recommend measuring HbA1c at least every 6 months for patients with T2D,[20] it can be argued that a 6-month wash-in should eliminate all prevalent cases. However, Supplementary Figure 3 shows that a 2-year wash-in was required for T2D incidence to stabilise. Similarly, for patients moving to the Central Denmark Region, we enforced a 2-year wash-in period in which no predictions were issued. If they met the criteria for T2D in this period, we assumed that they had been diagnosed outside of the Central Denmark Region and did not issue any further predictions.

Validity of the definition of T2D

Defining T2D in cohort studies comes with challenges, since T2D models may misclassify type 1 diabetes as T2D due to similar diagnostic criteria.[2] To address this, we excluded contacts with a prior type 1 diabetes diagnosis or insulin use, and only made predictions for patients aged ≥18 years, which should largely solve this issue.

Effects of potential model implementation

When prediction models are implemented, they should affect behaviour, for example by decreasing delay to diagnostic testing. Specifically, implementing a T2D prediction model would likely induce more relevant HbA1c-measurements. These model-induced measurements should improve the next prediction of the model, meaning that predictions following a positive prediction are likely less accurate in our dataset than they would be following implementation.
